# SARS-CoV-2 infection and risk of clinical sequelae during the post-acute phase: a retrospective cohort study

**DOI:** 10.1101/2021.03.12.21253448

**Authors:** Sarah E. Daugherty, Yinglong Guo, Kevin Heath, Micah C. Dasmariñas, Karol Giuseppe Jubilo, Jirapat Samranvedhya, Marc Lipsitch, Ken Cohen

## Abstract

**Objective:** Clinical sequelae have not been well characterized during the post-acute phase of SARS-CoV-2 among adults 18 to 65 years old, and this study sought to fill that gap by evaluating excess risk and relative hazards for developing incident clinical sequelae during the post-acute phase.

**Design:** Retrospective cohort study including three propensity-matched groups.

**Setting:** This study merged three data sources from a large United States health plan: a large national administrative claims database, an outpatient lab testing database, and an inpatient hospital admissions database.

**Participants:** Individuals 18 to 65 years old with continuous health plan enrollment from January 2019 to date of SARS-CoV-2 diagnosis. Three comparator groups were identified and propensity-score matched to individuals infected with SARS-CoV-2: a 2020 comparator group, a historical 2019 comparator group and a historical comparator group with viral lower respiratory tract illness (vLRTI).

**Main outcome measures:** Over 50 clinical sequelae during the post-acute phase (index date + 21 days) were ascertained using ICD-10 codes. Excess risk due to SARS-CoV-2 during the 4 months following the acute phase of illness and hazard ratios with 95% Bonferroni-corrected confidence intervals were calculated.

**Results:** This study found 14% of adults ≤65 years of age who were infected with SARS-CoV-2 (n=193113) had at least one new clinical sequelae that required medical attention during the post-acute phase of illness. When considering risk for specific sequelae attributable to SARS-Cov-2 infection during the post-acute phase, clinical outcomes including chronic respiratory failure, cardiac arrythmia, hypercoagulability, encephalopathy, peripheral neuropathy, amnesia (memory difficulty), diabetes, liver test abnormalities, myocarditis, anxiety and fatigue were significantly elevated compared to the three propensity-matched comparator groups (2020, 2019, vLRTI). Significant risk differences due to SARS-CoV-2 infection ranged from 0.02 to 2.26 per 100 people and hazard ratios ranged from 1.24 to 25.65 when compared to the 2020 comparator group.

**Conclusions:** Our results confirm excess risk for developing clinical sequelae due to SARS-CoV-2 during the post-acute phase, including specific types of sequelae less commonly seen among other viral illnesses. Although individuals who were older, had pre-existing conditions, and were hospitalized due to COVID-19 were at greatest excess risk, younger adults (≤50 years), adults who did not have pre-existing conditions or adults who were not hospitalized due to COVID-19 were still at elevated risk for developing new clinical sequelae. The elevated risk for incident sequelae during the post-acute phase is relevant for healthcare planning.

**Summary Box:** *What is already known on this topic:* Small observational studies and case reports of hospitalized patients have shown some COVID-19 survivors suffer from short- and long-term sequelae. Few studies have characterized the excess risk of clinical sequelae attributable to SARS-CoV-2 during the post-acute phase among adults ≤65 years of age in a large generalizable sample.

*What this study adds:* This study found 14% of individuals ≤65 years of age who were infected with SARS-CoV-2 (n=193113) had a diagnosis of at least one new sequelae that required medical attention during the post-acute phase of illness. Elevated risk for specific clinical sequelae during the post-acute phase of illness was noted across a range of organ systems including cardiovascular, neurologic, kidney, respiratory, and mental health complications. The risk for incident sequelae increases with age, pre-existing conditions, and hospitalization for COVID-19; however, even among adults ≤ 50 years of age and individuals without pre-existing conditions or hospitalization due to COVID-19, risk for some clinical sequelae is still elevated. These results indicate where additional diagnostic follow-up, rehabilitation, and symptom management may be warranted among younger adults with milder infection.

## Introduction

Emerging data suggest sequelae of infection with SARS-CoV-2 and the disease it causes, COVID-19, may vary in presentation and extend beyond the typical post-viral recovery period. As such, epidemiologic interest in post-acute morbidity among survivors is growing. Some survivors experience serious complications during the acute phase of the illness affecting pulmonary, cardiovascular, hepatic, renal, cognitive, and neurologic function.(1–4) Survivors also report a range of persistent symptoms adversely impacting physical, mental, and social well-being.(5–8) At least some of these complications may occur independent of COVID-19 severity.(9) While individuals hospitalized with community-acquired (non-COVID-19) pneumonia or influenza face risk of cardiovascular, cerebrovascular, and neurological complications,(10,11) the degree of increased risk resulting from SARS-CoV-2 infection remains unclear. Longitudinal studies on survivors from other coronaviruses (MERS and SARS) suggest long-term physical and mental sequelae are not uncommon.(12–14)

Most published studies to date have been small and primarily focused on clinical sequelae among hospitalized patients.(15) As a result, many of these studies may not be generalizable to the larger population of SARS-CoV-2 infected individuals. Little is known about the incidence of clinical sequelae due to SARS-CoV-2 infection during the post-acute phase of illness among adults 18 to 65 years old who may be considered at lower risk for severe COVID-19. Moreover, few studies have been powered to evaluate whether factors such as age, gender, pre-existing conditions, and hospitalization status modify risk of clinical sequelae in the post-acute phase of illness.

We have estimated the excess risk and hazard ratios of new clinical sequelae attributable to SARS-CoV-2 among adults 18 to 65 years old during the post-acute phase of the SARS-CoV-2 infection. This analysis includes a large generalizable sample of commercially insured adults, has ascertained objective outcomes utilizing valid International Classification of Diseases Tenth Revision (ICD-10) codes in claims, and has the power to both detect rare diagnoses and evaluate associations across subgroups.

## Methods

### Data Source

We conducted a retrospective cohort analysis utilizing three data sources within the UnitedHealth Group Clinical Discovery Database: de-identified administrative outpatient and inpatient claims, outpatient lab results for SARS-CoV-2, and a hospital admissions database updated daily with patients admitted with primary, secondary, or tertiary diagnosis of COVID-19. Quality control efforts for data were applied (eAppendix A). All data were for commercially enrolled patients from a single large national health insurance provider in the United States. This research was determined to be exempt from human research regulations by the UnitedHealth Group Office of Human Research Affairs (Action ID: 2020-0076-01).

### Study Population

#### Individuals Diagnosed with SARS-CoV-2

This group consisted of individuals 18 to 65 years old with continuous health plan enrollment from January 1, 2019 to first date of one of the following, which was denoted as the index date:

1. primary, secondary, or tertiary diagnosis of COVID-19 identified by administrative claims with ICD-10 code U07.1 or either B34.2 or B97.29 before April 1, 2020 **OR**
2. documentation of a positive PCR test within an outpatient lab dataset **OR**
3. hospitalization due to COVID-19 identified from a hospital admissions database with admitting diagnosis code or primary, secondary, or tertiary diagnosis code U07.1 or U07.2.

We excluded individuals with a positive SARS-CoV-2 antibody serology without documented infection (n= 28,810) because index dating of illness was not possible. We also excluded individuals with a diagnosis code of B34.2 or B97.29 on or after April 1, 2020 (n=24,865) and individuals hospitalized due to suspected COVID-19 but missing diagnosis code U07.1 or U07.2 in primary, secondary, or tertiary position (n=1,247). A flow chart with details of population sampling is provided in Figure 1.

**Figure 1:**
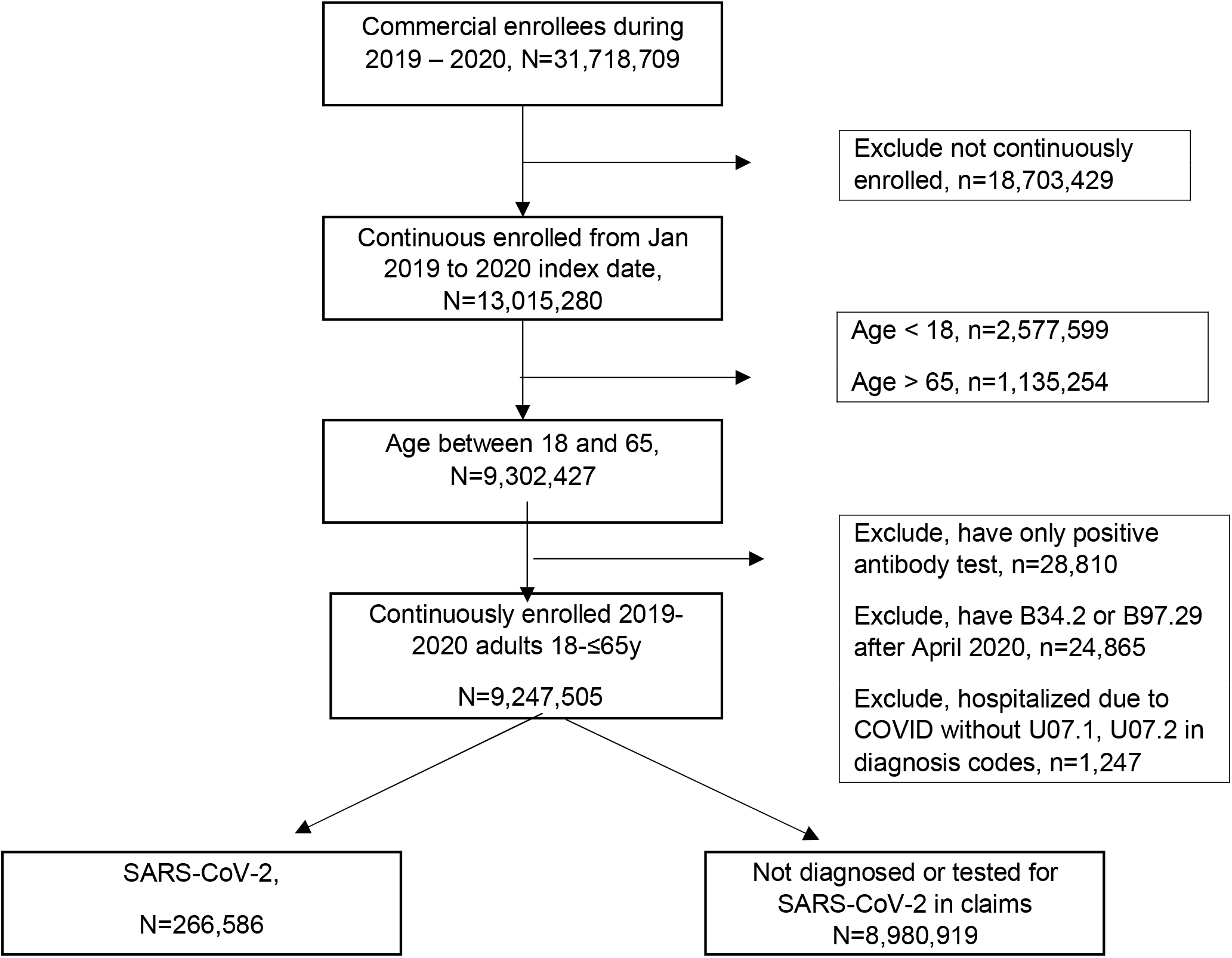
2020 Cohort Sample.

### 2020 Comparator Group

The 2020 comparator group consisted of individuals 18 to 65 years old without a COVID-19-related clinical diagnosis, positive PCR test, or hospitalization due to COVID-19 in 2020. Continuous health plan enrollment was required from January 1, 2019 to a randomly assigned index date drawn from the SARS-CoV-2 infection group.

### 2019 Comparator Group

We created this historical comparison group to account for possible ascertainment bias due to lower utilization of healthcare services during the 2020 pandemic. Individuals 18 to 65 years old were required to have continuous health plan enrollment from January 1, 2018 to a randomly assigned month and day in 2019 drawn from the SARS-CoV-2 infection group.

### Viral Lower Respiratory Tract Illness (vLRTI) Comparator Group

We created this historical comparison group to evaluate the clinical sequelae specific to SARS-CoV-2 infection, as many serious viral illnesses carry risk for developing post-acute illness morbidity. The vLRTI group included individuals 18 to 65 years old with a diagnosis of any one of the following: influenza (J09, J10, J11), non-bacterial pneumonia (J12, J18.9), acute bronchitis (J20), acute lower respiratory infection (J22), and COPD with acute lower respiratory infection (J44.0) between January 1, 2017 and October 31, 2017, January 1, 2018 and October 31, 2018, or January 1, 2019 and October 31,2019. We included COPD exacerbation since cases are typically virally induced and identified only with a J44 code. We defined the index date as the first date of diagnosis for vLRTI. Individuals were required to have continuous enrollment from January 1, 2016, January 1, 2017, or January 1, 2018 to the index date, respectively.

### Main Outcomes

We utilized ICD-10 codes to identify new clinical diagnoses from the administrative claims data between January 1, 2020 and October 31, 2020. See eTable 1 for ICD-10 classification details. We created domain clusters based on clinically similar diagnoses, and we included atopic dermatitis as a negative control.(16)

### Study Variables

We used administrative claims between January 1, 2019 and 30 days prior to index date to ascertain length of hospital stay, prior clinical conditions, history of comorbidities derived from the Charlson Index and Elixhauser score, and prior PCP and specialist visits. Demographic, clinical, and testing data were obtained from administrative claims between January 1, 2020 and October 31, 2020. We derived zip code-specific socioeconomic status scores (SES) and proportions of White, African American and Hispanic populations by zip code.(17) Missing values of zip-code derived variables (SES n=11,713; race n= 444,080) were imputed with the median of non-missing values for these variables. Since all other variables were derived from administrative claims and the continuous enrollment criteria were applied, clinical events without a claim were considered to have not occurred and were given a zero value.

### Follow-Up

Follow-up periods for the primary analysis for SARS-CoV-2 infected individuals and comparator groups started at index date + 21 days and continued through to a diagnostic event, disenrollment from insurance plan (due to death or withdrawal), or end of study period (October 31, 2020 or October 31, corresponding year for the historical comparator groups), whichever occurred first. We performed a secondary analysis evaluating rates by month of follow-up starting 30 days prior to index date and ending at 6 months follow-up.

### Public and Patient Involvement

This retrospective analysis did not directly involve patients or the public in the development of the research question or conduct of the analysis. A patient reviewer provided insightful comments and contributed to the expansion of the outcomes reported in this manuscript. The content of this paper will be summarized and made publicly available on the United In Research (unitedinresearch.com) platform sponsored by OptumLabs.

## Statistical Methods

### Propensity-score matching

We used propensity-score matching to create three cohorts similar on baseline characteristics and relevant confounders.(18) We constructed a propensity score for every individual based on 108 variables (age, numeric; gender, male Y/N; zip code-specific SES, numeric; proportion of White, African American and Hispanic populations in zip code, numeric; state, 10 binary variables in top 10 states; index month, Jan-Mar, Apr, May, Jun, Jul, Aug, Sept Y/N; pre-existing comorbidities, Y/N; total inpatient length of stay in the previous year, numeric in days; prior PCP, cardiologist, and nephrologist visit day count, numeric; and prior clinical conditions, Y/N) using logistic regression with ridge penalty.(19,20)

Due to the large study population (about 10 million), we did not perform conventional 1-1 nearest neighbor matching. We generated 40,000 bins based on the SARS-CoV-2 infected group’s propensity scores. In each small bin, we created equal-size sets of individuals with SARS-CoV-2 and the comparator group. We matched within the bins and then combined all sets to form the propensity-matched groups. This process was repeated for each of the comparator groups (2020, 2019, vLRTI). We achieved three balanced cohort groups with this approach, as shown in eTable 1a and eFigure 1a-c.

### Data Analysis

We evaluated demographic and clinical factors using the t-test and Pearson chi-square test for the unmatched population to compare numeric and categorical data, respectively. We calculated the proportion of individuals with follow-up time after 21 days post-index date who were diagnosed with no clinical sequelae, one, or more than one clinical sequelae during the post-acute period. Symptoms such as fatigue, myalgia, and anosmia as measured by ICD-10 codes were included in this post-acute assessment of clinical sequelae.

When calculating the risk for a specific incident outcome in the post-acute phase, individuals who had the diagnosis of interest prior to index date (in the SARS-CoV-2 and respective comparator group) or individuals (and their comparator match) who experienced the diagnosis of interest during the acute phase were removed from the post-acute calculation. Therefore, the denominator for the incidence of each outcome included only those at risk for the diagnosis of interest at index date+21 days. We reported the risk difference as the difference between the cumulative incidence calculated by the Kaplan-Meier estimator at the 120^th^ day from time origin (index date + 21 days) in the SARS-CoV-2 group and the comparator group, multiplied by 100. We calculated the 95% confidence intervals and p values using a pairwise bootstrap method, with a one-sided test We tested the proportional hazards assumption using Schoenfeld residuals.(21) Hazard ratios and 95% confidence intervals were estimated by fitting Cox proportional hazards models using robust variance estimator with clustering for matched pairs. We used the Wald-test to evaluate significance at the 0.05 level. Individuals were censored based on the outcome of interest, disenrollment from insurance plan, or end of the study period (October 31, 2020).

In a secondary analysis, we created seven 1-month time intervals (−1 month prior to index date to 6 months after index date). Within each time interval, we only included the matched pairs who are both at-risk for the diagnosis of interest at the beginning of each time interval. We estimated the hazard ratios and confidence intervals by fitting Cox proportional hazard models using robust variance estimator with clustering for matched pairs. Individuals were censored based on diagnosis of interest, disenrollment, or end of the time interval.

Due to the large number of comparisons, we applied the Bonferroni correction for all p values and confidence intervals, by multiplying p values by *N* and estimating 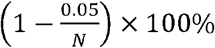 confidence intervals, where *N*= 51 and is the number of clinical sequelae we tested.

For all data analysis, we used Python with scikit-learn, statsmodels, lifelines, and scipy libraries and R with survminer,(22) survival,(23) glmnet,(24) and stats libraries.(25)

### Subgroup Analysis

We performed four stratified analyses [age (18-≤34, >34-≤50 >50), gender (M/F), any pre-existing clinical comorbidity (Y/N), and hospitalization due to COVID-19 (Y/N)] with the SARS-CoV-2 and 2020 comparator group. We constructed a propensity score and performed propensity score matching within each stratum before calculating the risk difference or hazard ratio. To test for interaction, we evaluated the significance of the fitted coefficient for the interaction term in a model including main effect variables, with a Bonferroni correction (*N*=51*number of levels in each subgroup analysis).

### Sensitivity Analysis

We evaluated the robustness of the post-acute phase definition by varying the cut points from index date at 14 and 28 days. We also considered a period effect due to changes in the availability of testing and advancements in management and treatment over time. We evaluated differences by two periods (January–June; July–October). Finally, we evaluated medical utilization during the post-acute phase by type of diagnostic method (PCR-positive, clinical diagnosis, hospitalization).

## Results

Among 9,247,505 individuals meeting the study criteria in 2020, we identified 266,586 individuals (2.9%) with SARS-CoV-2 infection. After propensity score matching, we identified 266,586 matched pairs for the primary (2020) and secondary (2019) comparison group (100% of SARS-CoV-2 individuals matched). We identified 244,276 matched pairs for the vLRTI comparison group (91.6% of SARS-CoV-2 individuals matched).

Individuals with SARS-CoV-2 infection were more likely than their unmatched 2020 and 2019 comparators to be female, have lower SES index, live in a zip code with majority black or Hispanic individuals, have a pre-existing comorbidity or at-risk condition, have an in-patient hospital stay during the preceding year, visit a PCP and other specialists more often, and live in the Northeast or Southern U.S (areas of high incidence during the study period; all p-values<0.05) (Table 1). Among those individuals with SARS-CoV-2, 8.2% were hospitalized and 1.1% were admitted to the ICU. We observed different patterns between the SARS-CoV-2 individuals and the vLRTI comparison group who were more likely to be older, female, have a comorbidity, smoke, visit a PCP or cardiologist, and live in the South. Following propensity score matching, most of these differences were resolved, although some small differences between the matched SARS-CoV-2 infected group and the vLRTI comparator remained significant (eTable 1a). Despite these minor differences, balance was achieved overall as shown by the standardized mean differences between matched groups by key variables at less than 10% (eFigure 1a-c).

Among the matched SARS-CoV-2 infected individuals identified in our study with follow-up time past 21 days from index date (n=193113), 85.98% did not have any new post-acute clinical sequelae that required medical attention during their follow-up, 10.01% had one new sequelae that required medical attention, and 4.01% had more than one type of new sequelae (Table 2). The proportion of individuals diagnosed with any new clinical sequelae during the post-acute phase was higher among the SARS-CoV-2 infected group than the three comparator groups, although the differences were smaller when compared to the vLRTI comparator group.

**Table 1:** Demographics, Comorbidities, and Clinical Factors Among Unmatched Adults 18 to ≤65y, UnitedHealth Group Clinical Discovery Database through October 31, 2020.

| Characteristic | Total 2020 Population | 2020 SARS-CoV-2 Infected n(%) | 2020 Comparator n(%)* | 2019 Comparator n(%)* | Viral LRTI n(%)* |
| --- | --- | --- | --- | --- | --- |
| **Total n** | 9247505 | 266586 | 8980919 | 9722381 | 1655907 |
| **Follow-up from index date in days: Median (IQR)** |  | 95 (42 - 135) |  |  | 226 (136 - 268) * |
| **age: Mean (SD)** | 42.4 (13.6) | 41.7 (13.9) | 42.4 (13.6) * | 42.5 (13.6) * | 44.5 (13.4) * |
| >17 - ≤34 | 2903571 (31.4) | 90497 (33.9) | 2813074 (31.3) | 3038968 (31.3) | 417742 (25.2) |
| >34 - ≤49 | 3035730 (32.8) | 82497 (30.9) | 2953233 (32.9) | 3199622 (32.9) | 547531 (33.1) |
| > 49 | 3308204 (35.8) | 93592 (35.1) | 3214612 (35.8) | 3483791 (35.8) | 690634 (41.7) |
| **gender** |  |  |  |  |  |
| Male | 4640393 (50.2) | 126980 (47.6) | 4513413 (50.3) * | 4861274 (50.0) * | 707084 (42.7) * |
| **ses_index: Median (IQR)** | 53.2 (51.6 - 55.2) | 52.9 (51.2 - 55.1) | 53.2 (51.6 - 55.2) * | 53.1 (51.6 - 55.2) * | 52.9 (51.2 - 55.2) |
| **White proportion: Mean (SD)** | 0.66 (0.24) | 0.60 (0.26) | 0.66 (0.24) * | 0.66 (0.24) * | 0.66 (0.24) * |
| <= 0.25 | 794354 (8.6) | 36954 (13.9) | 757400 (8.4) | 864308 (8.9) | 134213 (8.1) |
| >0.25- ≤0.50 | 1342855 (14.5) | 48299 (18.1) | 1294556 (14.4) | 1432247 (14.7) | 246827 (14.9) |
| >0.50 -≤0.75 | 3156844 (34.1) | 85969 (32.2) | 3070875 (34.2) | 3214324 (33.1) | 540297 (32.6) |
| > 0.75 | 3953452 (42.8) | 95364 (35.8) | 3858088 (43.0) | 4211502 (43.3) | 734570 (44.4) |
| **Black proportion: Mean (SD)** | 0.10 (0.15) | 0.13 (0.17) | 0.10 (0.15) * | 0.10 (0.15) * | 0.11 (0.15) * |
| <= 0.25 | 8218593 (88.9) | 226443 (84.9) | 7992150 (89.0) | 8618587 (88.6) | 1462821 (88.3) |
| >0.25- ≤0.50 | 666921 (7.2) | 25327 (9.5) | 641594 (7.1) | 712056 (7.3) | 131297 (7.9) |
| >0.50 -≤0.75 | 252382 (2.7) | 10241 (3.8) | 242141 (2.7) | 272726 (2.8) | 44280 (2.7) |
| > 0.75 | 109609 (1.2) | 4575 (1.7) | 105034 (1.2) | 119012 (1.2) | 17509 (1.1) |
| **Hispanic proportion: Mean (SD)** | 0.15 (0.17) | 0.19 (0.21) | 0.14 (0.17) * | 0.15 (0.17) * | 0.15 (0.17) * |
| <= 0.25 | 7705168 (83.3) | 201097 (75.4) | 7504071 (83.6) | 8053655 (82.8) | 1359968 (82.1) |
| >0.25- ≤0.50 | 1040572 (11.3) | 39727 (14.9) | 1000845 (11.1) | 1120296 (11.5) | 202486 (12.2) |
| >0.50 -≤0.75 | 352428 (3.8) | 16934 (6.4) | 335494 (3.7) | 380593 (3.9) | 64624 (3.9) |
| > 0.75 | 149337 (1.6) | 8828 (3.3) | 140509 (1.6) | 167837 (1.7) | 28829 (1.7) |
| **Index month** |  |  |  |  |  |
| Prior to March |  | 200 (0.1) |  |  | 777212 (46.9) * |
| March |  | 11116 (4.2) |  |  | 249299 (15.1) * |
| April |  | 23096 (8.7) |  |  | 146844 (8.9) * |
| May |  | 22183 (8.3) |  |  | 107928 (6.5) * |
| June |  | 36143 (13.6) |  |  | 78324 (4.7) * |
| July |  | 55274 (20.7) |  |  | 58609 (3.5) * |
| August |  | 35692 (13.4) |  |  | 60584 (3.7) * |
| September |  | 33555 (12.6) |  |  | 80926 (4.9) * |
| October |  | 49327 (18.5) |  |  | 96181 (5.8) * |
| **Comorbidity** |  |  |  |  |  |
| Any | 3311964 (35.8) | 120749 (45.3) | 3191215 (35.5) * | 3584107 (36.9) * | 996015 (60.1) * |
| AIDS/HIV | 17115 (0.2) | 992 (0.4) | 16123 (0.2) * | 18833 (0.2) * | 5234 (0.3) * |
| Alcohol Abuse | 74314 (0.8) | 2959 (1.1) | 71355 (0.8) * | 80222 (0.8) * | 19076 (1.2) |
| Anemia | 322983 (3.5) | 16157 (6.1) | 306826 (3.4) * | 360339 (3.7) * | 107686 (6.5) * |
| Rheumatoid Arthritis | 162750 (1.8) | 6793 (2.5) | 155957 (1.7) * | 181286 (1.9) * | 65297 (3.9) * |
| Cerebrovascular Disease | 89248 (1.0) | 4342 (1.6) | 84906 (0.9) * | 102768 (1.1) * | 34446 (2.1) * |
| Congestive Heart Failure | 61783 (0.7) | 3362 (1.3) | 58421 (0.7) * | 68361 (0.7) * | 29175 (1.8) * |
| Coagulopathy | 46482 (0.5) | 2193 (0.8) | 44289 (0.5) * | 53344 (0.5) * | 17711 (1.1) * |
| Chronic Pulmonary Disease | 523530 (5.7) | 22236 (8.3) | 501294 (5.6) * | 580623 (6.0) * | 252266 (15.2) * |
| Dementia | 3500 (0.0) | 205 (0.1) | 3295 (0.0) * | 4154 (0.0) * | 1524 (0.1) * |
| Depression | 553657 (6.0) | 19656 (7.4) | 534001 (5.9) * | 572283 (5.9) * | 164835 (10.0) * |
| Drug Abuse | 65075 (0.7) | 2485 (0.9) | 62590 (0.7) * | 71788 (0.7) * | 21781 (1.3) * |
| Hypertension | 1345161 (14.5) | 51908 (19.5) | 1293253 (14.4) * | 1473333 (15.2) * | 443477 (26.8) * |
| Hypothyroidism | 481219 (5.2) | 17335 (6.5) | 463884 (5.2) * | 533528 (5.5) * | 166571 (10.1) * |
| Liver Disease | 191592 (2.1) | 9017 (3.4) | 182575 (2.0) * | 208626 (2.1) * | 65363 (3.9) * |
| Moderate to Severe Liver Disease | 5996 (0.1) | 379 (0.1) | 5617 (0.1) * | 6625 (0.1) * | 2854 (0.2) * |
| Lymphoma | 15740 (0.2) | 802 (0.3) | 14938 (0.2) * | 17541 (0.2) * | 7965 (0.5) * |
| Fluid and Electrolyte Disorders | 168283 (1.8) | 8493 (3.2) | 159790 (1.8) * | 189208 (1.9) * | 65360 (3.9) * |
| Metastatic Cancer | 25341 (0.3) | 1415 (0.5) | 23926 (0.3) * | 27580 (0.3) * | 11766 (0.7) * |
| Myocardial Infarction | 32990 (0.4) | 1499 (0.6) | 31491 (0.4) * | 36238 (0.4) * | 12307 (0.7) * |
| Other Neurological Disorders | 160409 (1.7) | 6514 (2.4) | 153895 (1.7) * | 175054 (1.8) * | 56933 (3.4) * |
| Obesity | 835125 (9.0) | 36149 (13.6) | 798976 (8.9) * | 907130 (9.3) * | 249398 (15.1) * |
| Paralysis | 16713 (0.2) | 774 (0.3) | 15939 (0.2) * | 18579 (0.2) * | 6829 (0.4) * |
| Peripheral Vascular Disease | 94789 (1.0) | 4861 (1.8) | 89928 (1.0) * | 107517 (1.1) * | 38322 (2.3) * |
| Psychoses | 145773 (1.6) | 4878 (1.8) | 140895 (1.6) * | 151628 (1.6) * | 43743 (2.6) * |
| Peptic Ulcer Disease | 22179 (0.2) | 1168 (0.4) | 21011 (0.2) * | 26009 (0.3) * | 9354 (0.6) * |
| Pulmonary Circulation Disorder | 19038 (0.2) | 1022 (0.4) | 18016 (0.2) * | 21401 (0.2) * | 9921 (0.6) * |
| Renal Failure | 77877 (0.8) | 3893 (1.5) | 73984 (0.8) * | 82456 (0.8) * | 29103 (1.8) * |
| Solid Tumor without Metastasis | 175848 (1.9) | 6824 (2.6) | 169024 (1.9) * | 192765 (2.0) * | 61012 (3.7) * |
| Valvular Disease | 136937 (1.5) | 6331 (2.4) | 130606 (1.5) * | 154148 (1.6) * | 48726 (2.9) * |
| Weight Loss | 65384 (0.7) | 2909 (1.1) | 62475 (0.7) * | 72404 (0.7) * | 20963 (1.3) * |
| **Prior Conditions** |  |  |  |  |  |
| Alzheimer Dementia | 6760 (0.1) | 385 (0.1) | 6375 (0.1) * | 8283 (0.1) * | 3203 (0.2) * |
| Asthma | 355183 (3.8) | 15112 (5.7) | 340071 (3.8) * | 382552 (3.9) * | 162684 (9.8) * |
| Cystic Fibrosis | 1284 (0.0) | 64 (0.0) | 1220 (0.0) * | 1349 (0.0) * | 818 (0.0) * |
| Immunodeficiency | 57139 (0.6) | 3101 (1.2) | 54038 (0.6) * | 60545 (0.6) * | 22040 (1.3) * |
| Pulmonary Fibrosis | 9436 (0.1) | 488 (0.2) | 8948 (0.1) * | 10566 (0.1) * | 5444 (0.3) * |
| Sickle Cell Disease | 4008 (0.0) | 273 (0.1) | 3735 (0.0) * | 4258 (0.0) * | 1211 (0.1) * |
| Smoking | 289418 (3.1) | 8113 (3.0) | 281305 (3.1) * | 321713 (3.3) * | 104321 (6.3) * |
| T1D | 46947 (0.5) | 2157 (0.8) | 44790 (0.5) * | 52806 (0.5) * | 18342 (1.1) * |
| T2D | 474752 (5.1) | 22619 (8.5) | 452133 (5.0) * | 521746 (5.4) * | 171703 (10.4) * |
| Thalassemia | 6055 (0.1) | 226 (0.1) | 5829 (0.1) * | 6467 (0.1) * | 1358 (0.1) |
| **Prior Length of Stay in Hospital (days): Mean (SD)** | 0.3 (2.9) | 0.5 (5.9) | 0.3 (2.7) * | 0.3 (2.9) * | 0.5 (5.1) |
| **Prior PCP Visit (%)** | 5142739 (55.6) | 186313 (70.0) | 4956426 (55.2) | 5617767 (57.8) | 107686 (75.1) |
| **Count of PCP visit: Mean (SD)** | 1.7 (2.7) | 2.5 (3.5) | 1.7 (2.6) | 1.8 (2.8) | 2.9 (3.7) |
| **Count of cardiology visit: Mean (SD)** | 0.2 (1.0) | 0.3 (1.7) | 0.2 (1.0) | 0.2 (1.1) | 0.4 (1.7) |
| **Count of nephrology visit: Mean (SD)** | 0.0 (0.7) | 0.1 (1.6) | 0.0 (0.6) | 0.0 (0.7) | 0.1 (1.5) |
| **Region** |  |  |  |  |  |
| South | 3924142 (42.4) | 117292 (44.0) | 3806850 (42.4) * | 4039716 (41.6) * | 799967 (48.3) * |
| Midwest | 2171722 (23.5) | 54494 (20.4) | 2117228 (23.6) * | 2312541 (23.8) * | 363017 (21.9) * |
| Northeast | 1590979 (17.2) | 56071 (21.0) | 1534908 (17.1) * | 1714661 (17.6) * | 281385 (17.0) * |
| West | 1538866 (16.6) | 38300 (14.4) | 1500566 (16.7) * | 1631945 (16.8) * | 208353 (12.6) * |
| Unknown | 21796 (0.2) | 429 (0.2) | 21367 (0.2) * | 23518 (0.2) * | 3185 (0.2) * |
| **Clinical characteristics** |  |  |  |  |  |
| COVID- ICU visit flag |  | 2786 (1.1) |  |  |  |
| Hospitalization status |  | 21746 (8.2) |  |  |  |
| **Diagnostic method+** |  |  |  |  |  |
| PCR test positive |  | 77273 (29.0) |  |  |  |
| Clinical Diagnosis (not PCR test) |  | 178642 (67.0) |  |  |  |
| Hospitalization (not PCR or DX) |  | 10671 (4.0) |  |  |  |

\*all p-values $\leq 0.05$ comparing SARS-CoV-2 individuals with 3 comparison groups
+ presented as hierarchical in table, although many individuals had more than one diagnostic method recorded

**Table 2:** Proportion of Adults 18 to ≤65 Years of Age with Post-Acute Sequelae, UnitedHealth Group Clinical Discovery Database through October 31, 2020□□.

| **Sequelae**<br>*☐☐☐ | **SARS-Cov-2**<br>**Infected (matched to 2020)**<br>(N=193113)☐☐ | **2020**<br>**Comparator**<br>**(matched)**☐☐<br>(N=193113)☐☐ | **SARS-Cov-2**<br>**Infected (matched with 2019)**<br>(N=193264)☐☐ | **2019 Comparator**<br>**(matched)**<br>(N=193264)☐☐ | **SARS-Cov-2**<br>**Infected (matched with vLRTI)**<br>(N=181613)☐☐ | **vLRTI**<br>**Comparator (matched)**<br>(N=181613)☐☐ |
| --- | --- | --- | --- | --- | --- | --- |
| **None**☐☐☐ | 166039/193113<br>(85.98%)☐☐ | 175593/193113<br>(90.93%)☐☐ | 166094/193264<br>(85.94%)☐☐ | 174963/193264<br>(90.53%)☐☐ | 155083/181613<br>(85.39%)☐☐ | 158078/181613<br>(87.04%)☐☐ |
| **1**☐☐☐ | 19328/193113<br>(10.01%)☐☐ | 13671/193113<br>(7.08%)☐☐ | 19408/193264<br>(10.04%)☐☐ | 14224/193264<br>(7.36%)☐☐ | 18858/181613<br>(10.38%)☐☐ | 17627/181613<br>(9.71%)☐☐ |
| **2+**☐☐☐ | 7746/193113<br>(4.01%)☐☐ | 3849/193113<br>(1.99%)☐☐ | 7762/193264<br>(4.02%)☐☐ | 4077/193264<br>(2.11%)☐☐ | 7672/181613<br>(4.22%)☐☐ | 5908/181613<br>(3.25%)☐☐ |

\* sequelae ☐ are new clinical diagnoses ascertained after index date + 21 ☐ days ☐
† McNemar test was performed and all p-values <0.01

Estimates of risk difference by type of new clinical sequelae were calculated among individuals infected with SARS-CoV-2 who were still at-risk starting at 21 days post-index date (n=193113 for 2020 comparison). Follow-up time was considered through day 141 after index date (120 days from the start of post-acute period and 73.5 percentile of the follow-up distribution, with median = 87 days [IQR 45– 124 days]). The most common new clinical outcomes among the SARS-Cov-2 group (incidence ≥0.1%) are summarized in Figure 2, with less common outcomes in eTable 2a. Symptoms are included separately in eTable 2b and were not represented in Figure 2. Overall, the excess risk attributable to SARS-CoV-2 infection is low for incident diagnoses (0.02%–2.26%) 4 months from the start of the post-acute phase (Figure 2). The elevated risk, however, is consistently observed for many outcomes across all three comparison groups (2020, 2019, and vLRTI; eTable 2a).

**Figure 2:**
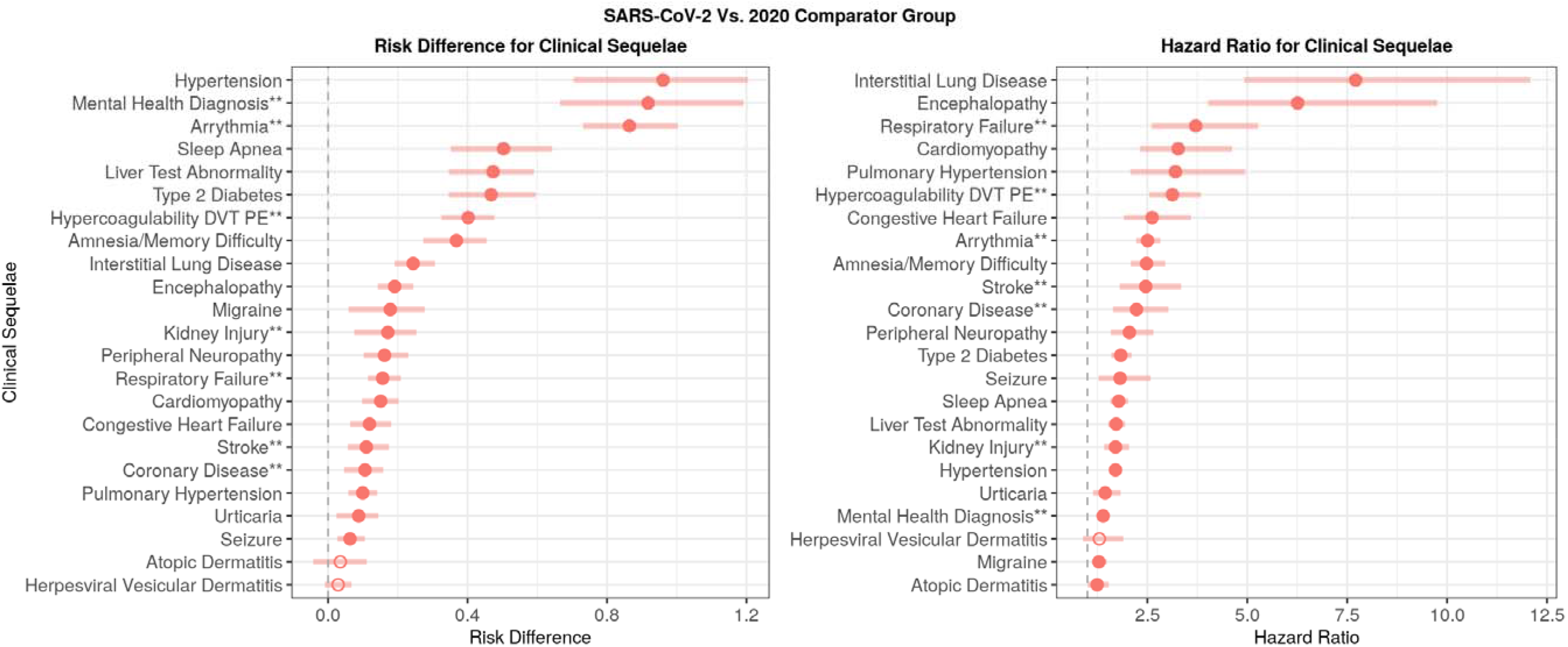
Risk Difference (per 100 individuals; left) and Hazard Ratios (right) for Most Common Clinical Sequelae, UnitedHealth Group Clinical Discovery Database through October 31, 2020.* *Clinical sequelae included in the graphs are diagnoses with incidence ≥0.1 in the SARS-CoV-2 group during the first 4 months of the post-acute phase (index date+21 days) and highest in hierarchy if an aggregate diagnosis. We adopted this rule to avoid confidence intervals that are too wide to display. Symptoms are not displayed in the graphs. All associations for each of the 51 outcomes are listed in Table e2a-2d. *Filled shapes indicate significant risk difference or hazard ratio (Bonferroni corrected p value <0.05). *Atopic dermatitis is present in all plots to present a negative control. ** Aggregate diagnosis includes all subdiagnoses as listed in eTable 1

Despite the small absolute risk attributable to SARS-CoV-2 infection, the HRs between SARS-CoV-2 infected individuals and 2020 comparator group during the post-acute phase can be large (significant HRs between 1.24 to 25.65; eTable 2c-c; Figure 2). When evaluating rates over time, HRs are highest in the first month of index date, but remain significantly elevated through month 6 suggesting the hazard for new clinical sequelae is sustained months after initial SARS-CoV-2 infection (eTable 3). Elevated risk seen 30 days prior to index date was likely due to a delay in testing or documentation of a confirmed case among symptomatic individuals. Select graphs of the cumulative hazards for the most common or most significant clinical sequelae are detailed in Figure 3a-c.

**Figure 3a-d:**
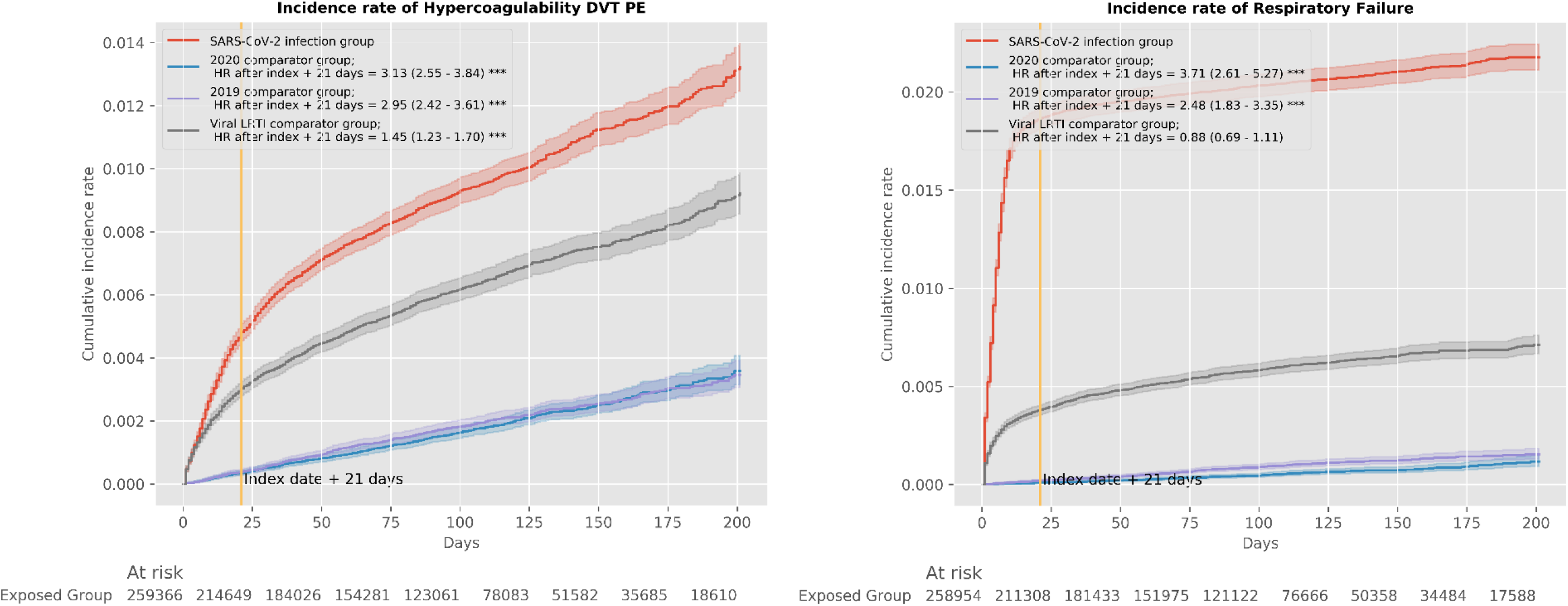
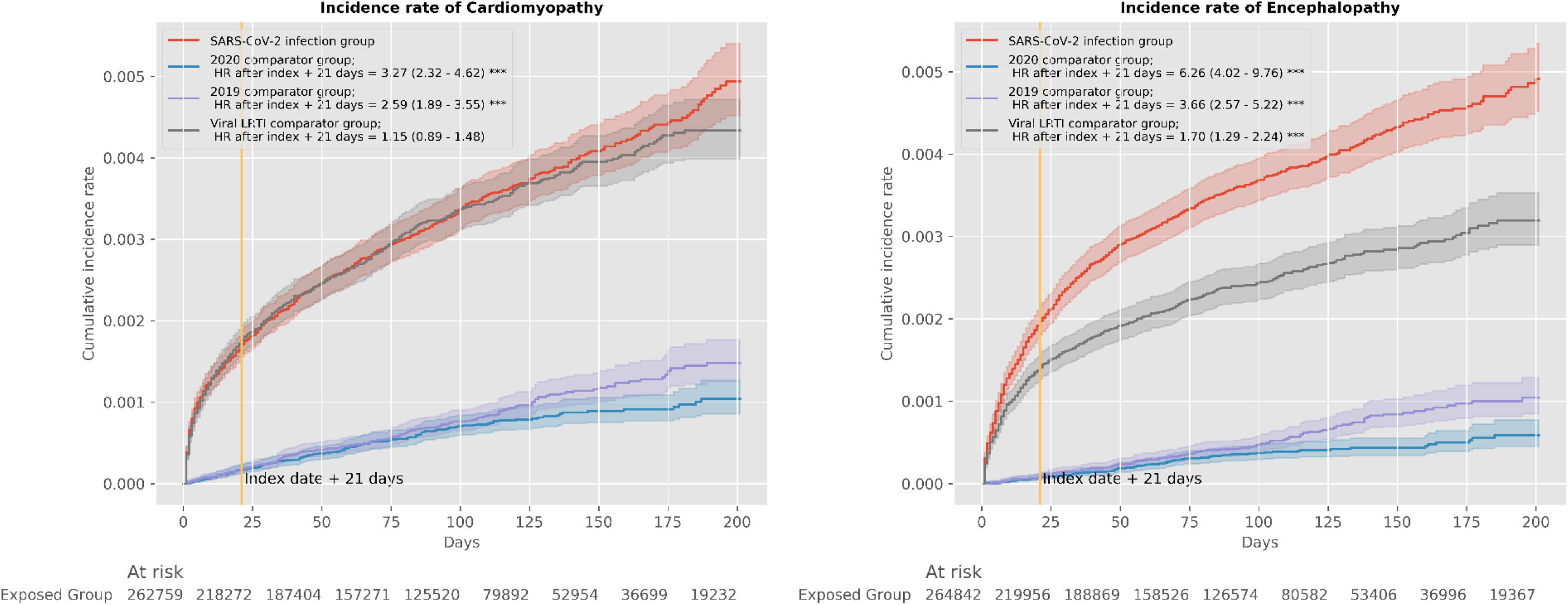
Cumulative Incidence Rate for New Clinical Sequelae and Symptoms after SARS-CoV-2 Infection With 3 Propensity-Matched Groups, UnitedHealth Group Clinical Discovery Database through October 31, 2020.

Excess risk for developing many new outcomes during the post-acute phase increased significantly with age (Figure 4a; eTable 4a). Risk for clinical sequelae was greatest in individuals >50 years old, however, the absolute risk among young adults 18 to 34 years old remained statistically significantly elevated for some conditions, albeit modestly so. Risk for developing any mental health outcome was significantly elevated regardless of age (p_interaction_ =0.35).

**Figure 4a-4d:**
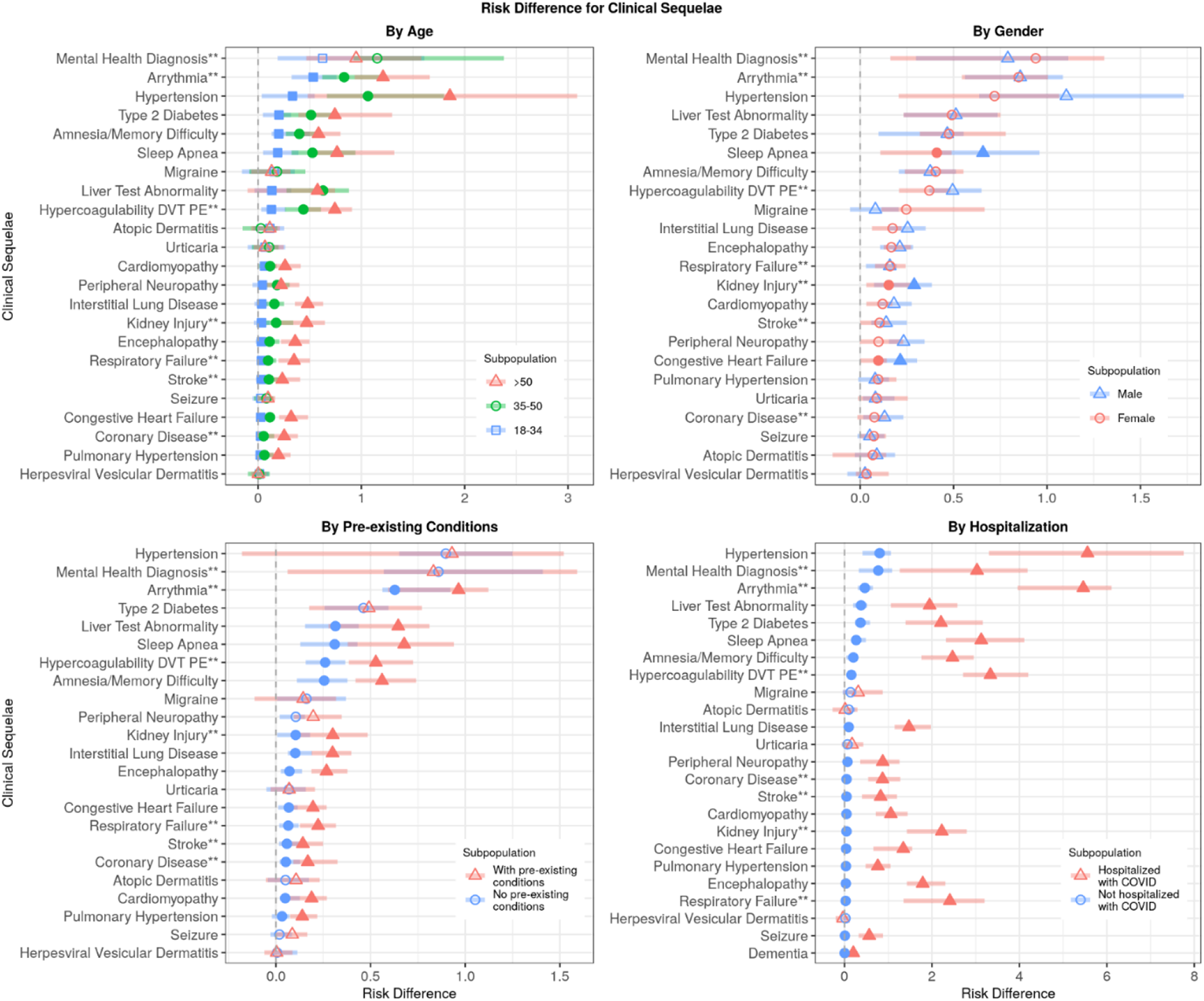
Risk difference (per 100 individuals) for Most Common Clinical Sequelae by Subpopulation, UnitedHealth Group Clinical Discovery Database through October 31, 2020.* *Clinical sequelae included in the graphs are diagnoses with incidence >0.1 in the SARS-CoV-2 group for any subgroup during the first 4 months of the post-acute phase (index date+21 days) and highest in hierarchy if an aggregate diagnosis. We adopted this rule to avoid confidence intervals that are too wide to display. Symptoms are not displayed in the graphs. All associations for each of the 51 outcomes are listed in Table e4a-4d. *Filled shapes indicate significant interaction term (Bonferroni corrected p value <0.05). *Atopic dermatitis is present in all plots to present a negative control. ** Aggregate diagnosis includes all subdiagnoses as listed in eTable 1

Excess risk for new clinical sequelae during the post-acute phase rarely differed by gender, apart from fatigue and anosmia (more commonly diagnosed in women) and myocarditis, hypercoagulability, DVT, kidney injury, and sleep apnea (more commonly diagnosed in men) (Figure 4b; eTable 4b). With a few exceptions, individuals with pre-existing conditions (Figure 4c; eTable 4c) and individuals hospitalized with COVID-19 (Figure 4d; eTable 4d) had greater excess risk for developing new clinical sequelae due to SARS-CoV-2 infection.

We did not see a significant period effect for most outcomes (eTable 4e). In our sensitivity analysis, the RDs increased when the post-acute phase was shortened to index date + 14 days (eTable 5a). Similar RDs were observed for index date + 21 days and index date + 28 days (eTable 5b), suggesting index date + 21 days is a reasonable start to the post-acute phase of illness.

## Discussion

Our retrospective study conducted within a large administrative database evaluated the excess risk of receiving a new diagnosis for a wide range of clinical sequelae during the post-acute phase of SARS-CoV-2 infection among commercially insured adults 18 to 65 years old. We report 14% of individuals ≤65 years of age who were infected with SARS-CoV-2 had a diagnosis of at least one new clinical sequelae that required medical attention during the post-acute phase of the SARS-CoV-2 infection. Our results confirm excess risk for specific types of sequelae during the 4 months following the start of the post-acute phase (index date + 21 days). Our analysis also demonstrates that while risk increases with age, pre-existing conditions, and hospitalization due to COVID-19, younger adults (≤50y), individuals with no pre-existing conditions, and non-hospitalized SARS-CoV-2 infected individuals are still at risk for new clinical sequelae in the post-acute phase. Finally, our results suggest risk for some clinical sequelae such as anxiety are elevated regardless of age and pre-existing condition.

When considering risk attributable to SARS-Cov-2 infection, several clinical sequelae remained elevated among survivors during the post-acute phase regardless of comparison group (2020, 2019, vLRTI). These outcomes included chronic respiratory failure, cardiac irregularities such as tachycardia and arrythmia, hypercoagulability in the form of pulmonary embolism and DVTs, anxiety, encephalopathy, peripheral neuropathy, amnesia, diabetes, liver test abnormalities, myocarditis, and fatigue. Many of these outcomes have been previously reported in case studies or observational studies during the acute phase of COVID-19 (including tachycardia,(26) hypercoagulability,(27) mental health outcomes,(28) encephalopathy,(29,30) diabetes,(31) amnesia(32)). A few studies have also highlighted persistent symptoms or new clinical diagnoses during the post-acute phase,(32–38) although few report on a full range of new clinical diagnoses across multiple organ systems in such a large population. Studies of individuals infected with other coronaviruses have shown arrythmia/tachycardia sequelae in SARS survivors(39,40) and central and peripheral nervous system sequelae among both SARS and MERS survivors.(41) The proportion of SARS-CoV-2 infected individuals with a new diagnosis of encephalopathy (0.23) and peripheral neuropathy (0.31) in our study (encephalopathy RD range 0.09-0.19 and peripheral neuropathy RD range 0.12-0.16) was closer to the higher rates of CNS and PNS sequelae seen with MERS.(41) Anxiety was reported in another study to be the most common mental health sequelae among individuals with COVID-19 diagnosis (HR=1.59–2.62) at 14–90 days post-SARS-CoV-2 infection.(42) Our results on mental health outcomes (anxiety; HRs 1.24–1.54) are similar, though absolute risks were lower in our study population.

We also identified excess risk for clinical sequelae that are not unique to SARS-CoV-2 and are commonly seen with other serious viral infections. Still, the magnitude of relative risk for these incident sequelae (e.g, hypertension, stroke, and kidney injury) are nearly twice what is typically seen in the general population in an ordinary year. Because of the scale of the SARS-CoV-2 pandemic in the U.S., these findings suggest additional healthcare resource planning is needed to address the health complications among survivors.

This is the first study, to our knowledge, to have the power to quantify the small, but non-trivial, risk among younger, healthier adults. Within our study population of adults ≤65, greater than 90% of individuals recovered at home. Younger individuals may experience complications due to COVID-19, especially if they are hospitalized and have pre-existing conditions(43), but few studies have reported new outcomes during the post-acute phase of illness among milder cases.

Fatigue is often the most commonly reported symptom during the post-acute phase, with self-reported estimates on surveys ranging from 13.6%(36) to 77.7%(44) depending on hospitalization status and length of follow-up. While fatigue was also the most common post-acute diagnosis in this cohort (4.64%), our estimates only reflect fatigue that was brought to medical attention and noted by the physician. Most symptoms, when associated with a primary viral illness like COVID-19, are frequently not coded by the clinician as they are presumed to be part of the infectious process. Moreover, many individuals with COVID-19 may not seek medical care for a symptom unless it was unusual in duration or severity. ICD-10 codes have been shown to be valid for many clinical diagnoses (45), but an inaccurate and unreliable method for ascertaining symptoms.(46) Therefore, we intentionally did not evaluate a broad list of symptoms in this study as we anticipate the true incidence of symptoms are not accurately reflected when ascertained by ICD-10 codes and would be best ascertained through patient-reported surveys.

We were also not able to ascertain race or ethnicity at the individual level in commercial claims. Additional research is needed to better understand how race and ethnicity modify risk for long-term clinical sequelae. Moreover, our population required continuous enrollment from January 2019 until index date, so individuals who may have been uninsured were not included in our sample.

We may have misclassified individuals due to the retrospective nature of our study and the inherent limitations of using claims to define variables. First, we relied on a lab results database to identify PCR-positive individuals. SARS-CoV-2 infected individuals may have been misclassified into the 2020 comparator group if they tested positive outside our network and were not symptomatic enough to require medical attention. This misclassification would likely have biased our findings towards the null. Second, due to our continuous enrollment requirement for study eligibility, we captured clinical diagnoses and comorbidities that occurred in the previous year and the year of the index date up until 30 days prior to the index date. It is possible that what we observe to be an incident or new diagnosis in the post-acute phase for a specific condition might be an exacerbation of a pre-existing condition that did not receive medical attention within the 13-to 22-month window created by our continuous enrollment criteria. Moreover, we may have misclassified some individuals in the no pre-existing comorbidities subpopulation, potentially inflating the incidence within that subgroup. This misclassification is likely minimal, however, because most medical conditions and comorbidities require individuals to participate in at least annual check-ups with PCPs or specialists. Third, we may not have captured a comprehensive list of all ICD-10 codes for each outcome, although the most common ones were included. Finally, since SARS-CoV-2 is a novel virus, we anticipate physicians may have underestimated the clinical significance of some outcomes, especially early in the pandemic. Our sensitivity analysis, however, does not support this temporal lack of documentation, as we observed no significant difference in risk by period.

Our small excess risks may be due to increased medical attention following SARS-CoV-2 infection, such that it is the ascertainment (diagnosis) rather than the incidence of a new diagnosis that is triggered by SARS-CoV-2 infection. This ascertainment bias is unlikely to fully explain our results, however, as individuals with vLRTI should receive similar medical attention post-acute illness and yet have fewer visits to PCP than any SARS-CoV-2 infected individual, regardless of diagnostic method (eFigure 2).

This retrospective analysis was strengthened by its large sample size that powered the assessment of multiple and rare outcomes simultaneously overall and across multiple subgroups, utilized valid ICD-10 codes to ascertain outcomes, and included a broad definition of SARS-CoV-2 infection (positive PCR results or clinical diagnosis), producing a more generalizable sample of individuals than has been previously studied.

With close to 70 million cases worldwide and rising, the number of SARS-CoV-2 infected survivors with potential post-COVID sequelae will continue to grow. To optimally manage these patients, it is important to understand the incidence and the natural history of each of these sequelae. Our results provide clinicians a comprehensive understanding of the excess risk for over 50 clinical morbidities across multiple organ systems affecting adults ≤65 years of age during the post-acute phase of SARS-Cov-2. Knowing the magnitude of risk for rare and common clinical sequelae may enhance diagnostic evaluation and management for individuals infected with SARS-CoV-2. Moreover, our results help providers and other key stakeholders anticipate the scale of future health complications as well as improve healthcare resource utilization planning.

## Conclusion

We observed 14% of individuals with SARS-CoV-2 infection diagnosed with a new clinical sequelae that required medical attention during the post-acute phase of illness. Elevated and sustained risk for clinical sequelae during the 4 months following acute illness, particularly but not exclusively among individuals with pre-existing conditions or hospitalized due to COVID-19, was observed. Additional follow-up is needed to determine resolution of risk over time.

## Data Availability

The data are proprietary and are not available for public use but, under certain conditions, may be made available to editors and their approved auditors under a data use agreement to confirm the findings of the current study.

## Transparency Statement

The lead author, Sarah Daugherty, affirms that this manuscript is an honest, accurate, and transparent account of the study being reported; that no important aspects of the study have been omitted; and that any discrepancies from the study as planned (and, if relevant, registered) have been explained.

## Contributors and sources

SED, senior researcher, epidemiologist

YG, director, data scientist

KH, National Medical Director for Clinical Intelligence, Physician

JS, principal data scientist, statistician

MCD, data scientist, data scientist

KGJ, data scientist, data scientist.

ML, Professor, epidemiology

KC, Executive Director Translational Research OptumLabs, clinician

SED, YG, KH, JS, MCD, KGJ, ML, KC played an active role in all aspects of the development of the research, including design, conduct, and interpretation of the data; preparation, review, and approval of the manuscript; and decision to submit the manuscript for publication. A description of the data source is included in Supplemental Appendix A.

## Copyright/license for publication

The Corresponding Author has the right to grant on behalf of all authors and does grant on behalf of all authors, a worldwide <u>licence</u> to the Publishers and its licensees in perpetuity, in all forms, formats and media (whether known now or created in the future), to i) publish, reproduce, distribute, display and store the Contribution, ii) translate the Contribution into other languages, create adaptations, reprints, include within collections and create summaries, extracts and/or, abstracts of the Contribution, iii) create any other derivative work(s) based on the Contribution, iv) to exploit all subsidiary rights in the Contribution, v) the inclusion of electronic links from the Contribution to third party material where-ever it may be located; and, vi) licence any third party to do any or all of the above.

## Contributor and guarantor information

The corresponding author attests that all listed authors meet authorship criteria and that no others meeting the criteria have been omitted. SED and YG serve as joint guarantors who accept full responsibility for the work and/or the conduct of the study, had access to the data, and controlled the decision to publish.

## Acknowledgements

We would like to thank Andrea Rotnitzky and Jamie Robins for reviewing and providing feedback on our methods.

## References

1. Leung TYM, Chan AYL, Chan EW, Chan VKY, Chui CSL, Cowling BJ, et al. Short-and potential long-term adverse health outcomes of COVID-19: a rapid review. Emerg Microbes Infect. 2020 Oct 1;9(1):2190–9.

2. Bonow RO, O’Gara PT, Yancy CW. Cardiology and COVID-19. JAMA. 2020 Sep 22;324(12):1131.

3. Winkelmayer WC, Khairallah P, Charytan DM. Nephrology and COVID-19. JAMA. 2020 Sep 22;324(12):1137.

4. Josephson SA, Kamel H. Neurology and COVID-19. JAMA. 2020 Sep 22;324(12):1139.

5. Sudre CH, Murray B, Varsavsky T, Graham MS, Penfold RS, Bowyer RC, et al. Attributes and predictors of Long-COVID: analysis of COVID cases and their symptoms collected by the Covid Symptoms Study App. medRxiv. 2020 Oct 21;2020.10.19.20214494.

6. Carfì A, Bernabei R, Landi F, for the Gemelli Against COVID-19 Post-Acute Care Study Group. Persistent Symptoms in Patients After Acute COVID-19. JAMA. 2020 Aug 11;324(6):603.

7. McGinty EE, Presskreischer R, Han H, Barry CL. Psychological Distress and Loneliness Reported by US Adults in 2018 and April 2020. JAMA. 2020 Jul 7;324(1):93.

8. Tenforde MW. Symptom Duration and Risk Factors for Delayed Return to Usual Health Among Outpatients with COVID-19 in a Multistate Health Care Systems Network — United States, March– June 2020. MMWR Morb Mortal Wkly Rep [Internet]. 2020 [cited 2020 Nov 30];69. Available from: https://www.cdc.gov/mmwr/volumes/69/wr/mm6930e1.htm

9. van den Borst B, Peters JB, Brink M, Schoon Y, Bleeker-Rovers CP, Schers H, et al. Comprehensive health assessment three months after recovery from acute COVID-19. Clin Infect Dis [Internet]. [cited 2020 Nov 25]; Available from: https://academic.oup.com/cid/advance-article/doi/10.1093/cid/ciaa1750/5998118

10. Chow EJ, Rolfes MA, O’Halloran A, Anderson EJ, Bennett NM, Billing L, et al. Acute Cardiovascular Events Associated With Influenza in Hospitalized Adults. Ann Intern Med. 2020 Aug 25;173(8):605– 13.

11. Sellers SA, Hagan RS, Hayden FG, Fischer WA. The hidden burden of influenza: A review of the extra-pulmonary complications of influenza infection. Influenza Other Respir Viruses. 2017;11(5):372– 93.

12. Das KM, Lee EY, Singh R, Enani MA, Dossari KA, Gorkom KV, et al. Follow-up chest radiographic findings in patients with MERS-CoV after recovery. Indian J Radiol Imaging. 2017 Jul 1;27(3):342.

13. Rogers JP, Chesney E, Oliver D, Pollak TA, McGuire P, Fusar-Poli P, et al. Psychiatric and neuropsychiatric presentations associated with severe coronavirus infections: a systematic review and meta-analysis with comparison to the COVID-19 pandemic. Lancet Psychiatry. 2020 Jul 1;7(7):611–27.

14. Wu Q, Zhou L, Sun X, Yan Z, Hu C, Wu J, et al. Altered Lipid Metabolism in Recovered SARS Patients Twelve Years after Infection | Scientific Reports. Sci Rep. 2017 Aug 22;7(1):9110.

15. Docherty AB, Harrison EM, Green CA, Hardwick HE, Pius R, Norman L, et al. Features of 20 133 UK patients in hospital with covid-19 using the ISARIC WHO Clinical Characterisation Protocol: prospective observational cohort study. BMJ [Internet]. 2020 May 22 [cited 2020 Nov 25];369. Available from: https://www.bmj.com/content/369/bmj.m1985

16. Lipsitch M, Tchetgen Tchetgen E, Cohen T. Negative controls: a tool for detecting confounding and bias in observational studies. Epidemiol Camb Mass. 2010 May;21(3):383–8.

17. Chapter 3 | AHRQ Archive [Internet]. [cited 2020 Nov 25]. Available from: https://archive.ahrq.gov/research/findings/final-reports/medicareindicators/medicareindicators3.html

18. Austin PC. An Introduction to Propensity Score Methods for Reducing the Effects of Confounding in Observational Studies. Multivar Behav Res. 2011 May;46(3):399–424.

19. Cessie S le, Houwelingen JC van. Ridge Estimators in Logistic Regression. J R Stat Soc Ser C Appl Stat. 1992;41(1):191–201.

20. Schaefer RL, Roi LD, Wolfe RA. A ridge logistic estimator. Commun Stat - Theory Methods. 1984 Jan 1;13(1):99–113.

21. Schoenfeld D. Partial residuals for the proportional hazards regression model. Biometrika. 1982 Apr 1;69(1):239–41.

22. Kassambara A, Kosinski M, Biecek P, Fabian S. survminer: Drawing Survival Curves using “ggplot2” [Internet]. 2020 [cited 2021 Mar 8]. Available from: https://CRAN.R-project.org/package=survminer

23. Therneau TM, until 2009) TL (original S->R port and R maintainer, Elizabeth A, Cynthia C. survival: Survival Analysis [Internet]. 2020 [cited 2021 Mar 8]. Available from: https://CRAN.R-project.org/package=survival

24. Friedman JH, Hastie T, Tibshirani R. Regularization Paths for Generalized Linear Models via Coordinate Descent. J Stat Softw. 2010 Feb 2;33(1):1–22.

25. R: The R Project for Statistical Computing [Internet]. [cited 2021 Mar 8]. Available from: https://www.r-project.org/

26. Bhatla A, Mayer MM, Adusumalli S, Hyman MC, Oh E, Tierney A, et al. COVID-19 and cardiac arrhythmias. Heart Rhythm. 2020 Sep 1;17(9):1439–44.

27. Malas MB, Naazie IN, Elsayed N, Mathlouthi A, Marmor R, Clary B. Thromboembolism risk of COVID-19 is high and associated with a higher risk of mortality: A systematic review and meta-analysis. EClinicalMedicine [Internet]. 2020 Dec 1 [cited 2021 Mar 8];29. Available from: https://www.thelancet.com/journals/eclinm/article/PIIS2589-5370(20)30383-7/abstract

28. Vindegaard N, Benros ME. COVID-19 pandemic and mental health consequences: Systematic review of the current evidence. Brain Behav Immun. 2020 Oct 1;89:531–42.

29. Ye M, Ren Y, Lv T. Encephalitis as a clinical manifestation of COVID-19. Brain Behav Immun. 2020 Aug 1;88:945–6.

30. Helms J, Kremer S, Merdji H, Clere-Jehl R, Schenck M, Kummerlen C, et al. Neurologic Features in Severe SARS-CoV-2 Infection. N Engl J Med. 2020 Jun 4;382(23):2268–70.

31. Sathish T, Kapoor N, Cao Y, Tapp RJ, Zimmet P. Proportion of newly diagnosed diabetes in COVID-19 patients: A systematic review and meta-analysis. Diabetes Obes Metab. 2021;23(3):870–4.

32. Garrigues E, Janvier P, Kherabi Y, Bot AL, Hamon A, Gouze H, et al. Post-discharge persistent symptoms and health-related quality of life after hospitalization for COVID-19. J Infect. 2020 Dec 1;81(6):e4–6.

33. Moreno-Pérez O, Merino E, Leon-Ramirez J-M, Andres M, Ramos JM, Arenas-Jiménez J, et al. Post-acute COVID-19 syndrome. Incidence and risk factors: A Mediterranean cohort study. J Infect. 2021 Jan 12;

34. Hill JB, Garcia D, Crowther M, Savage B, Peress S, Chang K, et al. Frequency of venous thromboembolism in 6513 patients with COVID-19: a retrospective study. Blood Adv. 2020 Nov 2;4(21):5373–7.

35. Huang C, Huang L, Wang Y, Li X, Ren L, Gu X, et al. 6-month consequences of COVID-19 in patients discharged from hospital: a cohort study. The Lancet. 2021 Jan 16;397(10270):220–32.

36. Logue JK, Franko NM, McCulloch DJ, McDonald D, Magedson A, Wolf CR, et al. Sequelae in Adults at 6 Months After COVID-19 Infection. JAMA Netw Open. 2021 Feb 19;4(2):e210830.

37. Chopra V, Flanders SA, O’Malley M, Malani AN, Prescott HC. Sixty-Day Outcomes Among Patients Hospitalized With COVID-19. Ann Intern Med [Internet]. 2020 Nov 11 [cited 2021 Mar 8]; Available from: https://www.acpjournals.org/doi/10.7326/M20-5661

38. Dennis A, Wamil M, Kapur S, Alberts J, Badley AD, Decker GA, et al. Multi-organ impairment in low-risk individuals with long COVID [Internet]. Health Policy; 2020 Oct [cited 2021 Mar 8]. Available from: http://medrxiv.org/lookup/doi/10.1101/2020.10.14.20212555

39. Lau S-T, Yu W-C, Mok N-S, Tsui P-T, Tong W-L, Stella WCC. Tachycardia amongst subjects recovering from severe acute respiratory syndrome (SARS). Int J Cardiol. 2005 Apr 8;100(1):167–9.

40. Yu C-M, Wong RS-M, Wu EB, Kong S-L, Wong J, Yip GW-K, et al. Cardiovascular complications of severe acute respiratory syndrome. Postgrad Med J. 2006 Feb 1;82(964):140–4.

41. Ellul MA, Benjamin L, Singh B, Lant S, Michael BD, Easton A, et al. Neurological associations of COVID-19. Lancet Neurol. 2020 Sep 1;19(9):767–83.

42. Taquet M, Luciano S, Geddes JR, Harrison PJ. Bidirectional associations between COVID-19 and psychiatric disorder: retrospective cohort studies of 62 354 COVID-19 cases in the USA. Lancet Psychiatry [Internet]. 2020 Nov 9 [cited 2020 Nov 25];0(0). Available from: https://www.thelancet.com/journals/lanpsy/article/PIIS2215-0366(20)30462-4/abstract

43. Cunningham JW, Vaduganathan M, Claggett BL, Jering KS, Bhatt AS, Rosenthal N, et al. Clinical Outcomes in Young US Adults Hospitalized With COVID-19. JAMA Intern Med [Internet]. 2020 Sep 9 [cited 2020 Nov 25]; Available from: https://jamanetwork.com/journals/jamainternalmedicine/fullarticle/2770542

44. Davis HE, Assaf GS, McCorkell L, Wei H, Low RJ, Re’em Y, et al. Characterizing Long COVID in an International Cohort: 7 Months of Symptoms and Their Impact [Internet]. Infectious Diseases (except HIV/AIDS); 2020 Dec [cited 2021 Mar 8]. Available from: http://medrxiv.org/lookup/doi/10.1101/2020.12.24.20248802

45. Quan H, Li B, Saunders LD, Parsons GA, Nilsson CI, Alibhai A, et al. Assessing Validity of ICD-9-CM and ICD-10 Administrative Data in Recording Clinical Conditions in a Unique Dually Coded Database. Health Serv Res. 2008;43(4):1424–41.

46. Crabb BT, Lyons A, Bale M, Martin V, Berger B, Mann S, et al. Comparison of International Classification of Diseases and Related Health Problems, Tenth Revision Codes With Electronic Medical Records Among Patients With Symptoms of Coronavirus Disease 2019. JAMA Netw Open. 2020 Aug 14;3(8):e2017703.

