## Supplemental Tables and Figures for "SARS-CoV-2 infection and risk of clinical sequelae during the post-acute phase: a retrospective cohort study"

#### Table 1: ICD-10 Diagnostic Codes Utilized to Identify New Clinical Sequelae

| | |
|---|---|
| **Hypertension** | |
| Hypertension | I10, I11, I12, I13, I14, I15, I16 |
| Pulmonary hypertension | I27 |
| **Cardiovascular Outcomes** | |
| Arrhythmia | |
| > Cardiac Arrhythmia | I49 |
| > Tachycardia | I47, R00.0 |
| > Postural orthostatic tachycardia syndrome (POTs) | I49.9 |
| Congestive heart failure | I50.3, I50.2 |
| Cardiomyopathy | I42.4, I42.2, I42.3, I42.0, I25.5, I42.1, I43, I42.5, B33.24, A36.81, I42.9, I42.8 |
| Myocarditis | I40, B33.22, I51.4 |
| Coronary disease | |
| > Myocardial infarction | I21, I22 |
| > Acute coronary syndrome | I20.0, I21, I24 |
| > Cardiogenic shock | R57.0 |
| Hypercoagulability, DVT, PE | |
| > Hypercoagulability | D68, I82 |
| > Deep vein thrombosis | I82 |
| > Pulmonary embolism | I26.0, I26.9 |
| Stroke | |
| > Ischemic stroke | H34.1, I63, I64 |
| > Hemorrhagic stroke | I60, I61, I62 |
| **Mental health** | |
| Mental Health Diagnosis | |
| > Anxiety Disorder | F41.1, F41.3, F41.8, F41.9<br>*Remark: F41.0 is panic disorder, not included.* |
| > Depression diagnosis | F32, F33 |
| > PTSD | F43.1 |
| **Neurological Outcomes**[xi] | |

| | |
|---|---|
| Amnesia/Memory Difficulty | R41 |
| Migraine | G43 |
| Peripheral neuropathy | G62.9, G63 |
| Encephalopathy | G93.4 |
| Seizures | G40 |
| Dementia | G31.0, F02, F01.5, F03 |
| Guillain-Barre Syndrome | G61.0 |
| Alzheimer | G30 |
| **Diabetes** | |
| T2D diagnosis | E11 |
| **Liver** | |
| Liver Test Abnormality | I85, B18, I86.4, K70, K71, K72, K73, K74, K75, K76 |
| **Nephrology Outcomes**[xiii] | |
| Kidney Injury | |
| > Chronic kidney disease | N18, I12, I13 |
| > Acute kidney injury | N17 |
| **Pulmonary outcomes** | |
| Respiratory Failure | |
| > Acute respiratory failure | J96.0 |
| > Chronic respiratory failure | J96.1 |
| Interstitial lung disease | J84 |
| **Dermatologic Outcomes**[x] | |
| Atopic Dermatitis diagnosis | L20 |
| Urticaria | L50 |
| Herpesviral vesicular dermatitis | B00.1 |
| **Other Outcomes** | |
| Sleep Apnea | G47.3 |
| **Symptoms** | |
| Anosmia | R43.0 |
| Fatigue Diagnosis | |
| > Chronic Fatigue | R53.82 |
| > Fatigue | R53 |
| > Postviral fatigue | G93.3 |
| Myalgia | B33, M79 |

### Table 1a: Demographics, Co-morbidities, and Clinical Factors Among Propensity Score-Matched Adults 18 to ≤65y, UnitedHealth Group Clinical Discovery Database through October 31, 2020

| | **2020-matched 2020 SARS-CoV-2 Infected n()** | **2020 Comparator n()** | **2019-matched 2020 SARS-CoV-2 Infected** | **Matched 2019 Comparator n()** | **vLRTI-matched 2020 SARS-CoV-2 Infected** | **Matched Viral LRTI Comparator n()** |
|---|---|---|---|---|---|---|
| Total | 266586 | 266586 | 266586 | 266586 | 244276 | 244276 |
| **Demographic** | | | | | | |
| age: Mean (SD) | 41.7 (13.9) | 41.6 (13.8) * | 41.7 (13.9) | 41.6 (13.8) * | 41.9 (13.9) | 42.8 (13.6) * |
| age <= 17.00 | 0 (0.0) | 0 (0.0) | 0 (0.0) | 0 (0.0) | 0 (0.0) | 0 (0.0) |
| age >17.00, <=34.00 | 90497 (33.9) | 91349 (34.3) | 90497 (33.9) | 90896 (34.1) | 81254 (33.3) | 72415 (29.6) |
| age >34.00, <=49.00 | 82497 (30.9) | 84991 (31.9) | 82497 (30.9) | 86057 (32.3) | 75458 (30.9) | 82416 (33.7) |
| age > 49.00 | 93592 (35.1) | 90246 (33.9) | 93592 (35.1) | 89633 (33.6) | 87564 (35.8) | 89445 (36.6) |
| Gender (male) | 126980 (47.6) | 126653 (47.5) | 126980 (47.6) | 126470 (47.4) | 114658 (46.9) | 115036 (47.1) |
| SES_index: Median (IQR) | 52.9 (51.2 - 55.1) | 52.9 (51.2 - 55.0) | 52.9 (51.2 - 55.1) | 52.9 (51.2 - 55.0) | 53.0 (51.2 - 55.2) | 53.0 (51.4 - 55.5) * |
| white_proportion: Mean (SD) | 0.60 (0.26) | 0.60 (0.26) | 0.60 (0.26) | 0.60 (0.27) | 0.62 (0.25) | 0.61 (0.26) * |
| white_proportion <= 0.25 | 36954 (13.9) | 37854 (14.2) | 36954 (13.9) | 38089 (14.3) | 29638 (12.1) | 30765 (12.6) |
| white_proportion >0.25, <=0.50 | 48299 (18.1) | 47799 (17.9) | 48299 (18.1) | 47686 (17.9) | 42725 (17.5) | 43587 (17.8) |
| white_proportion >0.50, <=0.75 | 85969 (32.2) | 83358 (31.3) | 85969 (32.2) | 83934 (31.5) | 79901 (32.7) | 78572 (32.2) |
| white_proportion > 0.75 | 95364 (35.8) | 97575 (36.6) | 95364 (35.8) | 96877 (36.3) | 92012 (37.7) | 91352 (37.4) |
| black_proportion: Mean (SD) | 0.13 (0.17) | 0.13 (0.18) | 0.13 (0.17) | 0.13 (0.18) | 0.12 (0.17) | 0.12 (0.17) |
| black_proportion <= 0.25 | 226443 (84.9) | 226523 (85.0) | 226443 (84.9) | 226367 (84.9) | 209234 (85.7) | 209309 (85.7) |
| black_proportion >0.25, <=0.50 | 25327 (9.5) | 24138 (9.1) | 25327 (9.5) | 24154 (9.1) | 22629 (9.3) | 21829 (8.9) |
| black_proportion >0.50, <=0.75 | 10241 (3.8) | 10601 (4.0) | 10241 (3.8) | 10735 (4.0) | 8709 (3.6) | 8789 (3.6) |
| black_proportion > 0.75 | 4575 (1.7) | 5324 (2.0) | 4575 (1.7) | 5330 (2.0) | 3704 (1.5) | 4349 (1.8) |
| hispanic_proportion: Mean (SD) | 0.19 (0.21) | 0.19 (0.20) | 0.19 (0.21) | 0.19 (0.20) | 0.18 (0.19) | 0.18 (0.20) |

| | | | | | | |
|---|---|---|---|---|---|---|
| hispanic_proportion <= 0.25 | 201097 (75.4) | 200370 (75.2) | 201097 (75.4) | 200362 (75.2) | 188860 (77.3) | 188881 (77.3) |
| hispanic_proportion >0.25, <=0.50 | 39727 (14.9) | 40778 (15.3) | 39727 (14.9) | 41028 (15.4) | 35016 (14.3) | 35336 (14.5) |
| hispanic_proportion >0.50, <=0.75 | 16934 (6.4) | 16819 (6.3) | 16934 (6.4) | 16501 (6.2) | 13739 (5.6) | 13213 (5.4) |
| hispanic_proportion > 0.75 | 8828 (3.3) | 8619 (3.2) | 8828 (3.3) | 8695 (3.3) | 6661 (2.7) | 6846 (2.8) |
| **Comorbidity** | | | | | | |
| Any | 120716 (45.3) | 116617 (43.7) * | 120716 (45.3) | 116515 (43.7) * | 115693 (47.4) | 132954 (54.4) * |
| Thalassemia | 226 (0.1) | 194 (0.1) | 226 (0.1) | 229 (0.1) | 212 (0.1) | 213 (0.1) |
| AIDS/HIV | 992 (0.4) | 979 (0.4) | 992 (0.4) | 948 (0.4) | 894 (0.4) | 933 (0.4) |
| Alcohol Abuse | 2959 (1.1) | 2867 (1.1) | 2959 (1.1) | 2920 (1.1) | 2825 (1.2) | 2758 (1.1) |
| anemia | 16157 (6.1) | 15523 (5.8) * | 16157 (6.1) | 15475 (5.8) * | 15412 (6.3) | 16248 (6.7) * |
| Rheumatoid Arthritis | 6793 (2.5) | 6656 (2.5) | 6793 (2.5) | 6480 (2.4) | 6661 (2.7) | 7133 (2.9) * |
| Cerebrovascular Disease | 4342 (1.6) | 4046 (1.5) | 4342 (1.6) | 4071 (1.5) | 4204 (1.7) | 4605 (1.9) * |
| Congestive Heart Failure | 3362 (1.3) | 2936 (1.1) * | 3362 (1.3) | 3090 (1.2) * | 3265 (1.3) | 3685 (1.5) * |
| Coagulopathy | 2193 (0.8) | 2032 (0.8) | 2193 (0.8) | 1986 (0.7) | 2103 (0.9) | 2621 (1.1) * |
| Chronic Pulmonary Disease | 22236 (8.3) | 21689 (8.1) | 22236 (8.3) | 21895 (8.2) | 22020 (9.0) | 23210 (9.5) * |
| Dementia | 205 (0.1) | 192 (0.1) | 205 (0.1) | 187 (0.1) | 192 (0.1) | 234 (0.1) |
| Depression | 19656 (7.4) | 19636 (7.4) | 19656 (7.4) | 19558 (7.3) | 19038 (7.8) | 20301 (8.3) * |
| Drug Abuse | 2485 (0.9) | 2439 (0.9) | 2485 (0.9) | 2467 (0.9) | 2397 (1.0) | 2722 (1.1) * |
| Hypertension | 51908 (19.5) | 51656 (19.4) | 51908 (19.5) | 51363 (19.3) | 50231 (20.6) | 54705 (22.4) * |
| Hypothyroidism | 17335 (6.5) | 17117 (6.4) | 17335 (6.5) | 17090 (6.4) | 17058 (7.0) | 17247 (7.1) |
| Liver Disease | 9017 (3.4) | 8612 (3.2) | 9017 (3.4) | 8682 (3.3) | 8657 (3.5) | 9638 (3.9) * |
| Moderate to Severe Liver Disease | 379 (0.1) | 312 (0.1) | 379 (0.1) | 341 (0.1) | 363 (0.1) | 421 (0.2) |
| Lymphoma | 802 (0.3) | 726 (0.3) | 802 (0.3) | 686 (0.3) | 777 (0.3) | 1040 (0.4) * |
| Fluid and Electrolyte Disorders | 8493 (3.2) | 7955 (3.0) * | 8493 (3.2) | 7972 (3.0) * | 8185 (3.4) | 8608 (3.5) |
| Metastatic Cancer | 1415 (0.5) | 1256 (0.5) | 1415 (0.5) | 1316 (0.5) | 1390 (0.6) | 1487 (0.6) |
| Myocardial Infarction | 1499 (0.6) | 1333 (0.5) | 1499 (0.6) | 1468 (0.6) | 1459 (0.6) | 1622 (0.7) |

| | | | | | | |
|---|---|---|---|---|---|---|
| Other Neurological Disorders | 6514 (2.4) | 6283 (2.4) | 6514 (2.4) | 6412 (2.4) | 6313 (2.6) | 6854 (2.8) * |
| Obesity | 36149 (13.6) | 35930 (13.5) | 36149 (13.6) | 35652 (13.4) | 34540 (14.1) | 35405 (14.5) * |
| Paralysis | 774 (0.3) | 727 (0.3) | 774 (0.3) | 684 (0.3) | 740 (0.3) | 1118 (0.5) * |
| Peripheral Vascular Disease | 4861 (1.8) | 4398 (1.6) * | 4861 (1.8) | 4509 (1.7) * | 4713 (1.9) | 5234 (2.1) * |
| Psychoses | 4878 (1.8) | 4833 (1.8) | 4878 (1.8) | 4891 (1.8) | 4738 (1.9) | 5293 (2.2) * |
| Peptic Ulcer Disease | 1168 (0.4) | 1074 (0.4) | 1168 (0.4) | 1076 (0.4) | 1111 (0.5) | 1280 (0.5) * |
| Pulmonary Circulation Disorder | 1022 (0.4) | 901 (0.3) | 1022 (0.4) | 912 (0.3) | 975 (0.4) | 1227 (0.5) * |
| Renal Failure | 3893 (1.5) | 3600 (1.4) * | 3893 (1.5) | 3567 (1.3) * | 3734 (1.5) | 4324 (1.8) * |
| Solid Tumor without Metastasis | 6824 (2.6) | 6624 (2.5) | 6824 (2.6) | 6526 (2.4) | 6686 (2.7) | 7440 (3.0) * |
| Valvular Disease | 6331 (2.4) | 5954 (2.2) * | 6331 (2.4) | 6173 (2.3) | 6155 (2.5) | 6830 (2.8) * |
| Weight Loss | 2909 (1.1) | 2827 (1.1) | 2909 (1.1) | 2817 (1.1) | 2766 (1.1) | 3203 (1.3) * |
| **Prior Conditions** | | | | | | |
| Alzheimers_Dementia | 385 (0.1) | 388 (0.1) | 385 (0.1) | 338 (0.1) | 358 (0.1) | 489 (0.2) * |
| Asthma | 15112 (5.7) | 14757 (5.5) | 15112 (5.7) | 14845 (5.6) | 14969 (6.1) | 14976 (6.1) |
| Cystic_Fibrosis | 64 (0.0) | 64 (0.0) | 64 (0.0) | 55 (0.0) | 60 (0.0) | 144 (0.1) * |
| Immunodeficiency | 3101 (1.2) | 2877 (1.1) | 3101 (1.2) | 2876 (1.1) | 2932 (1.2) | 3174 (1.3) |
| Pulmonary_Fibrosis | 488 (0.2) | 434 (0.2) | 488 (0.2) | 433 (0.2) | 480 (0.2) | 615 (0.3) * |
| Sickle_Cell_Disease | 273 (0.1) | 224 (0.1) | 273 (0.1) | 240 (0.1) | 249 (0.1) | 282 (0.1) |
| Smoking | 8113 (3.0) | 8141 (3.1) | 8113 (3.0) | 8161 (3.1) | 8068 (3.3) | 8086 (3.3) |
| T1D | 2157 (0.8) | 2015 (0.8) | 2157 (0.8) | 2124 (0.8) | 2090 (0.9) | 2408 (1.0) * |
| T2D | 22619 (8.5) | 21993 (8.2) | 22619 (8.5) | 21784 (8.2) * | 21647 (8.9) | 23863 (9.8) * |
| Prior Length of Stay in Hospital (days): Mean (SD) | 0.5 (5.9) | 0.5 (4.3) * | 0.5 (5.9) | 0.4 (4.8) * | 0.5 (5.4) | 0.7 (8.9) * |
| Prior PCP visit | 186313 (69.9) | 170543 (64.0) * | 186313 (69.9) | 172404 (64.7) * | 171608 (70.3) | 174126 (71.3) * |
| Prior PCP_visit: Mean (SD) | 2.5 (3.5) | 2.4 (3.8) * | 2.5 (3.5) | 2.4 (3.8) * | 2.5 (3.4) | 2.6 (3.4) |

| | | | | | | |
|---|---|---|---|---|---|---|
| Prior cardiology_visit: Mean (SD) | 0.3 (1.7) | 0.3 (1.6) * | 0.3 (1.7) | 0.3 (1.5) * | 0.3 (1.6) | 0.3 (2.0) |
| Prior nephrology_visit: Mean (SD) | 0.1 (1.6) | 0.1 (1.5) * | 0.1 (1.6) | 0.1 (1.2) * | 0.1 (1.4) | 0.1 (2.3) * |
| **Region** | | | | | | |
| South | 117292 (44.0) | 110703 (41.5) * | 117292 (44.0) | 111886 (42.0) * | 108320 (44.3) | 103977 (42.6) * |
| Midwest | 54494 (20.4) | 56566 (21.2) * | 54494 (20.4) | 56105 (21.0) * | 51483 (21.1) | 49625 (20.3) * |
| Northeast | 56071 (21.0) | 56947 (21.4) * | 56071 (21.0) | 57402 (21.5) * | 50909 (20.8) | 55964 (22.9) * |
| West | 38300 (14.4) | 41834 (15.7) * | 38300 (14.4) | 40661 (15.3) * | 33157 (13.6) | 34272 (14.0) * |
| Unknown | 429 (0.2) | 536 (0.2) * | 429 (0.2) | 532 (0.2) * | 407 (0.2) | 438 (0.2) * |

**Figure 1(a): Absolute Standardized Mean Difference before and after propensity score matching for key variables including demographic, comorbidities and previous medical utilization, with 2020 comparator group, UnitedHealth Group Clinical Discovery Database through October 31, 2020.**

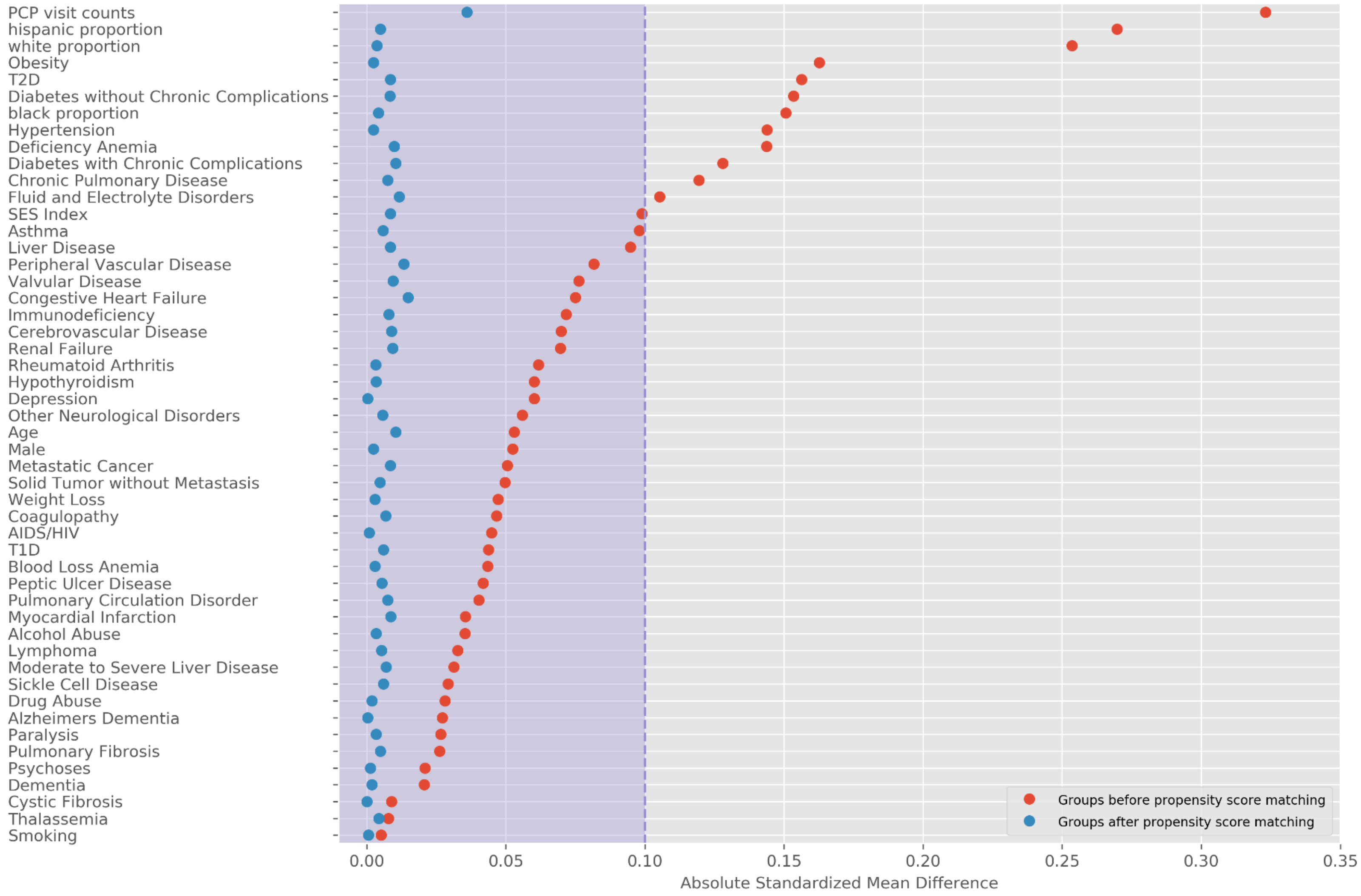

**Figure 1(b): Absolute standardized mean difference before and after matching SARS-CoV-2 infection group with 2019 comparator group, UnitedHealth Group Clinical Discovery Database through October 31, 2020.**

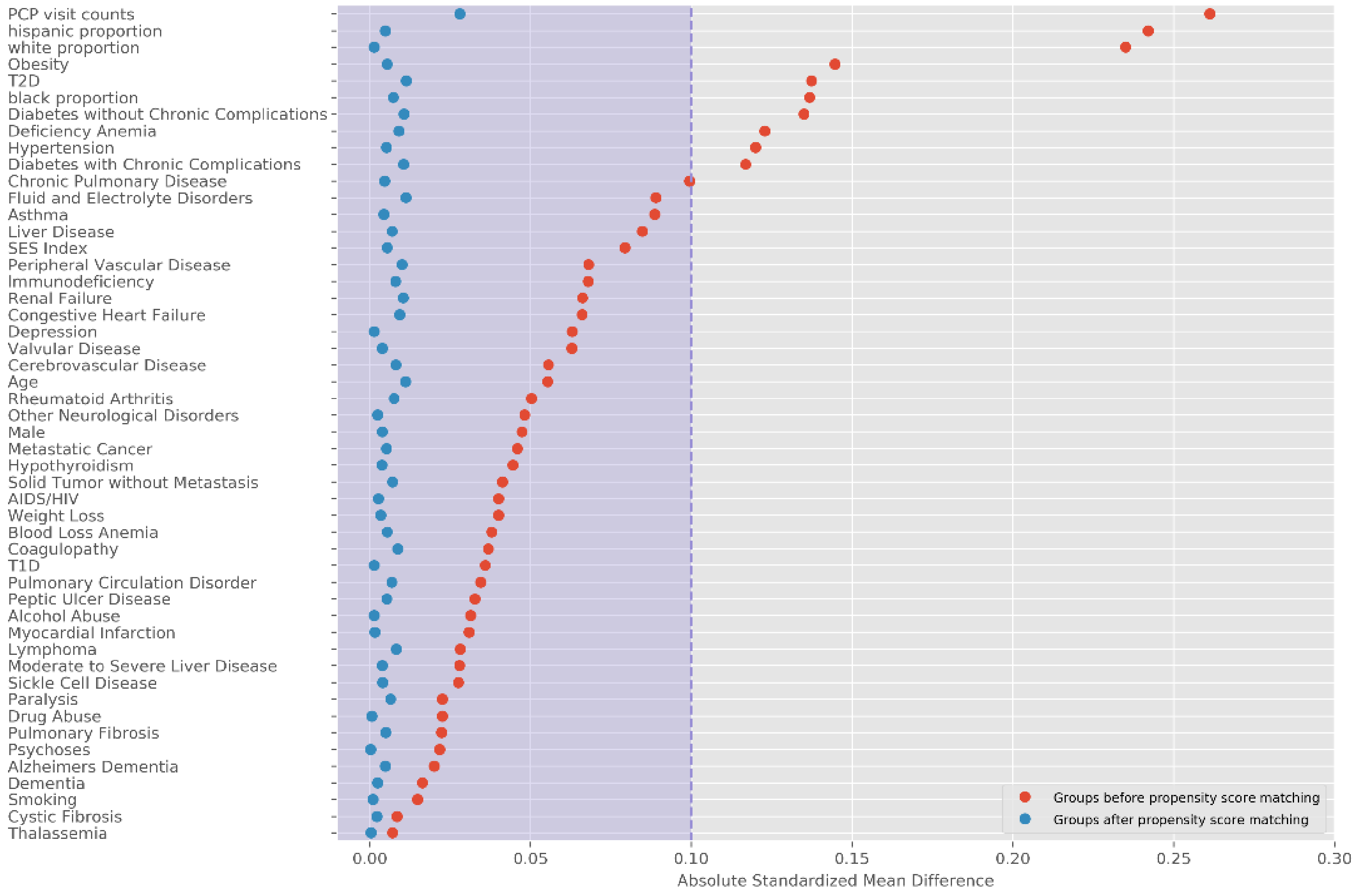

**Figure 1(c): Absolute standardized mean difference before and after matching SARS-CoV-2 infection group with viral LRTI comparator group, UnitedHealth Group Clinical Discovery Database through October 31, 2020.**

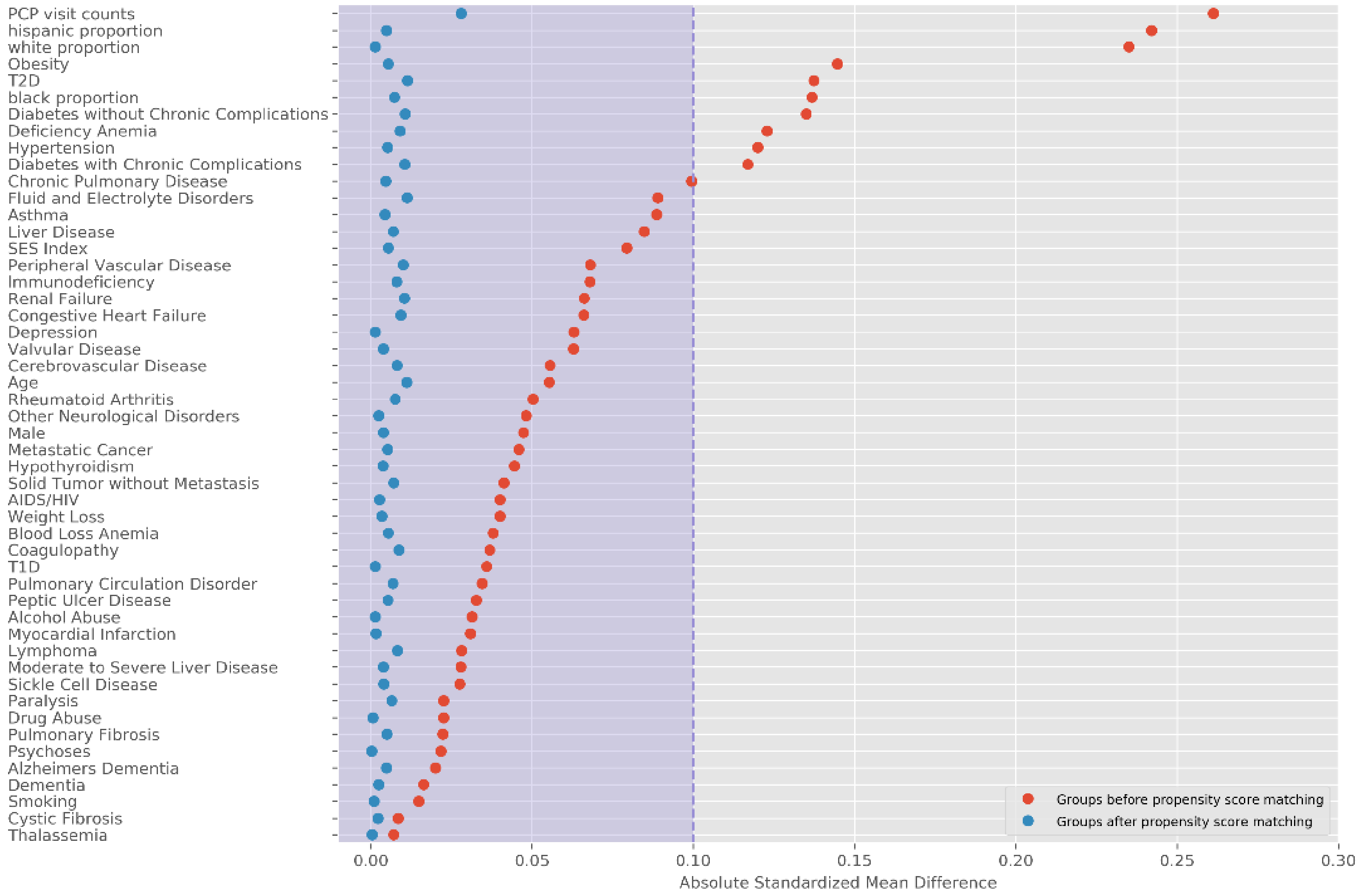

**Table 2a: Incidence and Risk Difference (per 100 individuals) for Clinical Sequelae by SARS-CoV-2 Infection and Comparator Groups, UnitedHealth Group Clinical Discovery Database through October 31, 2020.**

| Outcome | 2020 SARS-CoV-2 Infected † | 2020 Comparator group † | Risk Difference (CI ††) | 2019-matched 2020 SARS-CoV-2 Infected † | 2019 Comparator group † | Risk Difference (CI ††) | Viral LRTI-matched 2020 SARS-CoV-2 Infected † | Viral LRTI Comparator group † | Risk Difference (CI ††) |
|---|---|---|---|---|---|---|---|---|---|
| **Matched Group Size** | 193113 ††† | 193113 ††† | | 193264 ††† | 193264 ††† | | 181613 ††† | 181613 ††† | |
| **Follow-up period (days): median (IQR) ††††** | 87 (45 – 124) | 87 (45 – 124) | | 87 (45 – 124) | 87 (46 – 124) | | 87 (45 – 124) | 87 (43 – 130) | |
| **Hypertension** | | | | | | | | | |
| Hypertension | 2.59 | 1.63 | 0.96 (0.70 to 1.20) *** | 2.59 | 1.70 | 0.88 (0.63 to 1.14) *** | 2.60 | 2.47 | 0.13 (-0.18 to 0.41) |
| Pulmonary hypertension | 0.14 | 0.04 | 0.10 (0.06 to 0.14) *** | 0.14 | 0.05 | 0.08 (0.04 to 0.13) *** | 0.15 | 0.16 | -0.01 (-0.07 to 0.04) |
| **Cardiology** | | | | | | | | | |
| Arrythmia | 1.49 | 0.63 | 0.86 (0.73 to 1.00) *** | 1.52 | 0.69 | 0.83 (0.71 to 0.97) *** | 1.52 | 1.09 | 0.44 (0.26 to 0.62) *** |
| > Cardiac arrhythmia | 0.72 | 0.31 | 0.41 (0.31 to 0.49) *** | 0.72 | 0.37 | 0.35 (0.25 to 0.46) *** | 0.72 | 0.51 | 0.21 (0.10 to 0.31) *** |
| > Tachycardia | 1.07 | 0.42 | 0.65 (0.53 to 0.75) *** | 1.08 | 0.42 | 0.66 (0.54 to 0.77) *** | 1.11 | 0.77 | 0.34 (0.20 to 0.48) *** |
| > POTs | 0.28 | 0.13 | 0.16 (0.10 to 0.22) *** | 0.29 | 0.16 | 0.13 (0.07 to 0.20) *** | 0.29 | 0.22 | 0.07 (0.01 to 0.13) *** |
| Congestive heart failure | 0.21 | 0.09 | 0.12 (0.06 to 0.18) *** | 0.21 | 0.10 | 0.11 (0.06 to 0.16) *** | 0.23 | 0.23 | 0.00 (-0.06 to 0.07) |
| Cardiomyopathy | 0.23 | 0.07 | 0.15 (0.10 to 0.20) *** | 0.23 | 0.10 | 0.14 (0.09 to 0.20) *** | 0.23 | 0.21 | 0.02 (-0.04 to 0.09) |
| Myocarditis | 0.04 | 0.00 | 0.04 (0.02 to 0.07) *** | 0.04 | 0.00 | 0.04 (0.02 to 0.06) *** | 0.04 | 0.01 | 0.03 (0.01 to 0.05) *** |
| Coronary disease | 0.21 | 0.10 | 0.11 (0.05 to 0.16) *** | 0.21 | 0.12 | 0.08 (0.03 to 0.13) *** | 0.22 | 0.23 | -0.01 (-0.07 to 0.06) |

| | | | | | | | | | |
|---|---|---|---|---|---|---|---|---|---|
| > Myocardial infarction | 0.13 | 0.06 | 0.07 (0.03 to 0.11) *** | 0.13 | 0.08 | 0.06 (0.02 to 0.10) *** | 0.14 | 0.13 | 0.01 (-0.04 to 0.06) |
| > Acute coronary syndrome | 0.19 | 0.10 | 0.10 (0.05 to 0.14) *** | 0.20 | 0.12 | 0.08 (0.02 to 0.13) *** | 0.20 | 0.21 | -0.01 (-0.08 to 0.05) |
| > Cardiogenic shock | 0.02 | 0.01 | 0.02 (0.00 to 0.03) *** | 0.02 | 0.01 | 0.01 (-0.00 to 0.03) | 0.02 | 0.02 | -0.00 (-0.02 to 0.02) |
| Hypercoagulability DVT PE | 0.60 | 0.20 | 0.40 (0.33 to 0.48) *** | 0.62 | 0.21 | 0.41 (0.33 to 0.50) *** | 0.64 | 0.43 | 0.20 (0.11 to 0.30) *** |
| > Hypercoagulability | 0.50 | 0.17 | 0.33 (0.26 to 0.40) *** | 0.53 | 0.18 | 0.35 (0.27 to 0.43) *** | 0.54 | 0.39 | 0.15 (0.05 to 0.24) *** |
| > Deep vein thrombosis | 0.34 | 0.10 | 0.23 (0.17 to 0.30) *** | 0.35 | 0.10 | 0.25 (0.20 to 0.31) *** | 0.36 | 0.25 | 0.10 (0.03 to 0.18) *** |
| > Pulmonary embolism | 0.19 | 0.06 | 0.13 (0.09 to 0.18) *** | 0.19 | 0.06 | 0.13 (0.08 to 0.18) *** | 0.20 | 0.13 | 0.07 (0.02 to 0.12) *** |
| Stroke | 0.22 | 0.11 | 0.11 (0.06 to 0.17) *** | 0.21 | 0.14 | 0.08 (0.03 to 0.14) *** | 0.22 | 0.18 | 0.05 (-0.02 to 0.10) |
| > Ischemic stroke | 0.19 | 0.09 | 0.10 (0.05 to 0.14) *** | 0.18 | 0.11 | 0.07 (0.02 to 0.12) *** | 0.19 | 0.15 | 0.04 (-0.01 to 0.09) |
| > Hemorrhagic stroke | 0.06 | 0.03 | 0.03 (-0.00 to 0.05) | 0.06 | 0.04 | 0.02 (-0.00 to 0.05) | 0.06 | 0.05 | 0.01 (-0.02 to 0.04) |
| **Mental Health** | | | | | | | | | |
| Mental Health Diagnosis | 3.54 | 2.62 | 0.92 (0.67 to 1.19) *** | 3.59 | 2.47 | 1.12 (0.87 to 1.40) *** | 3.70 | 3.12 | 0.57 (0.27 to 0.88) *** |
| > Anxiety | 2.98 | 2.15 | 0.83 (0.60 to 1.02) *** | 3.04 | 1.97 | 1.07 (0.84 to 1.29) *** | 3.08 | 2.48 | 0.60 (0.32 to 0.84) *** |
| > Depression | 1.60 | 1.26 | 0.33 (0.17 to 0.51) *** | 1.58 | 1.32 | 0.26 (0.09 to 0.45) *** | 1.65 | 1.63 | 0.03 (-0.18 to 0.20) |
| > PTSD | 0.23 | 0.15 | 0.08 (0.03 to 0.15) *** | 0.22 | 0.16 | 0.05 (-0.01 to 0.11) | 0.22 | 0.15 | 0.07 (0.01 to 0.12) *** |
| **Neurology** | | | | | | | | | |
| Amnesia/Memory Difficulty | 0.69 | 0.32 | 0.37 (0.27 to 0.45) *** | 0.71 | 0.35 | 0.37 (0.26 to 0.46) *** | 0.72 | 0.48 | 0.24 (0.13 to 0.34) *** |
| Migraine | 0.76 | 0.58 | 0.18 (0.06 to 0.28) *** | 0.73 | 0.61 | 0.12 (0.01 to 0.23) *** | 0.77 | 0.78 | -0.01 (-0.13 to 0.10) |
| Peripheral neuropathy | 0.31 | 0.15 | 0.16 (0.10 to 0.23) *** | 0.30 | 0.16 | 0.14 (0.07 to 0.19) *** | 0.32 | 0.20 | 0.12 (0.05 to 0.20) *** |
| Encephalopathy | 0.23 | 0.03 | 0.19 (0.14 to 0.24) *** | 0.22 | 0.08 | 0.15 (0.10 to 0.20) *** | 0.23 | 0.14 | 0.09 (0.03 to 0.15) *** |
| Seizure | 0.14 | 0.08 | 0.06 (0.03 to 0.11) *** | 0.13 | 0.10 | 0.03 (-0.01 to 0.07) | 0.14 | 0.14 | -0.00 (-0.05 to 0.04) |

| | | | | | | | | | |
|---|---|---|---|---|---|---|---|---|---|
| Dementia | 0.04 | 0.01 | 0.02 (-0.00 to 0.04) | 0.03 | 0.02 | 0.01 (-0.01 to 0.03) | 0.04 | 0.03 | 0.00 (-0.02 to 0.03) |
| GuillianBarre Syndrome | 0.01 | 0.00 | 0.01 (-0.00 to 0.02) | 0.01 | 0.00 | 0.01 (-0.00 to 0.02) | 0.01 | 0.00 | 0.01 (-0.00 to 0.03) |
| Alzheimer | 0.01 | 0.00 | 0.00 (-0.01 to 0.01) | 0.00 | 0.00 | 0.00 (-0.01 to 0.01) | 0.00 | 0.00 | 0.00 (-0.01 to 0.01) |
| **Diabetes** | | | | | | | | | |
| Type 2 Diabetes | 1.04 | 0.57 | 0.47 (0.35 to 0.60) *** | 0.99 | 0.60 | 0.39 (0.25 to 0.53) *** | 1.05 | 0.77 | 0.28 (0.13 to 0.41) *** |
| **Liver** | | | | | | | | | |
| Liver Test Abnormality | 1.17 | 0.69 | 0.47 (0.35 to 0.59) *** | 1.20 | 0.69 | 0.50 (0.38 to 0.63) *** | 1.22 | 1.02 | 0.20 (0.06 to 0.33) *** |
| **Nephrology** | | | | | | | | | |
| Kidney Injury | 0.49 | 0.32 | 0.17 (0.08 to 0.25) *** | 0.51 | 0.34 | 0.17 (0.08 to 0.26) *** | 0.52 | 0.47 | 0.05 (-0.06 to 0.14) |
| > Chronic kidney disease | 0.35 | 0.25 | 0.11 (0.03 to 0.17) *** | 0.37 | 0.24 | 0.14 (0.06 to 0.21) *** | 0.37 | 0.31 | 0.06 (-0.02 to 0.13) |
| > Acute kidney injury | 0.37 | 0.15 | 0.23 (0.15 to 0.29) *** | 0.37 | 0.20 | 0.18 (0.10 to 0.26) *** | 0.38 | 0.33 | 0.05 (-0.03 to 0.14) |
| **Respiratory** | | | | | | | | | |
| Respiratory Failure | 0.22 | 0.06 | 0.16 (0.11 to 0.21) *** | 0.23 | 0.10 | 0.12 (0.07 to 0.18) *** | 0.23 | 0.27 | -0.04 (-0.11 to 0.02) |
| > Acute respiratory failure | 0.21 | 0.06 | 0.16 (0.11 to 0.21) *** | 0.21 | 0.10 | 0.12 (0.07 to 0.17) *** | 0.22 | 0.26 | -0.04 (-0.10 to 0.03) |
| > Chronic respiratory failure | 0.17 | 0.02 | 0.15 (0.12 to 0.19) *** | 0.17 | 0.02 | 0.15 (0.11 to 0.20) *** | 0.18 | 0.11 | 0.07 (0.02 to 0.12) *** |
| Interstitial lung disease | 0.28 | 0.04 | 0.24 (0.19 to 0.31) *** | 0.28 | 0.05 | 0.22 (0.16 to 0.28) *** | 0.28 | 0.23 | 0.05 (-0.02 to 0.11) |
| **Dermatology** | | | | | | | | | |
| Atopic dermatitis | 0.31 | 0.28 | 0.04 (-0.04 to 0.11) | 0.33 | 0.27 | 0.05 (-0.02 to 0.13) | 0.33 | 0.28 | 0.05 (-0.03 to 0.13) |
| Urticaria | 0.28 | 0.20 | 0.09 (0.02 to 0.14) *** | 0.28 | 0.25 | 0.03 (-0.04 to 0.08) | 0.28 | 0.36 | -0.08 (-0.15 to 0.00) |
| Herpesviral vesicular dermatitis | 0.10 | 0.07 | 0.03 (-0.01 to 0.07) | 0.11 | 0.10 | 0.00 (-0.04 to 0.05) | 0.10 | 0.12 | -0.01 (-0.06 to 0.02) |
| **Other** | | | | | | | | | |
| Sleep apnea | 1.17 | 0.67 | 0.50 (0.35 to 0.64) *** | 1.18 | 0.75 | 0.43 (0.30 to 0.55) *** | 1.23 | 1.36 | -0.13 (-0.28 to 0.05) |

† Cumulative incidence by Kaplan-Meier estimator 120 days after time origin (index date+21 days), which is 141 days after index date.

†† Confidence intervals are calculated based on Bonferroni correction, i.e., (1 - 0.05/N) confidence interval.

††† Individuals were excluded if they had less than index date + 21 days of follow-up, had already been diagnosed with outcome of interest prior to index date + 21 days or their matched pair was excluded for the same reasons.

†††† The outcome observation period is the length of follow-up (in days) measured from time origin (which is index date + 21 days) to end of observation of the members

*** p value < 0.001

** p value < 0.01

* p value < 0.05

**Table 2b: Incidence and Risk Difference (per 100 individuals) for Symptom Sequelae by SARS-CoV-2 Infection and Comparison Groups, UnitedHealth Group Clinical Discovery Database through October 31, 2020.**

| **Outcome** | **2020 SARS-CoV-2 Infected †** | **2020 Comparator group †** | **Risk Difference (CI ††)** | **2019-matched 2020 SARS-CoV-2 Infected †** | **2019 Comparator group †** | **Risk Difference (CI ††)** | **Viral LRTI-matched 2020 SARS-CoV-2 Infected †** | **Viral LRTI comparator group †** | **Risk Difference (CI ††)** |
|---|---|---|---|---|---|---|---|---|---|
| **Matched Group Size** | 193113 ††† | 193113 ††† | | 193264 ††† | 193264 ††† | | 181613 ††† | 181613 ††† | |
| **Follow-up period (days): median (IQR) ††††** | 87 (45 – 124) | 87 (45 – 124) | | 87 (45 – 124) | 87 (46 – 124) | | 87 (45 – 124) | 87 (43 – 130) | |
| Fatigue Diagnosis | 4.64 | 2.38 | 2.26 (1.97 to 2.53) *** | 4.68 | 2.33 | 2.35 (2.06 to 2.62) *** | 4.84 | 3.63 | 1.20 (0.92 to 1.50) *** |
| > Fatigue | 4.54 | 2.38 | 2.16 (1.87 to 2.47) *** | 4.57 | 2.32 | 2.25 (1.98 to 2.49) *** | 4.73 | 3.62 | 1.10 (0.78 to 1.45) *** |
| > Chronic Fatigue | 0.39 | 0.21 | 0.18 (0.11 to 0.26) *** | 0.40 | 0.24 | 0.16 (0.06 to 0.23) *** | 0.39 | 0.32 | 0.07 (-0.01 to 0.16) |
| > Postviral fatigue | 0.22 | 0.01 | 0.22 (0.18 to 0.25) *** | 0.23 | 0.01 | 0.22 (0.18 to 0.27) *** | 0.24 | 0.03 | 0.21 (0.16 to 0.26) *** |
| Myalgia | 4.34 | 2.90 | 1.44 (1.14 to 1.73) *** | 4.35 | 3.27 | 1.08 (0.80 to 1.44) *** | 4.53 | 4.23 | 0.30 (-0.08 to 0.66) |
| Anosmia | 0.23 | 0.04 | 0.19 (0.14 to 0.24) *** | 0.21 | 0.02 | 0.20 (0.16 to 0.24) *** | 0.22 | 0.04 | 0.18 (0.14 to 0.23) *** |

† Cumulative incidence by Kaplan-Meier estimator 120 days after time origin (index date+21 days), which is 141 days after index date.

†† Confidence intervals are calculated based on Bonferroni correction, i.e., (1 - 0.05/N) confidence interval.

††† Individuals were excluded if they had less than index date + 21 days of follow-up, had already been diagnosed with outcome of interest prior to index date + 21 days or their matched pair was excluded for the same reasons.

†††† The outcome observation period is the length of follow-up (in days) measured from time origin (which is index date + 21 days) to end of observation of the members

*** p value < 0.001

** p value < 0.01

* p value < 0.05

**Table 2c: Rates and Hazard Ratios for Clinical Sequelae by SARS-CoV-2 Infection and Comparison Groups, UnitedHealth Group Clinical Discovery Database through October 31, 2020.**

| Outcome | 2020 SARS-CoV-2 Infected: incidence rate (case/ person-year) | 2020 Comparator: incidence rate (case/ person-year) † | Hazard Ratio (95 CI ††) | 2019-matched 2020 SARS-CoV-2 Infected: incidence rate (case/ person-year) | 2019 Comparator: incidence rate (case/ person-year) † | Hazard Ratio (95 CI ††) | vLRTI-matched 2020 SARS-CoV-2 Infected: incidence rate (case/ person-year) | vLRTI comparator: incidence rate (case/ person-year) † | Hazard Ratio (95 CI ††) |
|---|---|---|---|---|---|---|---|---|---|
| **Matched Group Size** | **193113 †††** | **193113 †††** | | **193264 †††** | **193264 †††** | | **181613 †††** | **181613 †††** | |
| **Follow-up period (days): median (IQR) ††††** | **87 (45 – 124)** | **87 (45 – 124)** | | **87 (45 – 124)** | **87 (46 – 124)** | | **89 (45 – 135)** | **87 (43 – 130)** | |
| **Hypertension** | | | | | | | | | |
| Hypertension | 0.09 (2231 / 26249.51) | 0.05 (1374 / 27255.68) | 1.70 (1.55 to 1.87) *** | 0.09 (2240 / 26244.68) | 0.05 (1432 / 27414.12) | 1.59 (1.46 to 1.74) *** | 0.09 (2126 / 24690.74) | 0.08 (1921 / 25164.28) | 1.10 (1.01 to 1.19) ** |
| Pulmonary hypertension | 0.004 (212 / 47079.96) | 0.001 (61 / 47148.28) | 3.20 (2.07 to 4.95) *** | 0.004 (202 / 47071.03) | 0.002 (71 / 47439.48) | 3.14 (2.04 to 4.83) *** | 0.005 (207 / 45384.27) | 0.005 (228 / 46034.33) | 0.91 (0.68 to 1.24) |
| **Cardiology** | | | | | | | | | |
| Arrythmia | 0.05 (2044 / 42447.53) | 0.02 (840 / 43043.64) | 2.50 (2.22 to 2.83) *** | 0.05 (2080 / 42437.48) | 0.02 (943 / 43347.33) | 2.23 (1.99 to 2.50) *** | 0.05 (1977 / 40010.37) | 0.03 (1388 / 40748.83) | 1.46 (1.31 to 1.62) *** |
| > Cardiac arrhythmia | 0.02 (1017 / 45301.45) | 0.009 (434 / 45575.10) | 2.44 (2.05 to 2.90) *** | 0.02 (1028 / 45289.35) | 0.01 (524 / 45861.38) | 2.01 (1.71 to 2.36) *** | 0.02 (1003 / 43420.88) | 0.02 (712 / 44120.85) | 1.41 (1.22 to 1.64) *** |
| > Tachycardia | 0.03 (1533 / 43888.32) | 0.01 (571 / 44346.21) | 2.74 (2.37 to 3.16) *** | 0.04 (1564 / 43888.41) | 0.01 (605 / 44668.55) | 2.62 (2.27 to 3.02) *** | 0.04 (1502 / 41594.79) | 0.02 (997 / 42317.58) | 1.52 (1.34 to 1.72) *** |
| > POTs | 0.009 (419 / 46565.70) | 0.004 (185 / 46704.19) | 2.48 (1.89 to 3.25) *** | 0.009 (434 / 46541.80) | 0.005 (225 / 46987.87) | 1.89 (1.48 to 2.42) *** | 0.010 (432 / 44815.15) | 0.007 (340 / 45493.60) | 1.27 (1.01 to 1.59) * |
| Congestive heart failure | 0.007 (319 / 46556.83) | 0.003 (128 / 46663.40) | 2.62 (1.91 to 3.59) *** | 0.007 (321 / 46559.09) | 0.003 (145 / 46965.40) | 2.30 (1.70 to 3.10) *** | 0.007 (325 / 44776.02) | 0.007 (322 / 45396.49) | 0.98 (0.77 to 1.25) |
| Cardiomyopathy | 0.007 (334 / 46578.69) | 0.002 (104 / 46711.85) | 3.27 (2.32 to 4.62) *** | 0.007 (346 / 46569.49) | 0.003 (129 / 46981.84) | 2.59 (1.89 to 3.55) *** | 0.007 (318 / 44840.13) | 0.006 (283 / 45507.81) | 1.15 (0.89 to 1.48) |
| Myocarditis | 0.002 (75 / 47407.34) | 0.000 (4 / 47425.45) | 20.97 (3.88 to 113.30) *** | 0.002 (70 / 47405.34) | 0.000 (2 / 47734.24) | 42.17 (3.99 to 445.92) *** | 0.001 (66 / 45789.39) | 0.000 (15 / 46463.31) | 4.41 (1.86 to 10.46) *** |
| Coronary disease | 0.007 (311 / 46564.60) | 0.003 (147 / 46663.40) | 2.23 (1.64 to 3.03) *** | 0.007 (312 / 46545.49) | 0.004 (170 / 46944.96) | 1.86 (1.39 to 2.47) *** | 0.007 (306 / 44780.04) | 0.007 (304 / 45406.27) | 1.00 (0.78 to 1.28) |

| | | | | | | | | | |
|---|---|---|---|---|---|---|---|---|---|
| > Myocardial infarction | 0.004 (202 / 46853.39) | 0.002 (84 / 46912.81) | 2.45 (1.65 to 3.65) *** | 0.004 (204 / 46846.38) | 0.002 (107 / 47218.00) | 1.90 (1.32 to 2.73) *** | 0.004 (197 / 45130.20) | 0.004 (167 / 45768.08) | 1.14 (0.83 to 1.58) |
| > Acute coronary syndrome | 0.006 (292 / 46608.70) | 0.003 (139 / 46699.04) | 2.20 (1.60 to 3.02) *** | 0.006 (296 / 46592.34) | 0.004 (164 / 46987.89) | 1.81 (1.35 to 2.42) *** | 0.006 (286 / 44837.40) | 0.006 (281 / 45469.07) | 1.02 (0.79 to 1.32) |
| > Cardiogenic shock | 0.001 (38 / 47372.05) | 0.000 (9 / 47397.15) | 4.49 (1.42 to 14.23) *** | 0.001 (39 / 47373.87) | 0.000 (16 / 47709.36) | 2.51 (1.00 to 6.30) * | 0.001 (37 / 45732.29) | 0.001 (37 / 46400.81) | 0.94 (0.46 to 1.91) |
| Hypercoagulability DVT PE | 0.02 (877 / 45620.93) | 0.006 (288 / 45897.10) | 3.13 (2.55 to 3.84) *** | 0.02 (904 / 45599.57) | 0.007 (304 / 46166.21) | 2.95 (2.42 to 3.61) *** | 0.02 (876 / 43685.19) | 0.01 (595 / 44390.44) | 1.45 (1.23 to 1.70) *** |
| > Hypercoagulability | 0.02 (731 / 45939.24) | 0.005 (253 / 46179.03) | 3.00 (2.40 to 3.74) *** | 0.02 (764 / 45918.21) | 0.006 (265 / 46458.84) | 2.85 (2.30 to 3.54) *** | 0.02 (740 / 44122.84) | 0.01 (539 / 44836.13) | 1.33 (1.12 to 1.59) *** |
| > Deep vein thrombosis | 0.01 (503 / 46522.07) | 0.003 (146 / 46680.61) | 3.50 (2.63 to 4.65) *** | 0.01 (519 / 46492.32) | 0.003 (146 / 46962.45) | 3.46 (2.60 to 4.59) *** | 0.01 (500 / 44737.35) | 0.008 (352 / 45438.00) | 1.37 (1.11 to 1.69) *** |
| > Pulmonary embolism | 0.006 (305 / 46844.11) | 0.002 (85 / 46953.38) | 3.57 (2.46 to 5.18) *** | 0.006 (305 / 46844.01) | 0.002 (85 / 47239.24) | 3.63 (2.50 to 5.26) *** | 0.007 (298 / 45063.50) | 0.004 (191 / 45737.92) | 1.55 (1.17 to 2.06) *** |
| Stroke | 0.007 (331 / 46649.92) | 0.003 (141 / 46740.95) | 2.46 (1.81 to 3.35) *** | 0.007 (327 / 46659.48) | 0.004 (188 / 47060.20) | 1.82 (1.38 to 2.40) *** | 0.007 (334 / 44933.51) | 0.005 (241 / 45591.04) | 1.32 (1.02 to 1.71) * |
| > Ischemic stroke | 0.006 (286 / 46777.38) | 0.003 (120 / 46860.48) | 2.58 (1.84 to 3.60) *** | 0.006 (283 / 46788.71) | 0.003 (152 / 47183.87) | 1.97 (1.45 to 2.66) *** | 0.006 (290 / 45091.61) | 0.005 (213 / 45752.38) | 1.29 (0.98 to 1.70) |
| > Hemorrhagic stroke | 0.002 (90 / 47237.81) | 0.001 (36 / 47274.08) | 2.59 (1.41 to 4.74) *** | 0.002 (92 / 47239.13) | 0.001 (50 / 47579.10) | 1.87 (1.09 to 3.22) ** | 0.002 (88 / 45577.50) | 0.001 (63 / 46249.82) | 1.37 (0.82 to 2.28) |
| **Mental Health** | | | | | | | | | |
| Mental Health Diagnosis | 0.11 (3645 / 32736.86) | 0.08 (2670 / 33085.31) | 1.39 (1.30 to 1.50) *** | 0.11 (3691 / 32839.69) | 0.08 (2550 / 33484.42) | 1.46 (1.36 to 1.57) *** | 0.11 (3587 / 31239.04) | 0.10 (3054 / 31741.36) | 1.17 (1.09 to 1.25) *** |
| > Anxiety | 0.09 (3339 / 35659.08) | 0.07 (2406 / 35993.77) | 1.42 (1.32 to 1.53) *** | 0.10 (3397 / 35753.90) | 0.06 (2239 / 36436.62) | 1.54 (1.42 to 1.66) *** | 0.10 (3289 / 34227.73) | 0.08 (2659 / 34852.59) | 1.24 (1.15 to 1.34) *** |
| > Depression | 0.05 (1935 / 40289.18) | 0.04 (1556 / 40395.20) | 1.27 (1.15 to 1.40) *** | 0.05 (1954 / 40330.06) | 0.04 (1636 / 40697.83) | 1.21 (1.10 to 1.34) *** | 0.05 (1926 / 38804.30) | 0.05 (1936 / 39265.49) | 1.00 (0.90 to 1.10) |
| > PTSD | 0.007 (313 / 46616.36) | 0.005 (216 / 46671.27) | 1.42 (1.09 to 1.86) *** | 0.006 (302 / 46618.53) | 0.005 (228 / 46961.04) | 1.37 (1.05 to 1.79) ** | 0.007 (304 / 45043.43) | 0.005 (222 / 45715.31) | 1.37 (1.04 to 1.79) ** |
| **Neurologic** | | | | | | | | | |
| Amnesia/Memory Difficulty | 0.02 (1037 / 45562.64) | 0.009 (434 / 45781.27) | 2.48 (2.08 to 2.95) *** | 0.02 (1056 / 45553.90) | 0.01 (483 / 46065.83) | 2.18 (1.85 to 2.58) *** | 0.02 (1043 / 43756.61) | 0.01 (653 / 44434.07) | 1.55 (1.33 to 1.80) *** |
| Migraine | 0.02 (1030 / 43668.58) | 0.02 (788 / 43848.08) | 1.29 (1.12 to 1.48) *** | 0.02 (1009 / 43628.85) | 0.02 (840 / 44110.31) | 1.20 (1.05 to 1.38) *** | 0.02 (998 / 41977.65) | 0.02 (1039 / 42616.30) | 0.98 (0.85 to 1.12) |
| Peripheral neuropathy | 0.009 (436 / 46696.85) | 0.004 (204 / 46799.54) | 2.05 (1.59 to 2.65) *** | 0.009 (412 / 46690.33) | 0.005 (231 / 47110.58) | 1.87 (1.46 to 2.40) *** | 0.009 (427 / 45060.75) | 0.006 (282 / 45765.41) | 1.47 (1.16 to 1.87) *** |
| Encephalopathy | 0.007 (341 / 46929.65) | 0.001 (52 / 47042.60) | 6.26 (4.02 to 9.76) *** | 0.007 (331 / 46932.16) | 0.002 (96 / 47344.77) | 3.66 (2.57 to 5.22) *** | 0.007 (329 / 45170.58) | 0.004 (199 / 45829.85) | 1.70 (1.29 to 2.24) *** |

| | | | | | | | | | |
|---|---|---|---|---|---|---|---|---|---|
| Seizure | 0.004 (205 / 46726.88) | 0.002 (114 / 46794.54) | 1.82 (1.28 to 2.58) *** | 0.004 (197 / 46736.02) | 0.003 (137 / 47103.79) | 1.55 (1.11 to 2.16) *** | 0.004 (202 / 45078.09) | 0.004 (188 / 45748.68) | 1.09 (0.80 to 1.49) |
| Dementia | 0.001 (59 / 47358.31) | 0.000 (18 / 47394.42) | 3.21 (1.43 to 7.23) *** | 0.001 (51 / 47360.32) | 0.001 (24 / 47700.96) | 2.29 (1.12 to 4.69) ** | 0.001 (61 / 45742.28) | 0.001 (51 / 46418.87) | 1.25 (0.67 to 2.33) |
| GuillianBarre Syndrome | 0.000 (18 / 47425.05) | 0.000 (5 / 47442.76) | 3.36 (0.72 to 15.64) | 0.000 (17 / 47424.78) | 0.000 (3 / 47758.85) | 4.02 (0.77 to 20.93) | 0.000 (17 / 45819.84) | 0.000 (7 / 46494.72) | 3.07 (0.63 to 14.97) |
| Alzheimer | 0.000 (11 / 47425.22) | 0.000 (7 / 47442.69) | 1.57 (0.32 to 7.73) | 0.000 (8 / 47425.10) | 0.000 (5 / 47756.12) | 1.84 (0.35 to 9.82) | 0.000 (6 / 45820.12) | 0.000 (6 / 46495.42) | 1.04 (0.20 to 5.38) |
| **Diabetes** | | | | | | | | | |
| Type 2 Diabetes | 0.03 (1237 / 37085.27) | 0.02 (649 / 37875.12) | 1.83 (1.60 to 2.10) *** | 0.03 (1181 / 37033.66) | 0.02 (693 / 38099.10) | 1.80 (1.57 to 2.06) *** | 0.03 (1188 / 35400.00) | 0.02 (858 / 36103.69) | 1.39 (1.22 to 1.58) *** |
| **Liver** | | | | | | | | | |
| Liver Test Abnormality | 0.04 (1565 / 43543.52) | 0.02 (929 / 43932.35) | 1.72 (1.52 to 1.94) *** | 0.04 (1603 / 43504.37) | 0.02 (944 / 44159.93) | 1.70 (1.51 to 1.92) *** | 0.04 (1579 / 41670.68) | 0.03 (1307 / 42324.13) | 1.20 (1.07 to 1.34) *** |
| **Nephrology** | | | | | | | | | |
| Kidney Injury | 0.02 (696 / 44579.79) | 0.009 (425 / 44834.57) | 1.70 (1.42 to 2.04) *** | 0.02 (713 / 44567.47) | 0.01 (472 / 45134.13) | 1.58 (1.32 to 1.89) *** | 0.02 (700 / 42635.07) | 0.01 (607 / 43209.05) | 1.18 (0.99 to 1.39) . |
| > Chronic kidney disease | 0.01 (506 / 45588.98) | 0.007 (330 / 45770.03) | 1.60 (1.29 to 1.97) *** | 0.01 (530 / 45574.28) | 0.007 (331 / 46068.04) | 1.61 (1.31 to 2.00) *** | 0.01 (504 / 43903.56) | 0.009 (422 / 44562.96) | 1.24 (1.01 to 1.52) * |
| > Acute kidney injury | 0.01 (546 / 45776.15) | 0.004 (206 / 45977.32) | 2.73 (2.14 to 3.50) *** | 0.01 (547 / 45766.79) | 0.006 (269 / 46285.69) | 2.16 (1.72 to 2.71) *** | 0.01 (539 / 43775.96) | 0.009 (419 / 44362.22) | 1.28 (1.05 to 1.56) ** |
| **Respiratory** | | | | | | | | | |
| Respiratory Failure | 0.007 (323 / 44802.53) | 0.002 (92 / 45196.85) | 3.71 (2.61 to 5.27) *** | 0.007 (326 / 44810.45) | 0.003 (138 / 45477.42) | 2.48 (1.83 to 3.35) *** | 0.007 (317 / 42462.99) | 0.008 (353 / 42967.36) | 0.88 (0.69 to 1.11) |
| > Acute respiratory failure | 0.007 (313 / 44860.78) | 0.002 (82 / 45246.12) | 3.94 (2.73 to 5.70) *** | 0.007 (314 / 44872.80) | 0.003 (132 / 45538.50) | 2.48 (1.82 to 3.38) *** | 0.007 (301 / 42542.84) | 0.008 (331 / 43049.51) | 0.90 (0.70 to 1.15) |
| > Chronic respiratory failure | 0.005 (260 / 47198.41) | 0.001 (22 / 47295.36) | 12.85 (6.39 to 25.83) *** | 0.006 (262 / 47197.90) | 0.001 (23 / 47597.62) | 11.48 (5.92 to 22.24) *** | 0.006 (268 / 45517.10) | 0.004 (160 / 46180.36) | 1.63 (1.19 to 2.22) *** |
| Interstitial lung disease | 0.009 (412 / 46928.47) | 0.001 (53 / 47054.30) | 7.71 (4.92 to 12.08) *** | 0.009 (406 / 46929.76) | 0.002 (80 / 47370.44) | 5.03 (3.47 to 7.31) *** | 0.009 (391 / 45147.01) | 0.007 (319 / 45828.01) | 1.22 (0.97 to 1.54) |
| **Dermatologic** | | | | | | | | | |
| Atopic dermatitis | 0.009 (440 / 46253.29) | 0.008 (384 / 46310.86) | 1.24 (1.01 to 1.54) * | 0.01 (462 / 46246.41) | 0.009 (398 / 46638.83) | 1.16 (0.94 to 1.43) | 0.01 (446 / 44669.62) | 0.009 (390 / 45347.79) | 1.16 (0.93 to 1.43) |
| Urticaria | 0.009 (402 / 46402.65) | 0.006 (286 / 46454.38) | 1.44 (1.14 to 1.83) *** | 0.008 (391 / 46355.53) | 0.008 (356 / 46725.14) | 1.19 (0.95 to 1.49) | 0.009 (383 / 44740.90) | 0.01 (487 / 45385.93) | 0.79 (0.64 to 0.98) * |
| Herpesviral vesicular dermatitis | 0.003 (145 / 46948.42) | 0.002 (108 / 46977.87) | 1.30 (0.89 to 1.90) | 0.003 (144 / 46943.25) | 0.003 (150 / 47272.60) | 0.96 (0.68 to 1.36) | 0.003 (138 / 45353.41) | 0.004 (160 / 46017.56) | 0.86 (0.60 to 1.22) |
| **Others** | | | | | | | | | |

| | | | | | | | | | |
|---|---|---|---|---|---|---|---|---|---|
| Sleep apnea | 0.04 (1549 / 41676.21) | 0.02 (891 / 42016.58) | 1.78 (1.58 to 2.02) *** | 0.04 (1555 / 41663.30) | 0.02 (961 / 42299.37) | 1.60 (1.42 to 1.81) *** | 0.04 (1547 / 39876.21) | 0.04 (1633 / 40393.38) | 0.93 (0.84 to 1.03) |

† Denominator for incidence of each outcome is person-time for individuals at-risk starting 21 days after index date. 2 decimals are kept for incidence rates >= 0.01 and 3 decimals are kept for incidence rates < 0.01.

†† Confidence intervals are calculated based on Bonferroni correction, i.e., (1 - 0.05/N) * 100 confidence interval.

††† Individuals were excluded if they had less than index date + 21 days of follow-up, had already been diagnosed with outcome of interest prior to index date + 21 days or their matched pair was excluded for the same reasons.

†††† The outcome observation periods length (in days) is measured from time origin (index date + 21 days) to end of observation of the member.

*** p value < 0.001

** p value < 0.01

* p value < 0.05

### Table 2d: Rates and Hazard Ratios for Symptom Sequelae by SARS-CoV-2 Infection and Comparison Groups, UnitedHealth Group Clinical Discovery Database through October 31, 2020.

| **Outcome** | **2020 SARS-CoV-2 Infected: incidence rate (case/ person-year)** | **2020 Comparator: incidence rate (case/ person-year) †** | **Hazard Ratio (95 CI ††)** | **2019-matched 2020 SARS-CoV-2 Infected: incidence rate (case/ person-year)** | **2019 Comparator: incidence rate (case/ person-year) †** | **Hazard Ratio (95 CI ††)** | **vLRTI-matched 2020 SARS-CoV-2 Infected: incidence rate (case/ person-year)** | **vLRTI comparator: incidence rate (case/ person-year) †** | **Hazard Ratio (95 CI ††)** |
|---|---|---|---|---|---|---|---|---|---|
| **Matched Group Size** | **193113 †††** | **193113 †††** | | **193264 †††** | **193264 †††** | | **181613 †††** | **181613 †††** | |
| **Follow-up period (days): median (IQR) ††††** | **87 (45 – 124)** | **87 (45 – 124)** | | **87 (45 – 124)** | **87 (46 – 124)** | | **89 (45 – 135)** | **87 (43 – 130)** | |
| Fatigue Diagnosis | 0.15 (5177 / 34590.87) | 0.07 (2584 / 35579.05) | 2.11 (1.97 to 2.26) *** | 0.15 (5160 / 34327.11) | 0.07 (2594 / 35605.32) | 2.06 (1.93 to 2.21) *** | 0.15 (4875 / 31464.09) | 0.11 (3638 / 32246.65) | 1.36 (1.27 to 1.44) *** |
| > Fatigue | 0.15 (5062 / 34689.62) | 0.07 (2581 / 35641.79) | 2.06 (1.93 to 2.21) *** | 0.15 (5037 / 34431.62) | 0.07 (2585 / 35671.77) | 2.02 (1.88 to 2.16) *** | 0.15 (4763 / 31582.42) | 0.11 (3632 / 32336.83) | 1.33 (1.25 to 1.41) *** |
| > Chronic Fatigue | 0.01 (536 / 46341.01) | 0.007 (305 / 46426.64) | 1.78 (1.43 to 2.22) *** | 0.01 (546 / 46313.92) | 0.007 (339 / 46741.97) | 1.60 (1.30 to 1.98) *** | 0.01 (521 / 44685.73) | 0.010 (441 / 45339.92) | 1.17 (0.95 to 1.43) |
| > Postviral fatigue | 0.007 (343 / 47241.23) | 0.000 (13 / 47340.71) | 25.65 (11.09 to 59.29) *** | 0.007 (354 / 47237.29) | 0.000 (20 / 47650.36) | 19.65 (9.40 to 41.09) *** | 0.007 (344 / 45604.65) | 0.001 (45 / 46346.09) | 7.46 (4.62 to 12.04) *** |
| Myalgia | 0.14 (4609 / 33766.63) | 0.09 (3044 / 34620.02) | 1.55 (1.45 to 1.66) *** | 0.14 (4565 / 33401.85) | 0.10 (3464 / 34415.83) | 1.36 (1.28 to 1.45) *** | 0.14 (4311 / 30523.68) | 0.13 (4079 / 31114.77) | 1.06 (1.00 to 1.13) |
| Anosmia | 0.007 (323 / 46633.43) | 0.001 (52 / 46710.06) | 5.37 (3.47 to 8.30) *** | 0.006 (299 / 46651.89) | 0.001 (26 / 47059.87) | 13.12 (6.87 to 25.05) *** | 0.007 (300 / 45069.29) | 0.001 (51 / 45838.62) | 5.78 (3.66 to 9.12) *** |

† Denominator for incidence of each outcome is person-time for individuals at-risk starting 21 days after index date for outcome of interest. 2 decimals are kept for incidence rates >= 0.01 and 3 decimals are kept for incidence rates < 0.01.

†† Confidence intervals are calculated based on Bonferroni correction, i.e., (1 - 0.05/N) * 100 confidence interval.

††† Individuals were excluded if they had less than index date + 21 days of follow-up, had already been diagnosed with outcome of interest prior to index date + 21 days or their matched pair was excluded for the same reasons.

†††† The outcome observation periods length (in days) is measured from time origin (which is index date + 21 days) to end of observation of the member.

*** p value < 0.001

** p value < 0.01

* p value < 0.05

**Table 3: Rates and Hazard Ratios for New Clinical and Symptom Sequelae by Month of Follow-up, UnitedHealth Group Clinical Discovery Database through October 31, 2020.**

| **Clinical Sequelae** | **Months of Follow-up*** | | | | | | |
|---|---|---|---|---|---|---|---|
| | -1 | 1 (include index date) | 2 | 3 | 4 | 5 | 6 |
| **Hypertension** | | | | | | | |
| SARS-CoV-2 Infection | 1296 / 159380 | 3093 / 157402 | 683 / 99192 | 363 / 66933 | 195 / 41347 | 77 / 17484 | 43 / 5775 |
| 2020 comparator | 691 / 159380 | 571 / 157402 | 393 / 99192 | 262 / 66933 | 135 / 41347 | 59 / 17484 | 24 / 5775 |
| HR (95 CI) | 1.88 (1.71 to 2.06) | 5.48 (5.01 to 5.99) | 1.75 (1.54 to 1.98) | 1.40 (1.19 to 1.64) | 1.46 (1.17 to 1.81) | 1.34 (0.95 to 1.88) | 1.81 (1.10 to 2.96) |
| **Arrhythmia+** | | | | | | | |
| SARS-CoV-2 Infection | 1353 / 250328 | 5571 / 248546 | 642 / 156989 | 369 / 107416 | 170 / 66983 | 67 / 28212 | 26 / 9519 |
| 2020 comparator | 429 / 250328 | 397 / 248546 | 219 / 156989 | 172 / 107416 | 87 / 66983 | 31 / 28212 | 14 / 9519 |
| HR (95 CI) | 3.16 (2.83 to 3.52) | 14.20 (12.82 to 15.71) | 2.94 (2.52 to 3.43) | 2.15 (1.80 to 2.58) | 1.96 (1.51 to 2.54) | 2.17 (1.42 to 3.32) | 1.87 (0.98 to 3.59) |
| **Cardiomyopathy +** | | | | | | | |
| SARS-CoV-2 Infection | 123 / 263126 | 644 / 262916 | 96 / 170207 | 53 / 117220 | 36 / 73578 | 10 / 31293 | 3 / 10732 |
| 2020 comparator | 87 / 263126 | 57 / 262916 | 33 / 170207 | 17 / 117220 | 10 / 73578 | 5 / 31293 | 1 / 10732 |
| HR (95 CI) | 1.41 (1.07 to 1.86) | 11.31 (8.63 to 14.82) | 2.91 (1.96 to 4.32) | 3.12 (1.81 to 5.39) | 3.60 (1.79 to 7.26) | 1.99 (0.68 to 5.84) | 3.01 (0.31 to 28.94) |
| **Congestive heart failure** | | | | | | | |
| SARS-CoV-2 Infection | 177 / 263399 | 897 / 263160 | 107 / 170214 | 50 / 117157 | 32 / 73506 | 9 / 31320 | 6 / 10739 |

| | | | | | | | |
|---|---|---|---|---|---|---|---|
| 2020 comparator | 62 / 263399 | 45 / 263160 | 45 / 170214 | 24 / 117157 | 18 / 73506 | 3 / 31320 | 0 / 10739 |
| HR (95 CI) | 2.85 (2.14 to 3.81) | 19.96 (14.80 to 26.93) | 2.38 (1.68 to 3.37) | 2.09 (1.28 to 3.39) | 1.78 (1.00 to 3.17) | 2.99 (0.81 to 11.05) | |
| **Coronary disease** | | | | | | | |
| SARS-CoV-2 Infection | 319 / 263985 | 1278 / 263604 | 94 / 170221 | 62 / 117201 | 29 / 73575 | 11 / 31307 | 3 / 10721 |
| 2020 comparator | 62 / 263985 | 50 / 263604 | 39 / 170221 | 28 / 117201 | 15 / 73575 | 8 / 31307 | 4 / 10721 |
| HR (95 CI) | 5.15 (3.92 to 6.76) | 25.61 (19.31 to 33.98) | 2.41 (1.66 to 3.50) | 2.22 (1.42 to 3.46) | 1.94 (1.04 to 3.61) | 1.37 (0.55 to 3.40) | 0.75 (0.17 to 3.36) |
| **Hypercoagulability DVT PE+** | | | | | | | |
| SARS-CoV-2 Infection | 427 / 260709 | 2110 / 260137 | 299 / 167328 | 147 / 115032 | 52 / 72067 | 28 / 30630 | 6 / 10446 |
| 2020 comparator | 145 / 260709 | 119 / 260137 | 65 / 167328 | 57 / 115032 | 29 / 72067 | 9 / 30630 | 8 / 10446 |
| HR (95 CI) | 2.95 (2.44 to 3.56) | 17.80 (14.79 to 21.41) | 4.61 (3.52 to 6.03) | 2.58 (1.90 to 3.51) | 1.79 (1.14 to 2.83) | 3.11 (1.47 to 6.59) | 0.76 (0.26 to 2.18) |
| **Mental Health Diagnosis+** | | | | | | | |
| SARS-CoV-2 Infection | 1943 / 194808 | 3380 / 191487 | 1134 / 121180 | 654 / 82379 | 346 / 51086 | 118 / 21543 | 49 / 7363 |
| 2020 comparator | 1400 / 194808 | 1238 / 191487 | 746 / 121180 | 474 / 82379 | 299 / 51086 | 121 / 21543 | 36 / 7363 |
| HR (95 CI) | 1.39 (1.30 to 1.49) | 2.75 (2.58 to 2.93) | 1.52 (1.39 to 1.67) | 1.38 (1.23 to 1.55) | 1.16 (0.99 to 1.35) | 0.98 (0.76 to 1.26) | 1.37 (0.89 to 2.11) |
| **Amnesia/ Memory Difficulty** | | | | | | | |
| SARS-CoV-2 Infection | 471 / 259801 | 1608 / 259151 | 341 / 167024 | 159 / 114785 | 88 / 71891 | 21 / 30552 | 12 / 10417 |
| 2020 comparator | 179 / 259801 | 182 / 259151 | 109 / 167024 | 69 / 114785 | 57 / 71891 | 25 / 30552 | 5 / 10417 |

| | | | | | | | |
|---|---|---|---|---|---|---|---|
| HR (95 CI) | 2.63 (2.22 to 3.13) | 8.86 (7.60 to 10.33) | 3.13 (2.52 to 3.89) | 2.31 (1.74 to 3.06) | 1.55 (1.11 to 2.16) | 0.84 (0.47 to 1.50) | 2.42 (0.85 to 6.86) |
| **Migraine** | | | | | | | |
| SARS-CoV-2 Infection | 511 / 248797 | 689 / 247901 | 291 / 160128 | 178 / 109937 | 109 / 68737 | 45 / 29253 | 16 / 9987 |
| 2020 comparator | 386 / 248797 | 296 / 247901 | 231 / 160128 | 138 / 109937 | 98 / 68737 | 21 / 29253 | Aug-87 |
| HR (95 CI) | 1.32 (1.16 to 1.51) | 2.33 (2.03 to 2.67) | 1.26 (1.06 to 1.50) | 1.29 (1.03 to 1.61) | 1.11 (0.85 to 1.46) | 2.14 (1.27 to 3.59) | 2.01 (0.86 to 4.70) |
| **Peripheral neuropathy** | | | | | | | |
| SARS-CoV-2 Infection | 122 / 263493 | 177 / 263289 | 109 / 170751 | 79 / 117552 | 55 / 73711 | 21 / 31404 | 6 / 10767 |
| 2020 comparator | 83 / 263493 | 88 / 263289 | 49 / 170751 | 36 / 117552 | 28 / 73711 | 10 / 31404 | 1 / 10767 |
| HR (95 CI) | 1.47 (1.11 to 1.94) | 2.01 (1.56 to 2.60) | 2.23 (1.59 to 3.12) | 2.20 (1.48 to 3.26) | 1.97 (1.25 to 3.10) | 2.09 (0.98 to 4.44) | 6.04 (0.72 to 50.21) |
| **Encephalopathy** | | | | | | | |
| SARS-CoV-2 Infection group | 155 / 265385 | 921 / 265192 | 122 / 171556 | 59 / 118144 | 26 / 74135 | 6 / 31550 | 3 / 10788 |
| 2020 comparator group | 38 / 265385 | 24 / 265192 | 18 / 171556 | 10 / 118144 | 4 / 74135 | 2 / 31550 | 1 / 10788 |
| HR (95 CI) | 4.08 (2.86 to 5.82) | 38.42 (25.62 to 57.62) | 6.78 (4.13 to 11.13) | 5.91 (3.02 to 11.55) | 6.51 (2.27 to 18.64) | 2.99 (0.60 to 14.81) | 3.03 (0.31 to 29.16) |
| **Type 2 Diabetes** | | | | | | | |
| SARS-CoV-2 Infection | 536 / 215755 | 1629 / 214943 | 346 / 137676 | 204 / 93927 | 114 / 58694 | 54 / 24942 | 22 / 8444 |
| 2020 comparator | 278 / 215755 | 280 / 214943 | 159 / 137676 | 110 / 93927 | 74 / 58694 | 37 / 24942 | Sep-44 |
| HR (95 CI) | 1.93 (1.67 to 2.23) | 5.85 (5.15 to 6.64) | 2.19 (1.81 to 2.64) | 1.86 (1.48 to 2.35) | 1.55 (1.16 to 2.08) | 1.47 (0.97 to 2.24) | 2.47 (1.14 to 5.38) |

| | | | | | | | |
|---|---|---|---|---|---|---|---|
| **Liver Test Abnormality** | | | | | | | |
| SARS-CoV-2 Infection | 832 / 250678 | 1763 / 249453 | 469 / 160302 | 252 / 109852 | 168 / 68584 | 70 / 29063 | 16 / 9905 |
| 2020 comparator | 394 / 250678 | 329 / 249453 | 274 / 160302 | 161 / 109852 | 88 / 68584 | 46 / 29063 | 16 / 9905 |
| HR (95 CI) | 2.11 (1.87 to 2.38) | 5.38 (4.78 to 6.05) | 1.72 (1.48 to 1.99) | 1.57 (1.29 to 1.91) | 1.91 (1.48 to 2.48) | 1.52 (1.05 to 2.20) | 1.01 (0.50 to 2.02) |
| **Kidney Injury+** | | | | | | | |
| SARS-CoV-2 Infection | 593 / 256325 | 3096 / 255535 | 210 / 163540 | 120 / 112403 | 73 / 70436 | 22 / 29943 | 10 / 10221 |
| 2020 comparator | 199 / 256325 | 199 / 255535 | 97 / 163540 | 87 / 112403 | 42 / 70436 | 19 / 29943 | 4 / 10221 |
| HR (95 CI) | 2.98 (2.54 to 3.50) | 15.65 (13.56 to 18.07) | 2.17 (1.70 to 2.76) | 1.38 (1.05 to 1.82) | 1.74 (1.19 to 2.54) | 1.16 (0.64 to 2.11) | 2.52 (0.79 to 8.01) |
| **Respiratory failure+** | | | | | | | |
| SARS-CoV-2 Infection | 614 / 264151 | 9259 / 263477 | 102 / 164808 | 48 / 113354 | 31 / 70990 | 8 / 30114 | 4 / 10178 |
| 2020 comparator | 60 / 264151 | 31 / 263477 | 22 / 164808 | 13 / 113354 | 11 / 70990 | 4 / 30114 | 3 / 10178 |
| HR (95 CI) | 10.24 (7.86 to 13.35) | 304.05 (213.72 to 432.54) | 4.64 (2.93 to 7.36) | 3.70 (2.01 to 6.83) | 2.83 (1.42 to 5.62) | 2.00 (0.60 to 6.65) | 1.35 (0.30 to 6.02) |
| **Interstitial lung disease** | | | | | | | |
| SARS-CoV-2 Infection | 115 / 265094 | 575 / 264948 | 127 / 171619 | 72 / 118144 | 41 / 74118 | 14 / 31543 | 7 / 10806 |
| 2020 comparator | 31 / 265094 | 34 / 264948 | 17 / 171619 | 12 / 118144 | 5 / 74118 | 2 / 31543 | 1 / 10806 |
| HR (95 CI) | 3.71 (2.50 to 5.52) | 16.92 (11.97 to 23.91) | 7.48 (4.51 to 12.40) | 6.01 (3.26 to 11.07) | 8.21 (3.24 to 20.77) | 6.97 (1.58 to 30.69) | 7.06 (0.87 to 57.30) |

| | | | | | | | |
|---|---|---|---|---|---|---|---|
| **Atopic dermatitis** | | | | | | | |
| SARS-CoV-2 Infection | 183 / 261197 | 173 / 260842 | 101 / 169096 | 86 / 116369 | 49 / 72975 | 18 / 31045 | 5 / 10628 |
| 2020 comparator | 172 / 261197 | 149 / 260842 | 107 / 169096 | 80 / 116369 | 42 / 72975 | 16 / 31045 | 7 / 10628 |
| HR (95 CI) | 1.06 (0.86 to 1.31) | 1.16 (0.93 to 1.44) | 0.94 (0.72 to 1.24) | 1.08 (0.79 to 1.46) | 1.17 (0.77 to 1.76) | 1.12 (0.57 to 2.19) | 0.72 (0.23 to 2.27) |
| **Urticaria** | | | | | | | |
| SARS-CoV-2 Infection | 200 / 262026 | 279 / 261699 | 111 / 169601 | 79 / 116727 | 53 / 73163 | 9 / 31077 | 6 / 10642 |
| 2020 comparator | 127 / 262026 | 123 / 261699 | 75 / 169601 | 65 / 116727 | 30 / 73163 | 10 / 31077 | 5 / 10642 |
| HR (95 CI) | 1.57 (1.26 to 1.97) | 2.27 (1.83 to 2.80) | 1.48 (1.10 to 1.99) | 1.22 (0.88 to 1.69) | 1.77 (1.13 to 2.77) | 0.90 (0.36 to 2.21) | 1.20 (0.37 to 3.95) |
| **Sleep apnea** | | | | | | | |
| SARS-CoV-2 Infection | 548 / 238638 | 910 / 237669 | 482 / 153466 | 259 / 105105 | 156 / 65652 | 59 / 27826 | 32 / 9471 |
| 2020 comparator | 422 / 238638 | 344 / 237669 | 246 / 153466 | 165 / 105105 | 97 / 65652 | 32 / 27826 | 14 / 9471 |
| HR (95 CI) | 1.30 (1.14 to 1.48) | 2.65 (2.34 to 3.00) | 1.96 (1.68 to 2.29) | 1.57 (1.30 to 1.91) | 1.61 (1.25 to 2.08) | 1.85 (1.20 to 2.84) | 2.31 (1.23 to 4.32) |
| **Fatigue Diagnosis+** | | | | | | | |
| SARS-CoV-2 Infection | 4401 / 219007 | 10576 / 213409 | 1640 / 130171 | 818 / 87610 | 480 / 53897 | 201 / 22624 | 78 / 7555 |
| 2020 comparator | 1215 / 219007 | 1067 / 213409 | 722 / 130171 | 487 / 87610 | 269 / 53897 | 101 / 22624 | 36 / 7555 |
| HR (95 CI) | 3.64 (3.42 to 3.88) | 10.18 (9.56 to 10.83) | 2.29 (2.10 to 2.50) | 1.69 (1.51 to 1.89) | 1.79 (1.54 to 2.08) | 2.00 (1.57 to 2.54) | 2.20 (1.48 to 3.27) |

* For j month follow-up, we only consider members who had at least (j-1)*30 days follow-up period.
* Incidence is reporting number of new cases within jth follow-up month dividing by population at-risk at jth follow-up month.
* The hazard ratio and 95 confidence interval are estimated when considering start of the jth follow-up month as time 0 and end of jth follow-up month as end of observation.

* We only considered members in the interval whose matched pair is also available in the same time interval. This criterion reduced population significantly.
+ Aggregate diagnosis includes all subdiagnoses as listed in eTable 1

**Table 4a: Risk Difference (per 100 individuals) for Clinical Sequelae by Age, UnitedHealth Group Clinical Discovery Database through October 31, 2020.**

| **Clinical Sequelae** | **Category** | **18-34 (†)** | **35-50 (†)** | **>50 (†)** | **p-interaction** |
|---|---|---|---|---|---|
| | **Matched Group Size** | **66425 vs 66425 ††** | **64758 vs 64758 ††** | **64011 vs 64011 ††** | |
| **Hypertension** | | | | | |
| Hypertension | SARS-CoV-2 Group | 0.83 | 2.83 | 5.40 | |
| | 2020 Comparator Group | 0.49 | 1.77 | 3.54 | |
| | Risk Difference | 0.33 (0.04 to 0.55) | 1.06 (0.67 to 1.8) | 1.86 (0.48 to 3.09) | |
| | p-value | <0.001 | <0.001 | <0.001 | <0.001 |
| Pulmonary Hypertension | SARS-CoV-2 Group | 0.03 | 0.10 | 0.29 | |
| | 2020 Comparator Group | 0.01 | 0.04 | 0.09 | |
| | Risk Difference | 0.02 (-0.02 to 0.06) | 0.06 (0 to 0.15) | 0.2 (0.07 to 0.32) | |
| | p-value | 1 | 0.15 | <0.001 | <0.001 |
| **Cardiology** | | | | | |
| Arrythmia | SARS-CoV-2 Group | 0.97 | 1.46 | 2.21 | |
| | 2020 Comparator Group | 0.44 | 0.62 | 1 | |
| | Risk Difference | 0.53 (0.32 to 0.76) | 0.83 (0.62 to 1.23) | 1.21 (0.94 to 1.66) | |
| | p-value | <0.001 | <0.001 | <0.001 | <0.001 |
| > Cardiac Arrythmia | SARS-CoV-2 Group | 0.41 | 0.61 | 1.16 | |
| | 2020 Comparator Group | 0.16 | 0.28 | 0.57 | |
| | Risk Difference | 0.25 (0.09 to 0.4) | 0.34 (0.15 to 0.53) | 0.59 (0.34 to 0.86) | |
| | p-value | <0.001 | <0.001 | <0.001 | <0.001 |
| > Tachycardia | SARS-CoV-2 Group | 0.75 | 1.08 | 1.50 | |
| | 2020 Comparator Group | 0.35 | 0.43 | 0.58 | |
| | Risk Difference | 0.4 (0.25 to 0.63) | 0.65 (0.43 to 0.93) | 0.92 (0.55 to 1.09) | |
| | p-value | <0.001 | <0.001 | <0.001 | <0.001 |
| > POTs | SARS-CoV-2 Group | 0.19 | 0.20 | 0.47 | |
| | 2020 Comparator Group | 0.07 | 0.10 | 0.22 | |
| | Risk Difference | 0.13 (0.03 to 0.25) | 0.1 (-0.04 to 0.17) | 0.25 (0.09 to 0.39) | |
| | p-value | <0.001 | 1 | <0.001 | <0.001 |
| Congestive Heart Failure | SARS-CoV-2 Group | 0.03 | 0.16 | 0.49 | |
| | 2020 Comparator Group | 0 | 0.05 | 0.17 | |
| | Risk Difference | 0.03 (0 to 0.06) | 0.11 (0 to 0.16) | 0.32 (0.2 to 0.48) | |
| | p-value | <0.001 | 0.15 | <0.001 | <0.001 |
| Cardiomyopathy | SARS-CoV-2 Group | 0.07 | 0.17 | 0.43 | |
| | 2020 Comparator Group | 0.01 | 0.05 | 0.17 | |
| | Risk Difference | 0.06 (0.02 to 0.12) | 0.12 (0.01 to 0.18) | 0.26 (0.14 to 0.41) | |

| | | | | | |
|---|---|---|---|---|---|
| | p-value | <0.001 | <0.001 | <0.001 | <0.001 |
| Myocarditis | SARS-CoV-2 Group | 0.09 | 0.02 | 0.03 | |
| | 2020 Comparator Group | 0 | 0 | 0 | |
| | Risk Difference | 0.08 (0.03 to 0.16) | 0.02 (0 to 0.06) | 0.03 (0.01 to 0.1) | |
| | p-value | <0.001 | <0.001 | <0.001 | 0.002 |
| Coronary Disease | SARS-CoV-2 Group | 0.03 | 0.14 | 0.47 | |
| | 2020 Comparator Group | 0.01 | 0.09 | 0.21 | |
| | Risk Difference | 0.02 (-0.01 to 0.06) | 0.05 (-0.02 to 0.16) | 0.25 (0.07 to 0.39) | |
| | p-value | 1 | 1 | <0.001 | <0.001 |
| > Myocardial Infarction | SARS-CoV-2 Group | 0.03 | 0.08 | 0.30 | |
| | 2020 Comparator Group | 0.01 | 0.05 | 0.14 | |
| | Risk Difference | 0.02 (-0.01 to 0.05) | 0.03 (-0.04 to 0.08) | 0.16 (0.01 to 0.29) | |
| | p-value | 1 | 1 | <0.001 | <0.001 |
| > Acute Coronary Syndrome | SARS-CoV-2 Group | 0.03 | 0.13 | 0.44 | |
| | 2020 Comparator Group | 0.01 | 0.08 | 0.21 | |
| | Risk Difference | 0.02 (-0.01 to 0.06) | 0.05 (-0.02 to 0.17) | 0.23 (0.08 to 0.4) | |
| | p-value | 0.61 | 1 | <0.001 | <0.001 |
| > Cardiogenic Shock | SARS-CoV-2 Group | 0 | 0.02 | 0.05 | |
| | 2020 Comparator Group | 0 | 0.01 | 0.01 | |
| | Risk Difference | 0 (0 to 0.03) | 0.01 (-0.04 to 0.04) | 0.03 (-0.02 to 0.09) | |
| | p-value | <0.001 | 1 | 0.76 | 0.32 |
| Hypercoagulability DVT PE | SARS-CoV-2 Group | 0.23 | 0.61 | 1.03 | |
| | 2020 Comparator Group | 0.10 | 0.17 | 0.29 | |
| | Risk Difference | 0.13 (0.03 to 0.27) | 0.44 (0.26 to 0.61) | 0.74 (0.47 to 0.91) | |
| | p-value | <0.001 | <0.001 | <0.001 | <0.001 |
| > Hypercoagulability | SARS-CoV-2 Group | 0.20 | 0.50 | 0.88 | |
| | 2020 Comparator Group | 0.09 | 0.17 | 0.27 | |
| | Risk Difference | 0.11 (0.04 to 0.25) | 0.33 (0.23 to 0.53) | 0.6 (0.37 to 0.77) | |
| | p-value | <0.001 | <0.001 | <0.001 | <0.001 |
| > Deep Vein Thrombosis | SARS-CoV-2 Group | 0.09 | 0.32 | 0.62 | |
| | 2020 Comparator Group | 0.04 | 0.10 | 0.17 | |
| | Risk Difference | 0.05 (-0.02 to 0.11) | 0.23 (0.1 to 0.33) | 0.45 (0.25 to 0.59) | |
| | p-value | 1 | <0.001 | <0.001 | <0.001 |
| > Pulmonary Embolism | SARS-CoV-2 Group | 0.08 | 0.18 | 0.33 | |
| | 2020 Comparator Group | 0.01 | 0.05 | 0.07 | |
| | Risk Difference | 0.07 (0 to 0.12) | 0.13 (0.05 to 0.21) | 0.26 (0.15 to 0.39) | |
| | p-value | <0.001 | <0.001 | <0.001 | <0.001 |
| Stroke | SARS-CoV-2 Group | 0.05 | 0.18 | 0.42 | |

| | | | | | |
|---|---|---|---|---|---|
| | 2020 Comparator Group | 0.02 | 0.08 | 0.19 | |
| | Risk Difference | 0.03 (-0.02 to 0.08) | 0.11 (0.01 to 0.21) | 0.23 (0.12 to 0.41) | |
| | p-value | 1 | <0.001 | <0.001 | <0.001 |
| > Ischemic Stroke | SARS-CoV-2 Group | 0.05 | 0.15 | 0.37 | |
| | 2020 Comparator Group | 0.02 | 0.07 | 0.18 | |
| | Risk Difference | 0.03 (-0.02 to 0.08) | 0.07 (-0.03 to 0.15) | 0.19 (0.08 to 0.37) | |
| | p-value | 1 | 0.92 | <0.001 | <0.001 |
| > Hemorrhagic Stroke | SARS-CoV-2 Group | 0.01 | 0.06 | 0.11 | |
| | 2020 Comparator Group | 0 | 0.01 | 0.03 | |
| | Risk Difference | 0.01 (-0.01 to 0.03) | 0.05 (0.01 to 0.1) | 0.08 (0 to 0.17) | |
| | p-value | 1 | <0.001 | 0.15 | <0.001 |
| **Mental Health** | | | | | |
| Mental Health Diagnosis | SARS-CoV-2 Group | 3.87 | 3.90 | 3.29 | |
| | 2020 Comparator Group | 3.24 | 2.75 | 2.35 | |
| | Risk Difference | 0.63 (0.19 to 1.61) | 1.15 (0.93 to 2.38) | 0.95 (0.47 to 1.58) | |
| | p-value | <0.001 | <0.001 | <0.001 | 0.35 |
| > Anxiety | SARS-CoV-2 Group | 3.34 | 3.29 | 2.63 | |
| | 2020 Comparator Group | 2.81 | 2.29 | 1.83 | |
| | Risk Difference | 0.53 (-0.1 to 1.14) | 1.01 (0.49 to 1.66) | 0.79 (0.35 to 1.43) | |
| | p-value | 0.15 | <0.001 | <0.001 | 0.84 |
| > Depression | SARS-CoV-2 Group | 1.71 | 1.73 | 1.52 | |
| | 2020 Comparator Group | 1.76 | 1.32 | 1.08 | |
| | Risk Difference | -0.05 (-0.62 to 0.32) | 0.41 (-0.01 to 0.69) | 0.44 (0.17 to 0.84) | |
| | p-value | 1 | 0.15 | <0.001 | <0.001 |
| > PTSD | SARS-CoV-2 Group | 0.24 | 0.27 | 0.16 | |
| | 2020 Comparator Group | 0.22 | 0.14 | 0.08 | |
| | Risk Difference | 0.03 (-0.1 to 0.15) | 0.13 (0.01 to 0.25) | 0.08 (-0.02 to 0.15) | |
| | p-value | 1 | <0.001 | 1 | 1 |
| **Neurologic** | | | | | |
| Amnesia/Memory Difficulty | SARS-CoV-2 Group | 0.47 | 0.62 | 1.02 | |
| | 2020 Comparator Group | 0.27 | 0.22 | 0.44 | |
| | Risk Difference | 0.2 (0.13 to 0.5) | 0.4 (0.27 to 0.6) | 0.59 (0.38 to 0.8) | |
| | p-value | <0.001 | <0.001 | <0.001 | <0.001 |
| Migraine | SARS-CoV-2 Group | 0.74 | 0.93 | 0.62 | |
| | 2020 Comparator Group | 0.58 | 0.75 | 0.49 | |
| | Risk Difference | 0.16 (-0.16 to 0.35) | 0.18 (-0.08 to 0.46) | 0.13 (-0.07 to 0.31) | |
| | p-value | 1 | 1 | 1 | 1 |
| Peripheral Neuropathy | SARS-CoV-2 Group | 0.07 | 0.30 | 0.52 | |

| | | | | | |
|---|---|---|---|---|---|
| | 2020 Comparator Group | 0.03 | 0.12 | 0.30 | |
| | Risk Difference | 0.04 (-0.05 to 0.09) | 0.18 (0.08 to 0.31) | 0.22 (0.05 to 0.4) | |
| | p-value | 1 | <0.001 | <0.001 | <0.001 |
| Encephalopathy | SARS-CoV-2 Group | 0.06 | 0.16 | 0.47 | |
| | 2020 Comparator Group | 0.02 | 0.05 | 0.11 | |
| | Risk Difference | 0.03 (-0.02 to 0.1) | 0.11 (0.04 to 0.21) | 0.36 (0.22 to 0.5) | |
| | p-value | 1 | <0.001 | <0.001 | <0.001 |
| Seizure | SARS-CoV-2 Group | 0.09 | 0.16 | 0.17 | |
| | 2020 Comparator Group | 0.06 | 0.08 | 0.08 | |
| | Risk Difference | 0.03 (-0.05 to 0.13) | 0.08 (-0.04 to 0.15) | 0.1 (-0.01 to 0.17) | |
| | p-value | 1 | 1 | 0.46 | 0.13 |
| Dementia | SARS-CoV-2 Group | 0.01 | 0.02 | 0.07 | |
| | 2020 Comparator Group | 0.01 | 0 | 0.03 | |
| | Risk Difference | 0 (-0.02 to 0.02) | 0.02 (0 to 0.05) | 0.05 (0 to 0.1) | |
| | p-value | 1 | 1 | 0.46 | <0.001 |
| GuillianBarre Syndrome | SARS-CoV-2 Group | 0.02 | 0.01 | 0.02 | |
| | 2020 Comparator Group | 0 | 0 | 0 | |
| | Risk Difference | 0.01 (-0.01 to 0.07) | 0.01 (-0.04 to 0.02) | 0.01 (-0.01 to 0.05) | |
| | p-value | 1 | 1 | 1 | 1 |
| Alzheimer | SARS-CoV-2 Group | 0 | 0 | 0.01 | |
| | 2020 Comparator Group | 0 | 0 | 0 | |
| | Risk Difference | 0 (0 to 0) | 0 (-0.01 to 0.03) | 0.01 (0 to 0.04) | |
| | p-value | <0.001 | 1 | <0.001 | 0.05 |
| **Diabetes** | | | | | |
| Type 2 Diabetes | SARS-CoV-2 Group | 0.36 | 1.09 | 1.81 | |
| | 2020 Comparator Group | 0.16 | 0.58 | 1.07 | |
| | Risk Difference | 0.2 (0.05 to 0.32) | 0.51 (0.17 to 0.75) | 0.74 (0.39 to 1.3) | |
| | p-value | <0.001 | <0.001 | <0.001 | <0.001 |
| **Liver** | | | | | |
| Liver Test Abnormality | SARS-CoV-2 Group | 0.42 | 1.37 | 1.78 | |
| | 2020 Comparator Group | 0.29 | 0.74 | 1.20 | |
| | Risk Difference | 0.13 (-0.03 to 0.32) | 0.63 (0.28 to 0.88) | 0.58 (-0.1 to 0.74) | |
| | p-value | 0.92 | <0.001 | 0.15 | <0.001 |
| **Nephrology** | | | | | |
| Kidney Injury | SARS-CoV-2 Group | 0.10 | 0.43 | 1.05 | |
| | 2020 Comparator Group | 0.06 | 0.25 | 0.58 | |
| | Risk Difference | 0.03 (-0.04 to 0.11) | 0.17 (0.01 to 0.34) | 0.47 (0.14 to 0.65) | |
| | p-value | 1 | <0.001 | <0.001 | <0.001 |

| | | | | | |
|---|---|---|---|---|---|
| > Chronic Kidney Disease | SARS-CoV-2 Group | 0.05 | 0.28 | 0.77 | |
| | 2020 Comparator Group | 0.03 | 0.16 | 0.45 | |
| | Risk Difference | 0.02 (-0.01 to 0.11) | 0.12 (-0.05 to 0.18) | 0.33 (0.16 to 0.6) | |
| | p-value | 0.61 | 1 | <0.001 | <0.001 |
| > Acute Kidney Injury | SARS-CoV-2 Group | 0.08 | 0.28 | 0.79 | |
| | 2020 Comparator Group | 0.04 | 0.14 | 0.31 | |
| | Risk Difference | 0.03 (-0.06 to 0.1) | 0.14 (-0.01 to 0.23) | 0.48 (0.25 to 0.66) | |
| | p-value | 1 | 0.46 | <0.001 | <0.001 |
| **Respiratory** | | | | | |
| Respiratory Failure | SARS-CoV-2 Group | 0.05 | 0.16 | 0.49 | |
| | 2020 Comparator Group | 0.02 | 0.06 | 0.14 | |
| | Risk Difference | 0.03 (-0.01 to 0.09) | 0.1 (0 to 0.18) | 0.35 (0.2 to 0.5) | |
| | p-value | 0.31 | 0.31 | <0.001 | <0.001 |
| > Acute Respiratory Failure | SARS-CoV-2 Group | 0.05 | 0.15 | 0.46 | |
| | 2020 Comparator Group | 0.02 | 0.06 | 0.12 | |
| | Risk Difference | 0.04 (0.01 to 0.09) | 0.09 (0.03 to 0.21) | 0.34 (0.2 to 0.53) | |
| | p-value | <0.001 | <0.001 | <0.001 | <0.001 |
| > Chronic Respiratory Failure | SARS-CoV-2 Group | 0.02 | 0.11 | 0.39 | |
| | 2020 Comparator Group | 0 | 0.01 | 0.03 | |
| | Risk Difference | 0.02 (0 to 0.06) | 0.1 (0.06 to 0.18) | 0.36 (0.26 to 0.5) | |
| | p-value | <0.001 | <0.001 | <0.001 | <0.001 |
| Interstitial Lung Disease | SARS-CoV-2 Group | 0.04 | 0.19 | 0.57 | |
| | 2020 Comparator Group | 0.01 | 0.03 | 0.09 | |
| | Risk Difference | 0.04 (-0.03 to 0.09) | 0.16 (0.05 to 0.25) | 0.48 (0.36 to 0.63) | |
| | p-value | 1 | <0.001 | <0.001 | <0.001 |
| **Dermatologic** | | | | | |
| Atopic Dermatitis | SARS-CoV-2 Group | 0.38 | 0.30 | 0.32 | |
| | 2020 Comparator Group | 0.27 | 0.28 | 0.21 | |
| | Risk Difference | 0.11 (-0.05 to 0.25) | 0.03 (-0.15 to 0.19) | 0.11 (-0.07 to 0.21) | |
| | p-value | 1 | 1 | 1 | 1 |
| Urticaria | SARS-CoV-2 Group | 0.30 | 0.31 | 0.25 | |
| | 2020 Comparator Group | 0.23 | 0.21 | 0.18 | |
| | Risk Difference | 0.07 (-0.1 to 0.26) | 0.11 (-0.06 to 0.2) | 0.07 (-0.04 to 0.25) | |
| | p-value | 1 | 1 | 1 | 1 |
| Herpesviral Vesicular Dermatitis | SARS-CoV-2 Group | 0.10 | 0.14 | 0.09 | |
| | 2020 Comparator Group | 0.09 | 0.13 | 0.08 | |
| | Risk Difference | 0.01 (-0.05 to 0.1) | 0.01 (-0.1 to 0.11) | 0 (-0.07 to 0.08) | |
| | p-value | 1 | 1 | 1 | 1 |

| | | | | | |
|---|---|---|---|---|---|
| **Others** | | | | | |
| Sleep Apnea | SARS-CoV-2 Group | 0.41 | 1.35 | 1.81 | |
| | 2020 Comparator Group | 0.22 | 0.82 | 1.05 | |
| | Risk Difference | 0.19 (0.05 to 0.39) | 0.53 (0.32 to 0.94) | 0.76 (0.55 to 1.32) | |
| | p-value | <0.001 | <0.001 | <0.001 | <0.001 |
| **Symptom** | | | | | |
| Myalgia | SARS-CoV-2 Group | 2.93 | 4.55 | 5.87 | |
| | 2020 Comparator Group | 1.88 | 3.07 | 3.79 | |
| | Risk Difference | 1.05 (0.46 to 1.53) | 1.48 (0.7 to 2.01) | 2.08 (1.1 to 2.89) | |
| | p-value | <0.001 | <0.001 | <0.001 | <0.001 |
| Fatigue Diagnosis | SARS-CoV-2 Group | 3.15 | 5.35 | 5.98 | |
| | 2020 Comparator Group | 1.85 | 2.62 | 2.52 | |
| | Risk Difference | 1.29 (0.83 to 1.88) | 2.73 (2.22 to 3.5) | 3.46 (3.06 to 4.47) | |
| | p-value | <0.001 | <0.001 | <0.001 | <0.001 |
| > Fatigue | SARS-CoV-2 Group | 3.09 | 5.22 | 5.82 | |
| | 2020 Comparator Group | 1.85 | 2.61 | 2.51 | |
| | Risk Difference | 1.24 (0.52 to 1.58) | 2.61 (1.93 to 3.31) | 3.31 (2.54 to 3.97) | |
| | p-value | <0.001 | <0.001 | <0.001 | <0.001 |
| > Chronic Fatigue | SARS-CoV-2 Group | 0.27 | 0.47 | 0.39 | |
| | 2020 Comparator Group | 0.15 | 0.24 | 0.22 | |
| | Risk Difference | 0.12 (-0.01 to 0.22) | 0.23 (0.12 to 0.45) | 0.17 (-0.02 to 0.29) | |
| | p-value | 0.76 | <0.001 | 0.31 | 0.27 |
| > Postviral Fatigue | SARS-CoV-2 Group | 0.10 | 0.29 | 0.30 | |
| | 2020 Comparator Group | 0 | 0.02 | 0.01 | |
| | Risk Difference | 0.1 (0.06 to 0.18) | 0.27 (0.2 to 0.4) | 0.28 (0.19 to 0.39) | |
| | p-value | <0.001 | <0.001 | <0.001 | <0.001 |
| Anosmia | SARS-CoV-2 Group | 0.23 | 0.23 | 0.21 | |
| | 2020 Comparator Group | 0.06 | 0.03 | 0.02 | |
| | Risk Difference | 0.17 (0.08 to 0.32) | 0.2 (0.11 to 0.3) | 0.19 (0.13 to 0.29) | |
| | p-value | <0.001 | <0.001 | <0.001 | 1 |

† Cumulative incidence by Kaplan-Meier estimator 120 days after time origin (index date+21 days), which is 141 days after index date.

† Confidence intervals are calculated based on Bonferroni correction, i.e., (1 - 0.05/N) confidence interval.

†† Individuals were excluded if they had less than index date + 21 days of follow-up, had already been diagnosed with outcome of interest prior to index date + 21 days or their matched pair was excluded for the same reasons.

**Table 4b: Risk Differences (per 100 individuals) for Clinical Sequelae by Gender, UnitedHealth Group Clinical Discovery Database through October 31, 2020.**

| Clinical Sequelae | Category | Female (†) | Male (†) | p-interaction |
|---|---|---|---|---|
| | **Matched Group Size** | **102482 vs 102482 ††** | **92593 vs 92593 ††** | |
| **Hypertension** | | | | |
| Hypertension | SARS-CoV-2 Group | 2.30 | 3.03 | |
| | 2020 Comparator Group | 1.58 | 1.92 | |
| | Risk Difference | 0.72 (0.21 to 1.07) | 1.1 (0.64 to 1.73) | |
| | p-value | <0.001 | <0.001 | 0.14 |
| Pulmonary Hypertension | SARS-CoV-2 Group | 0.14 | 0.14 | |
| | 2020 Comparator Group | 0.05 | 0.06 | |
| | Risk Difference | 0.1 (0.06 to 0.19) | 0.08 (-0.01 to 0.15) | |
| | p-value | <0.001 | 0.1 | 1 |
| **Cardiology** | | | | |
| Arrythmia | SARS-CoV-2 Group | 1.62 | 1.48 | |
| | 2020 Comparator Group | 0.77 | 0.62 | |
| | Risk Difference | 0.85 (0.54 to 1) | 0.86 (0.56 to 1.09) | |
| | p-value | <0.001 | <0.001 | 1 |
| > Cardiac Arrythmia | SARS-CoV-2 Group | 0.75 | 0.71 | |
| | 2020 Comparator Group | 0.38 | 0.29 | |
| | Risk Difference | 0.38 (0.2 to 0.55) | 0.41 (0.2 to 0.51) | |
| | p-value | <0.001 | <0.001 | 1 |
| > Tachycardia | SARS-CoV-2 Group | 1.18 | 1.04 | |
| | 2020 Comparator Group | 0.50 | 0.40 | |
| | Risk Difference | 0.68 (0.45 to 0.82) | 0.64 (0.46 to 0.84) | |
| | p-value | <0.001 | <0.001 | 1 |
| > POTs | SARS-CoV-2 Group | 0.29 | 0.29 | |
| | 2020 Comparator Group | 0.13 | 0.12 | |
| | Risk Difference | 0.16 (0.02 to 0.22) | 0.17 (0.05 to 0.25) | |
| | p-value | <0.001 | <0.001 | 1 |
| Congestive Heart Failure | SARS-CoV-2 Group | 0.15 | 0.31 | |
| | 2020 Comparator Group | 0.06 | 0.10 | |
| | Risk Difference | 0.1 (0 to 0.14) | 0.21 (0.1 to 0.3) | |
| | p-value | 0.1 | <0.001 | 0.05 |
| Cardiomyopathy | SARS-CoV-2 Group | 0.18 | 0.28 | |
| | 2020 Comparator Group | 0.06 | 0.10 | |
| | Risk Difference | 0.12 (0.03 to 0.17) | 0.18 (0.09 to 0.28) | |

| | | | | |
|---|---|---|---|---|
| | p-value | <0.001 | <0.001 | 0.07 |
| Myocarditis | SARS-CoV-2 Group | 0.03 | 0.07 | |
| | 2020 Comparator Group | 0 | 0 | |
| | Risk Difference | 0.02 (-0.01 to 0.05) | 0.06 (0.02 to 0.1) | |
| | p-value | 0.31 | <0.001 | 0.004 |
| Coronary Disease | SARS-CoV-2 Group | 0.15 | 0.30 | |
| | 2020 Comparator Group | 0.07 | 0.17 | |
| | Risk Difference | 0.08 (-0.01 to 0.14) | 0.13 (0.02 to 0.23) | |
| | p-value | 0.31 | <0.001 | 1 |
| > Myocardial Infarction | SARS-CoV-2 Group | 0.08 | 0.20 | |
| | 2020 Comparator Group | 0.04 | 0.11 | |
| | Risk Difference | 0.04 (0 to 0.11) | 0.09 (-0.04 to 0.14) | |
| | p-value | 0.2 | 1 | 1 |
| > Acute Coronary Syndrome | SARS-CoV-2 Group | 0.14 | 0.28 | |
| | 2020 Comparator Group | 0.07 | 0.16 | |
| | Risk Difference | 0.07 (0 to 0.13) | 0.12 (-0.04 to 0.19) | |
| | p-value | 0.31 | 1 | 1 |
| > Cardiogenic Shock | SARS-CoV-2 Group | 0.02 | 0.03 | |
| | 2020 Comparator Group | 0 | 0.01 | |
| | Risk Difference | 0.01 (0 to 0.04) | 0.02 (0 to 0.06) | |
| | p-value | 0.51 | 0.1 | 1 |
| Hypercoagulability DVT PE | SARS-CoV-2 Group | 0.59 | 0.67 | |
| | 2020 Comparator Group | 0.22 | 0.18 | |
| | Risk Difference | 0.37 (0.21 to 0.48) | 0.5 (0.37 to 0.65) | |
| | p-value | <0.001 | <0.001 | 0.13 |
| > Hypercoagulability | SARS-CoV-2 Group | 0.48 | 0.59 | |
| | 2020 Comparator Group | 0.20 | 0.15 | |
| | Risk Difference | 0.27 (0.14 to 0.41) | 0.44 (0.35 to 0.61) | |
| | p-value | <0.001 | <0.001 | <0.001 |
| > Deep Vein Thrombosis | SARS-CoV-2 Group | 0.30 | 0.40 | |
| | 2020 Comparator Group | 0.09 | 0.10 | |
| | Risk Difference | 0.21 (0.12 to 0.32) | 0.3 (0.19 to 0.4) | |
| | p-value | <0.001 | <0.001 | 0.01 |
| > Pulmonary Embolism | SARS-CoV-2 Group | 0.20 | 0.20 | |
| | 2020 Comparator Group | 0.04 | 0.06 | |
| | Risk Difference | 0.16 (0.08 to 0.22) | 0.13 (0.03 to 0.2) | |
| | p-value | <0.001 | <0.001 | 1 |
| Stroke | SARS-CoV-2 Group | 0.22 | 0.23 | |

| | | | | |
|---|---|---|---|---|
| | 2020 Comparator Group | 0.12 | 0.09 | |
| | Risk Difference | 0.1 (0 to 0.16) | 0.14 (0.06 to 0.25) | |
| | p-value | <0.001 | <0.001 | 1 |
| > Ischemic Stroke | SARS-CoV-2 Group | 0.19 | 0.19 | |
| | 2020 Comparator Group | 0.10 | 0.08 | |
| | Risk Difference | 0.1 (0.01 to 0.16) | 0.11 (0.02 to 0.19) | |
| | p-value | <0.001 | <0.001 | 1 |
| > Hemorrhagic Stroke | SARS-CoV-2 Group | 0.05 | 0.07 | |
| | 2020 Comparator Group | 0.03 | 0.01 | |
| | Risk Difference | 0.02 (-0.03 to 0.06) | 0.06 (0.02 to 0.11) | |
| | p-value | 1 | <0.001 | 0.52 |
| **Mental Health** | | | | |
| Mental Health Diagnosis | SARS-CoV-2 Group | 4.79 | 2.59 | |
| | 2020 Comparator Group | 3.85 | 1.80 | |
| | Risk Difference | 0.94 (0.16 to 1.31) | 0.79 (0.3 to 1.11) | |
| | p-value | <0.001 | <0.001 | 1 |
| > Anxiety | SARS-CoV-2 Group | 3.99 | 2.15 | |
| | 2020 Comparator Group | 3.22 | 1.40 | |
| | Risk Difference | 0.77 (0.21 to 1.35) | 0.75 (0.46 to 1.16) | |
| | p-value | <0.001 | <0.001 | 1 |
| > Depression | SARS-CoV-2 Group | 2.17 | 1.10 | |
| | 2020 Comparator Group | 1.84 | 0.85 | |
| | Risk Difference | 0.34 (0 to 0.73) | 0.26 (0.03 to 0.56) | |
| | p-value | 0.1 | <0.001 | 1 |
| > PTSD | SARS-CoV-2 Group | 0.28 | 0.16 | |
| | 2020 Comparator Group | 0.24 | 0.09 | |
| | Risk Difference | 0.04 (-0.11 to 0.16) | 0.07 (-0.01 to 0.18) | |
| | p-value | 1 | 0.2 | 1 |
| **Neurologic** | | | | |
| Amnesia/Memory Difficulty | SARS-CoV-2 Group | 0.73 | 0.69 | |
| | 2020 Comparator Group | 0.32 | 0.31 | |
| | Risk Difference | 0.4 (0.24 to 0.55) | 0.38 (0.21 to 0.51) | |
| | p-value | <0.001 | <0.001 | 1 |
| Migraine | SARS-CoV-2 Group | 1.18 | 0.32 | |
| | 2020 Comparator Group | 0.93 | 0.24 | |
| | Risk Difference | 0.25 (0.11 to 0.67) | 0.08 (-0.05 to 0.2) | |
| | p-value | <0.001 | 1 | 0.31 |
| Peripheral Neuropathy | SARS-CoV-2 Group | 0.28 | 0.34 | |

| | | | | |
|---|---|---|---|---|
| | 2020 Comparator Group | 0.18 | 0.11 | |
| | Risk Difference | 0.1 (0 to 0.23) | 0.23 (0.15 to 0.35) | |
| | p-value | 0.1 | <0.001 | 0.27 |
| Encephalopathy | SARS-CoV-2 Group | 0.20 | 0.27 | |
| | 2020 Comparator Group | 0.03 | 0.06 | |
| | Risk Difference | 0.17 (0.13 to 0.27) | 0.21 (0.11 to 0.28) | |
| | p-value | <0.001 | <0.001 | 0.8 |
| Seizure | SARS-CoV-2 Group | 0.15 | 0.13 | |
| | 2020 Comparator Group | 0.08 | 0.08 | |
| | Risk Difference | 0.07 (0 to 0.14) | 0.05 (-0.01 to 0.13) | |
| | p-value | 0.1 | 0.51 | 1 |
| Dementia | SARS-CoV-2 Group | 0.03 | 0.04 | |
| | 2020 Comparator Group | 0.01 | 0.02 | |
| | Risk Difference | 0.02 (-0.02 to 0.07) | 0.02 (-0.02 to 0.06) | |
| | p-value | 1 | 1 | 1 |
| GuillianBarre Syndrome | SARS-CoV-2 Group | 0.01 | 0.01 | |
| | 2020 Comparator Group | 0 | 0 | |
| | Risk Difference | 0.01 (0 to 0.04) | 0.01 (0 to 0.03) | |
| | p-value | <0.001 | 1 | 1 |
| Alzheimer | SARS-CoV-2 Group | 0.01 | 0 | |
| | 2020 Comparator Group | 0 | 0 | |
| | Risk Difference | 0.01 (-0.01 to 0.02) | 0 (-0.01 to 0.01) | |
| | p-value | 1 | 1 | 1 |
| **Diabetes** | | | | |
| Type 2 Diabetes | SARS-CoV-2 Group | 1.06 | 1.05 | |
| | 2020 Comparator Group | 0.58 | 0.58 | |
| | Risk Difference | 0.48 (0.32 to 0.78) | 0.47 (0.1 to 0.55) | |
| | p-value | <0.001 | <0.001 | 1 |
| **Liver** | | | | |
| Liver Test Abnormality | SARS-CoV-2 Group | 1.23 | 1.17 | |
| | 2020 Comparator Group | 0.74 | 0.66 | |
| | Risk Difference | 0.49 (0.23 to 0.75) | 0.51 (0.23 to 0.74) | |
| | p-value | <0.001 | <0.001 | 1 |
| **Nephrology** | | | | |
| Kidney Injury | SARS-CoV-2 Group | 0.41 | 0.65 | |
| | 2020 Comparator Group | 0.26 | 0.36 | |
| | Risk Difference | 0.15 (0.03 to 0.3) | 0.29 (0.08 to 0.38) | |
| | p-value | <0.001 | <0.001 | 0.02 |

| | | | | |
|---|---|---|---|---|
| > Chronic Kidney Disease | SARS-CoV-2 Group | 0.29 | 0.46 | |
| | 2020 Comparator Group | 0.20 | 0.26 | |
| | Risk Difference | 0.09 (-0.04 to 0.18) | 0.2 (0.09 to 0.38) | |
| | p-value | 1 | <0.001 | 0.07 |
| > Acute Kidney Injury | SARS-CoV-2 Group | 0.30 | 0.49 | |
| | 2020 Comparator Group | 0.11 | 0.20 | |
| | Risk Difference | 0.18 (0.05 to 0.25) | 0.29 (0.18 to 0.46) | |
| | p-value | <0.001 | <0.001 | 0.001 |
| **Respiratory** | | | | |
| Respiratory Failure | SARS-CoV-2 Group | 0.22 | 0.25 | |
| | 2020 Comparator Group | 0.06 | 0.09 | |
| | Risk Difference | 0.16 (0.08 to 0.24) | 0.16 (0.03 to 0.2) | |
| | p-value | <0.001 | <0.001 | 1 |
| > Acute Respiratory Failure | SARS-CoV-2 Group | 0.22 | 0.23 | |
| | 2020 Comparator Group | 0.05 | 0.09 | |
| | Risk Difference | 0.16 (0.09 to 0.25) | 0.14 (0.03 to 0.21) | |
| | p-value | <0.001 | <0.001 | 1 |
| > Chronic Respiratory Failure | SARS-CoV-2 Group | 0.15 | 0.20 | |
| | 2020 Comparator Group | 0.01 | 0.01 | |
| | Risk Difference | 0.14 (0.07 to 0.19) | 0.19 (0.13 to 0.25) | |
| | p-value | <0.001 | <0.001 | 0.52 |
| Interstitial Lung Disease | SARS-CoV-2 Group | 0.23 | 0.32 | |
| | 2020 Comparator Group | 0.06 | 0.06 | |
| | Risk Difference | 0.17 (0.06 to 0.22) | 0.25 (0.15 to 0.35) | |
| | p-value | <0.001 | <0.001 | 0.34 |
| **Dermatologic** | | | | |
| Atopic Dermatitis | SARS-CoV-2 Group | 0.42 | 0.24 | |
| | 2020 Comparator Group | 0.35 | 0.15 | |
| | Risk Difference | 0.07 (-0.15 to 0.14) | 0.09 (-0.03 to 0.19) | |
| | p-value | 1 | 1 | 1 |
| Urticaria | SARS-CoV-2 Group | 0.38 | 0.19 | |
| | 2020 Comparator Group | 0.29 | 0.11 | |
| | Risk Difference | 0.09 (-0.01 to 0.26) | 0.08 (0.02 to 0.18) | |
| | p-value | 0.82 | <0.001 | 1 |
| Herpesviral Vesicular Dermatitis | SARS-CoV-2 Group | 0.14 | 0.07 | |
| | 2020 Comparator Group | 0.11 | 0.05 | |
| | Risk Difference | 0.03 (-0.02 to 0.15) | 0.03 (-0.07 to 0.06) | |
| | p-value | 1 | 1 | 1 |

| | | | | |
|---|---|---|---|---|
| **Others** | | | | |
| Sleep Apnea | SARS-CoV-2 Group | 0.92 | 1.51 | |
| | 2020 Comparator Group | 0.51 | 0.85 | |
| | Risk Difference | 0.41 (0.11 to 0.49) | 0.66 (0.39 to 0.96) | |
| | p-value | <0.001 | <0.001 | 0.008 |
| **Symptom** | | | | |
| Myalgia | SARS-CoV-2 Group | 4.94 | 3.91 | |
| | 2020 Comparator Group | 3.47 | 2.52 | |
| | Risk Difference | 1.47 (0.97 to 2.39) | 1.39 (0.67 to 1.77) | |
| | p-value | <0.001 | <0.001 | 1 |
| Fatigue Diagnosis | SARS-CoV-2 Group | 5.62 | 3.96 | |
| | 2020 Comparator Group | 2.93 | 1.76 | |
| | Risk Difference | 2.69 (2.06 to 3.12) | 2.2 (1.97 to 2.96) | |
| | p-value | <0.001 | <0.001 | 0.02 |
| > Fatigue | SARS-CoV-2 Group | 5.48 | 3.88 | |
| | 2020 Comparator Group | 2.92 | 1.76 | |
| | Risk Difference | 2.56 (1.83 to 3.16) | 2.12 (1.91 to 2.93) | |
| | p-value | <0.001 | <0.001 | 0.05 |
| > Chronic Fatigue | SARS-CoV-2 Group | 0.47 | 0.27 | |
| | 2020 Comparator Group | 0.29 | 0.13 | |
| | Risk Difference | 0.19 (0.03 to 0.29) | 0.14 (0.03 to 0.24) | |
| | p-value | <0.001 | <0.001 | 1 |
| > Postviral Fatigue | SARS-CoV-2 Group | 0.28 | 0.18 | |
| | 2020 Comparator Group | 0.02 | 0 | |
| | Risk Difference | 0.26 (0.16 to 0.34) | 0.17 (0.12 to 0.26) | |
| | p-value | <0.001 | <0.001 | 0.04 |
| Anosmia | SARS-CoV-2 Group | 0.29 | 0.15 | |
| | 2020 Comparator Group | 0.05 | 0.04 | |
| | Risk Difference | 0.24 (0.18 to 0.36) | 0.11 (0.05 to 0.21) | |
| | p-value | <0.001 | <0.001 | <0.001 |

† Cumulative incidence by Kaplan-Meier estimator 120 days after time origin (index date+21 days), which is 141 days after index date.

† Confidence intervals are calculated based on Bonferroni correction, i.e., (1 - 0.05/N) confidence interval.

†† Individuals were excluded if they had less than index date + 21 days of follow-up, had already been diagnosed with outcome of interest prior to index date + 21 days or their matched pair was excluded for the same reasons.

**Table 4c: Risk Differences (per 100 individuals) for Clinical Sequelae by Pre-existing Conditions, UnitedHealth Group Clinical Discovery Database through October 31, 2020.**

| Clinical Sequelae | Category | No pre-existing conditions (†) | With pre-existing conditions (†) | p-interaction |
|---|---|---|---|---|
| | **Matched Group Size** | **82766 vs 82766 ††** | **112919 vs 112919 ††** | |
| **Hypertension** | | | | |
| Hypertension | SARS-CoV-2 Group | 2.11 | 3.26 | |
| | 2020 Comparator Group | 1.22 | 2.33 | |
| | Risk Difference | 0.9 (0.65 to 1.25) | 0.93 (-0.18 to 1.52) | |
| | p-value | <0.001 | 1 | 1 |
| Pulmonary Hypertension | SARS-CoV-2 Group | 0.05 | 0.21 | |
| | 2020 Comparator Group | 0.01 | 0.07 | |
| | Risk Difference | 0.03 (-0.02 to 0.07) | 0.14 (0.08 to 0.22) | |
| | p-value | 1 | <0.001 | <0.001 |
| **Cardiology** | | | | |
| Arrythmia | SARS-CoV-2 Group | 0.95 | 2.01 | |
| | 2020 Comparator Group | 0.32 | 1.04 | |
| | Risk Difference | 0.63 (0.56 to 0.92) | 0.96 (0.59 to 1.12) | |
| | p-value | <0.001 | <0.001 | <0.001 |
| > Cardiac Arrythmia | SARS-CoV-2 Group | 0.39 | 0.97 | |
| | 2020 Comparator Group | 0.15 | 0.54 | |
| | Risk Difference | 0.24 (0.14 to 0.39) | 0.43 (0.21 to 0.57) | |
| | p-value | <0.001 | <0.001 | 0.004 |
| > Tachycardia | SARS-CoV-2 Group | 0.65 | 1.46 | |
| | 2020 Comparator Group | 0.18 | 0.65 | |
| | Risk Difference | 0.46 (0.3 to 0.66) | 0.81 (0.52 to 0.94) | |
| | p-value | <0.001 | <0.001 | <0.001 |
| > POTs | SARS-CoV-2 Group | 0.16 | 0.39 | |
| | 2020 Comparator Group | 0.04 | 0.21 | |
| | Risk Difference | 0.11 (0.04 to 0.2) | 0.18 (0.01 to 0.24) | |
| | p-value | <0.001 | <0.001 | 0.27 |
| Congestive Heart Failure | SARS-CoV-2 Group | 0.08 | 0.34 | |
| | 2020 Comparator Group | 0.01 | 0.14 | |
| | Risk Difference | 0.07 (0.01 to 0.11) | 0.2 (0.07 to 0.27) | |
| | p-value | <0.001 | <0.001 | <0.001 |
| Cardiomyopathy | SARS-CoV-2 Group | 0.08 | 0.34 | |
| | 2020 Comparator Group | 0.03 | 0.15 | |

| | | | | |
|---|---|---|---|---|
| | Risk Difference | 0.05 (0.02 to 0.13) | 0.19 (0.07 to 0.27) | |
| | p-value | <0.001 | <0.001 | <0.001 |
| Myocarditis | SARS-CoV-2 Group | 0.05 | 0.04 | |
| | 2020 Comparator Group | 0 | 0.01 | |
| | Risk Difference | 0.05 (0.02 to 0.09) | 0.03 (-0.01 to 0.05) | |
| | p-value | <0.001 | 0.31 | 1 |
| Coronary Disease | SARS-CoV-2 Group | 0.08 | 0.32 | |
| | 2020 Comparator Group | 0.03 | 0.15 | |
| | Risk Difference | 0.05 (0.02 to 0.13) | 0.17 (0.09 to 0.33) | |
| | p-value | <0.001 | <0.001 | <0.001 |
| > Myocardial Infarction | SARS-CoV-2 Group | 0.04 | 0.20 | |
| | 2020 Comparator Group | 0.02 | 0.10 | |
| | Risk Difference | 0.03 (-0.01 to 0.06) | 0.11 (0.05 to 0.2) | |
| | p-value | 0.61 | <0.001 | 0.003 |
| > Acute Coronary Syndrome | SARS-CoV-2 Group | 0.08 | 0.30 | |
| | 2020 Comparator Group | 0.03 | 0.14 | |
| | Risk Difference | 0.04 (0.01 to 0.11) | 0.15 (0.1 to 0.28) | |
| | p-value | <0.001 | <0.001 | <0.001 |
| > Cardiogenic Shock | SARS-CoV-2 Group | 0.01 | 0.04 | |
| | 2020 Comparator Group | 0 | 0.01 | |
| | Risk Difference | 0.01 (-0.01 to 0.03) | 0.03 (0.01 to 0.08) | |
| | p-value | 1 | <0.001 | 0.09 |
| Hypercoagulability DVT PE | SARS-CoV-2 Group | 0.33 | 0.85 | |
| | 2020 Comparator Group | 0.07 | 0.32 | |
| | Risk Difference | 0.26 (0.16 to 0.37) | 0.53 (0.39 to 0.72) | |
| | p-value | <0.001 | <0.001 | <0.001 |
| > Hypercoagulability | SARS-CoV-2 Group | 0.28 | 0.71 | |
| | 2020 Comparator Group | 0.06 | 0.28 | |
| | Risk Difference | 0.22 (0.11 to 0.32) | 0.43 (0.24 to 0.55) | |
| | p-value | <0.001 | <0.001 | <0.001 |
| > Deep Vein Thrombosis | SARS-CoV-2 Group | 0.16 | 0.48 | |
| | 2020 Comparator Group | 0.03 | 0.17 | |
| | Risk Difference | 0.14 (0.09 to 0.27) | 0.31 (0.23 to 0.45) | |
| | p-value | <0.001 | <0.001 | <0.001 |
| > Pulmonary Embolism | SARS-CoV-2 Group | 0.10 | 0.27 | |
| | 2020 Comparator Group | 0.02 | 0.10 | |
| | Risk Difference | 0.09 (0.04 to 0.16) | 0.17 (0.07 to 0.22) | |
| | p-value | <0.001 | <0.001 | <0.001 |

| | | | | |
|---|---|---|---|---|
| Stroke | SARS-CoV-2 Group | 0.09 | 0.32 | |
| | 2020 Comparator Group | 0.03 | 0.18 | |
| | Risk Difference | 0.06 (0.02 to 0.16) | 0.14 (0.03 to 0.25) | |
| | p-value | <0.001 | <0.001 | 0.007 |
| > Ischemic Stroke | SARS-CoV-2 Group | 0.08 | 0.27 | |
| | 2020 Comparator Group | 0.02 | 0.15 | |
| | Risk Difference | 0.05 (0.01 to 0.13) | 0.13 (0.04 to 0.25) | |
| | p-value | <0.001 | <0.001 | 0.002 |
| > Hemorrhagic Stroke | SARS-CoV-2 Group | 0.03 | 0.08 | |
| | 2020 Comparator Group | 0.01 | 0.05 | |
| | Risk Difference | 0.02 (-0.01 to 0.04) | 0.03 (-0.03 to 0.07) | |
| | p-value | 1 | 1 | 1 |
| **Mental Health** | | | | |
| Mental Health Diagnosis | SARS-CoV-2 Group | 3.02 | 4.30 | |
| | 2020 Comparator Group | 2.16 | 3.47 | |
| | Risk Difference | 0.86 (0.57 to 1.41) | 0.83 (0.06 to 1.59) | |
| | p-value | <0.001 | <0.001 | 1 |
| > Anxiety | SARS-CoV-2 Group | 2.48 | 3.60 | |
| | 2020 Comparator Group | 1.72 | 2.90 | |
| | Risk Difference | 0.75 (0.39 to 1.13) | 0.7 (-0.13 to 1.15) | |
| | p-value | <0.001 | 0.61 | 1 |
| > Depression | SARS-CoV-2 Group | 0.99 | 2.18 | |
| | 2020 Comparator Group | 0.81 | 1.82 | |
| | Risk Difference | 0.18 (-0.01 to 0.48) | 0.35 (0.06 to 0.74) | |
| | p-value | 0.2 | <0.001 | 1 |
| > PTSD | SARS-CoV-2 Group | 0.14 | 0.28 | |
| | 2020 Comparator Group | 0.08 | 0.20 | |
| | Risk Difference | 0.05 (-0.01 to 0.17) | 0.08 (-0.01 to 0.21) | |
| | p-value | 0.51 | 0.2 | 1 |
| **Neurologic** | | | | |
| Amnesia/Memory Difficulty | SARS-CoV-2 Group | 0.39 | 0.94 | |
| | 2020 Comparator Group | 0.13 | 0.38 | |
| | Risk Difference | 0.26 (0.11 to 0.38) | 0.56 (0.42 to 0.74) | |
| | p-value | <0.001 | <0.001 | <0.001 |
| Migraine | SARS-CoV-2 Group | 0.59 | 0.89 | |
| | 2020 Comparator Group | 0.43 | 0.74 | |
| | Risk Difference | 0.16 (0.01 to 0.37) | 0.14 (-0.11 to 0.32) | |
| | p-value | <0.001 | 1 | 1 |

| | | | | |
|---|---|---|---|---|
| Peripheral Neuropathy | SARS-CoV-2 Group | 0.13 | 0.43 | |
| | 2020 Comparator Group | 0.02 | 0.23 | |
| | Risk Difference | 0.1 (0.02 to 0.15) | 0.2 (0.1 to 0.35) | |
| | p-value | <0.001 | <0.001 | 0.43 |
| Encephalopathy | SARS-CoV-2 Group | 0.08 | 0.34 | |
| | 2020 Comparator Group | 0.01 | 0.07 | |
| | Risk Difference | 0.07 (0.03 to 0.14) | 0.27 (0.19 to 0.38) | |
| | p-value | <0.001 | <0.001 | <0.001 |
| Seizure | SARS-CoV-2 Group | 0.06 | 0.20 | |
| | 2020 Comparator Group | 0.04 | 0.11 | |
| | Risk Difference | 0.02 (-0.03 to 0.09) | 0.09 (0 to 0.17) | |
| | p-value | 1 | 0.1 | 0.41 |
| Dementia | SARS-CoV-2 Group | 0.01 | 0.05 | |
| | 2020 Comparator Group | 0 | 0.02 | |
| | Risk Difference | 0.01 (-0.01 to 0.03) | 0.03 (0 to 0.09) | |
| | p-value | 1 | 0.1 | 0.1 |
| GuillianBarre Syndrome | SARS-CoV-2 Group | 0.01 | 0.02 | |
| | 2020 Comparator Group | 0 | 0.01 | |
| | Risk Difference | 0.01 (-0.01 to 0.03) | 0.01 (-0.01 to 0.03) | |
| | p-value | 1 | 1 | 1 |
| Alzheimer | SARS-CoV-2 Group | 0 | 0.01 | |
| | 2020 Comparator Group | 0 | 0.01 | |
| | Risk Difference | 0 (0 to 0) | 0 (-0.01 to 0.02) | |
| | p-value | <0.001 | 1 | 1 |
| **Diabetes** | | | | |
| Type 2 Diabetes | SARS-CoV-2 Group | 0.78 | 1.30 | |
| | 2020 Comparator Group | 0.31 | 0.81 | |
| | Risk Difference | 0.46 (0.26 to 0.59) | 0.49 (0.18 to 0.77) | |
| | p-value | <0.001 | <0.001 | 1 |
| **Liver** | | | | |
| Liver Test Abnormality | SARS-CoV-2 Group | 0.57 | 1.68 | |
| | 2020 Comparator Group | 0.26 | 1.03 | |
| | Risk Difference | 0.31 (0.15 to 0.44) | 0.65 (0.28 to 0.81) | |
| | p-value | <0.001 | <0.001 | <0.001 |
| **Nephrology** | | | | |
| Kidney Injury | SARS-CoV-2 Group | 0.18 | 0.78 | |
| | 2020 Comparator Group | 0.07 | 0.48 | |
| | Risk Difference | 0.1 (0.01 to 0.18) | 0.3 (0.11 to 0.48) | |

| | | | | |
|---|---|---|---|---|
| | p-value | <0.001 | <0.001 | <0.001 |
| > Chronic Kidney Disease | SARS-CoV-2 Group | 0.12 | 0.55 | |
| | 2020 Comparator Group | 0.04 | 0.35 | |
| | Risk Difference | 0.08 (0.02 to 0.14) | 0.21 (0.02 to 0.29) | |
| | p-value | <0.001 | <0.001 | 0.06 |
| > Acute Kidney Injury | SARS-CoV-2 Group | 0.12 | 0.58 | |
| | 2020 Comparator Group | 0.04 | 0.23 | |
| | Risk Difference | 0.08 (0.04 to 0.15) | 0.35 (0.2 to 0.44) | |
| | p-value | <0.001 | <0.001 | <0.001 |
| **Respiratory** | | | | |
| Respiratory Failure | SARS-CoV-2 Group | 0.08 | 0.35 | |
| | 2020 Comparator Group | 0.01 | 0.13 | |
| | Risk Difference | 0.07 (0.02 to 0.12) | 0.22 (0.13 to 0.32) | |
| | p-value | <0.001 | <0.001 | <0.001 |
| > Acute Respiratory Failure | SARS-CoV-2 Group | 0.07 | 0.34 | |
| | 2020 Comparator Group | 0.01 | 0.12 | |
| | Risk Difference | 0.06 (0.03 to 0.13) | 0.22 (0.11 to 0.31) | |
| | p-value | <0.001 | <0.001 | <0.001 |
| > Chronic Respiratory Failure | SARS-CoV-2 Group | 0.06 | 0.26 | |
| | 2020 Comparator Group | 0 | 0.03 | |
| | Risk Difference | 0.06 (0.03 to 0.1) | 0.23 (0.15 to 0.32) | |
| | p-value | <0.001 | <0.001 | <0.001 |
| Interstitial Lung Disease | SARS-CoV-2 Group | 0.12 | 0.39 | |
| | 2020 Comparator Group | 0.01 | 0.09 | |
| | Risk Difference | 0.1 (0.06 to 0.19) | 0.3 (0.19 to 0.4) | |
| | p-value | <0.001 | <0.001 | <0.001 |
| **Dermatologic** | | | | |
| Atopic Dermatitis | SARS-CoV-2 Group | 0.27 | 0.38 | |
| | 2020 Comparator Group | 0.22 | 0.27 | |
| | Risk Difference | 0.05 (-0.04 to 0.17) | 0.11 (-0.05 to 0.23) | |
| | p-value | 1 | 1 | 1 |
| Urticaria | SARS-CoV-2 Group | 0.22 | 0.34 | |
| | 2020 Comparator Group | 0.15 | 0.27 | |
| | Risk Difference | 0.07 (-0.05 to 0.16) | 0.07 (-0.03 to 0.21) | |
| | p-value | 1 | 1 | 1 |
| Herpesviral Vesicular Dermatitis | SARS-CoV-2 Group | 0.09 | 0.12 | |
| | 2020 Comparator Group | 0.08 | 0.12 | |
| | Risk Difference | 0.01 (-0.01 to 0.11) | 0 (-0.06 to 0.09) | |

| | | | | |
|---|---|---|---|---|
| | p-value | 0.51 | 1 | 1 |
| **Others** | | | | |
| Sleep Apnea | SARS-CoV-2 Group | 0.57 | 1.69 | |
| | 2020 Comparator Group | 0.26 | 1.01 | |
| | Risk Difference | 0.31 (0.13 to 0.43) | 0.68 (0.38 to 0.94) | |
| | p-value | <0.001 | <0.001 | <0.001 |
| **Symptom** | | | | |
| Myalgia | SARS-CoV-2 Group | 3.05 | 5.54 | |
| | 2020 Comparator Group | 1.87 | 3.97 | |
| | Risk Difference | 1.18 (0.52 to 1.47) | 1.57 (0.82 to 2.12) | |
| | p-value | <0.001 | <0.001 | 0.09 |
| Fatigue Diagnosis | SARS-CoV-2 Group | 3.43 | 5.93 | |
| | 2020 Comparator Group | 1.41 | 2.98 | |
| | Risk Difference | 2.01 (1.67 to 2.57) | 2.94 (2.37 to 3.5) | |
| | p-value | <0.001 | <0.001 | <0.001 |
| > Fatigue | SARS-CoV-2 Group | 3.34 | 5.79 | |
| | 2020 Comparator Group | 1.41 | 2.97 | |
| | Risk Difference | 1.93 (1.51 to 2.49) | 2.83 (2.36 to 3.47) | |
| | p-value | <0.001 | <0.001 | <0.001 |
| > Chronic Fatigue | SARS-CoV-2 Group | 0.28 | 0.45 | |
| | 2020 Comparator Group | 0.11 | 0.24 | |
| | Risk Difference | 0.17 (0.07 to 0.3) | 0.2 (0.08 to 0.31) | |
| | p-value | <0.001 | <0.001 | 1 |
| > Postviral Fatigue | SARS-CoV-2 Group | 0.16 | 0.29 | |
| | 2020 Comparator Group | 0.01 | 0.03 | |
| | Risk Difference | 0.16 (0.1 to 0.26) | 0.26 (0.16 to 0.3) | |
| | p-value | <0.001 | <0.001 | <0.001 |
| Anosmia | SARS-CoV-2 Group | 0.21 | 0.23 | |
| | 2020 Comparator Group | 0.03 | 0.05 | |
| | Risk Difference | 0.18 (0.12 to 0.29) | 0.18 (0.11 to 0.26) | |
| | p-value | <0.001 | <0.001 | 1 |

† Cumulative incidence by Kaplan-Meier estimator 120 days after time origin (index date+21 days), which is 141 days after index date.

† Confidence intervals are calculated based on Bonferroni correction, i.e., (1 - 0.05/N) confidence interval.

†† Individuals were excluded if they had less than index date + 21 days of follow-up, had already been diagnosed with outcome of interest prior to index date + 21 days or their matched pair was excluded for the same reasons.

**Table 4d: Risk Differences (per 100 individuals) for Clinical Sequelae by Hospitalization Status, UnitedHealth Group Clinical Discovery Database through October 31, 2020.**

| Clinical Sequelae | Category | Not hospitalized with COVID (†) | Hospitalized with COVID (†) | p-interaction |
|---|---|---|---|---|
| | **Matched Group Size** | **176981 vs 176981 ††** | **18118 vs 18118 ††** | |
| **Hypertension** | | | | |
| Hypertension | SARS-CoV-2 Group | 2.29 | 8.28 | |
| | 2020 Comparator Group | 1.49 | 2.72 | |
| | Risk Difference | 0.8 (0.41 to 1.06) | 5.56 (3.31 to 7.76) | |
| | p-value | <0.001 | <0.001 | <0.001 |
| Pulmonary Hypertension | SARS-CoV-2 Group | 0.07 | 0.89 | |
| | 2020 Comparator Group | 0.03 | 0.13 | |
| | Risk Difference | 0.03 (-0.01 to 0.06) | 0.76 (0.48 to 1.05) | |
| | p-value | 1 | <0.001 | <0.001 |
| **Cardiology** | | | | |
| Arrythmia | SARS-CoV-2 Group | 1.17 | 6.41 | |
| | 2020 Comparator Group | 0.70 | 0.95 | |
| | Risk Difference | 0.46 (0.31 to 0.65) | 5.46 (3.96 to 6.11) | |
| | p-value | <0.001 | <0.001 | <0.001 |
| > Cardiac Arrythmia | SARS-CoV-2 Group | 0.54 | 2.80 | |
| | 2020 Comparator Group | 0.35 | 0.48 | |
| | Risk Difference | 0.19 (0.1 to 0.35) | 2.31 (1.58 to 2.82) | |
| | p-value | <0.001 | <0.001 | <0.001 |
| > Tachycardia | SARS-CoV-2 Group | 0.78 | 5.10 | |
| | 2020 Comparator Group | 0.45 | 0.73 | |
| | Risk Difference | 0.33 (0.19 to 0.46) | 4.37 (3.16 to 5.16) | |
| | p-value | <0.001 | <0.001 | <0.001 |
| > POTs | SARS-CoV-2 Group | 0.20 | 1.27 | |
| | 2020 Comparator Group | 0.13 | 0.21 | |
| | Risk Difference | 0.07 (0 to 0.14) | 1.06 (0.51 to 1.29) | |
| | p-value | 0.1 | <0.001 | <0.001 |
| Congestive Heart Failure | SARS-CoV-2 Group | 0.11 | 1.54 | |
| | 2020 Comparator Group | 0.07 | 0.20 | |
| | Risk Difference | 0.04 (-0.01 to 0.09) | 1.34 (0.66 to 1.55) | |
| | p-value | 1 | <0.001 | <0.001 |
| Cardiomyopathy | SARS-CoV-2 Group | 0.12 | 1.28 | |
| | 2020 Comparator Group | 0.08 | 0.23 | |

| | | | | |
|---|---|---|---|---|
| | Risk Difference | 0.04 (-0.01 to 0.1) | 1.06 (0.71 to 1.44) | |
| | p-value | 0.41 | <0.001 | <0.001 |
| Myocarditis | SARS-CoV-2 Group | 0.04 | 0.09 | |
| | 2020 Comparator Group | 0 | 0.01 | |
| | Risk Difference | 0.04 (0.02 to 0.07) | 0.08 (-0.06 to 0.19) | |
| | p-value | <0.001 | 1 | 1 |
| Coronary Disease | SARS-CoV-2 Group | 0.14 | 1.05 | |
| | 2020 Comparator Group | 0.09 | 0.18 | |
| | Risk Difference | 0.05 (-0.01 to 0.12) | 0.87 (0.54 to 1.27) | |
| | p-value | 0.2 | <0.001 | <0.001 |
| > Myocardial Infarction | SARS-CoV-2 Group | 0.08 | 0.70 | |
| | 2020 Comparator Group | 0.05 | 0.12 | |
| | Risk Difference | 0.03 (-0.03 to 0.06) | 0.58 (0.3 to 0.96) | |
| | p-value | 1 | <0.001 | <0.001 |
| > Acute Coronary Syndrome | SARS-CoV-2 Group | 0.13 | 0.94 | |
| | 2020 Comparator Group | 0.09 | 0.17 | |
| | Risk Difference | 0.04 (0 to 0.11) | 0.77 (0.47 to 1.15) | |
| | p-value | 0.1 | <0.001 | <0.001 |
| > Cardiogenic Shock | SARS-CoV-2 Group | 0.01 | 0.20 | |
| | 2020 Comparator Group | 0 | 0.02 | |
| | Risk Difference | 0 (-0.01 to 0.02) | 0.18 (0.03 to 0.34) | |
| | p-value | 1 | <0.001 | <0.001 |
| Hypercoagulability DVT PE | SARS-CoV-2 Group | 0.34 | 3.78 | |
| | 2020 Comparator Group | 0.19 | 0.44 | |
| | Risk Difference | 0.16 (0.06 to 0.24) | 3.34 (2.71 to 4.2) | |
| | p-value | <0.001 | <0.001 | <0.001 |
| > Hypercoagulability | SARS-CoV-2 Group | 0.28 | 3.15 | |
| | 2020 Comparator Group | 0.17 | 0.37 | |
| | Risk Difference | 0.11 (0.05 to 0.21) | 2.78 (2.29 to 3.62) | |
| | p-value | <0.001 | <0.001 | <0.001 |
| > Deep Vein Thrombosis | SARS-CoV-2 Group | 0.16 | 2.28 | |
| | 2020 Comparator Group | 0.10 | 0.30 | |
| | Risk Difference | 0.06 (0.02 to 0.15) | 1.99 (1.52 to 2.46) | |
| | p-value | <0.001 | <0.001 | <0.001 |
| > Pulmonary Embolism | SARS-CoV-2 Group | 0.10 | 1.25 | |
| | 2020 Comparator Group | 0.04 | 0.14 | |
| | Risk Difference | 0.06 (0.02 to 0.1) | 1.11 (0.69 to 1.39) | |
| | p-value | <0.001 | <0.001 | <0.001 |

| | | | | |
|---|---|---|---|---|
| Stroke | SARS-CoV-2 Group | 0.14 | 1.12 | |
| | 2020 Comparator Group | 0.09 | 0.29 | |
| | Risk Difference | 0.05 (0 to 0.1) | 0.83 (0.4 to 1.2) | |
| | p-value | 0.31 | <0.001 | <0.001 |
| > Ischemic Stroke | SARS-CoV-2 Group | 0.12 | 0.94 | |
| | 2020 Comparator Group | 0.07 | 0.22 | |
| | Risk Difference | 0.04 (-0.01 to 0.09) | 0.72 (0.29 to 0.97) | |
| | p-value | 0.51 | <0.001 | <0.001 |
| > Hemorrhagic Stroke | SARS-CoV-2 Group | 0.03 | 0.38 | |
| | 2020 Comparator Group | 0.02 | 0.08 | |
| | Risk Difference | 0.01 (-0.01 to 0.04) | 0.3 (0.11 to 0.56) | |
| | p-value | 1 | <0.001 | <0.001 |
| **Mental Health** | | | | |
| Mental Health Diagnosis | SARS-CoV-2 Group | 3.48 | 5.92 | |
| | 2020 Comparator Group | 2.70 | 2.89 | |
| | Risk Difference | 0.77 (0.32 to 1.09) | 3.03 (1.26 to 4.19) | |
| | p-value | <0.001 | <0.001 | <0.001 |
| > Anxiety | SARS-CoV-2 Group | 2.93 | 4.57 | |
| | 2020 Comparator Group | 2.26 | 2.16 | |
| | Risk Difference | 0.67 (0.4 to 1.04) | 2.41 (1 to 3.16) | |
| | p-value | <0.001 | <0.001 | <0.001 |
| > Depression | SARS-CoV-2 Group | 1.49 | 3.33 | |
| | 2020 Comparator Group | 1.28 | 1.47 | |
| | Risk Difference | 0.21 (0.02 to 0.45) | 1.85 (0.99 to 2.85) | |
| | p-value | <0.001 | <0.001 | <0.001 |
| > PTSD | SARS-CoV-2 Group | 0.19 | 0.51 | |
| | 2020 Comparator Group | 0.18 | 0.18 | |
| | Risk Difference | 0.02 (-0.06 to 0.09) | 0.33 (0.05 to 0.64) | |
| | p-value | 1 | <0.001 | <0.001 |
| **Neurologic** | | | | |
| Amnesia/Memory Difficulty | SARS-CoV-2 Group | 0.50 | 2.90 | |
| | 2020 Comparator Group | 0.30 | 0.43 | |
| | Risk Difference | 0.2 (0.04 to 0.24) | 2.47 (1.76 to 2.96) | |
| | p-value | <0.001 | <0.001 | <0.001 |
| Migraine | SARS-CoV-2 Group | 0.75 | 0.90 | |
| | 2020 Comparator Group | 0.61 | 0.58 | |
| | Risk Difference | 0.14 (-0.04 to 0.27) | 0.31 (-0.03 to 0.87) | |
| | p-value | 0.92 | 0.2 | 1 |

| | | | | |
|---|---|---|---|---|
| Peripheral Neuropathy | SARS-CoV-2 Group | 0.21 | 1.23 | |
| | 2020 Comparator Group | 0.14 | 0.35 | |
| | Risk Difference | 0.07 (-0.02 to 0.14) | 0.87 (0.36 to 1.26) | |
| | p-value | 0.82 | <0.001 | <0.001 |
| Encephalopathy | SARS-CoV-2 Group | 0.07 | 1.87 | |
| | 2020 Comparator Group | 0.04 | 0.08 | |
| | Risk Difference | 0.03 (-0.01 to 0.08) | 1.79 (1.42 to 2.31) | |
| | p-value | 0.41 | <0.001 | <0.001 |
| Seizure | SARS-CoV-2 Group | 0.09 | 0.68 | |
| | 2020 Comparator Group | 0.08 | 0.12 | |
| | Risk Difference | 0.01 (-0.04 to 0.06) | 0.56 (0.32 to 0.88) | |
| | p-value | 1 | <0.001 | <0.001 |
| Dementia | SARS-CoV-2 Group | 0.01 | 0.23 | |
| | 2020 Comparator Group | 0.01 | 0.03 | |
| | Risk Difference | 0 (-0.02 to 0.02) | 0.2 (0.07 to 0.3) | |
| | p-value | 1 | <0.001 | <0.001 |
| GuillianBarre Syndrome | SARS-CoV-2 Group | 0.01 | 0.03 | |
| | 2020 Comparator Group | 0 | 0 | |
| | Risk Difference | 0.01 (-0.02 to 0.03) | 0.03 (0 to 0.09) | |
| | p-value | 1 | <0.001 | 0.05 |
| Alzheimer | SARS-CoV-2 Group | 0 | 0.04 | |
| | 2020 Comparator Group | 0 | 0 | |
| | Risk Difference | 0 (0 to 0.01) | 0.04 (0 to 0.1) | |
| | p-value | 1 | <0.001 | <0.001 |
| **Diabetes** | | | | |
| Type 2 Diabetes | SARS-CoV-2 Group | 0.90 | 3.04 | |
| | 2020 Comparator Group | 0.53 | 0.83 | |
| | Risk Difference | 0.37 (0.24 to 0.58) | 2.21 (1.4 to 3.16) | |
| | p-value | <0.001 | <0.001 | <0.001 |
| **Liver** | | | | |
| Liver Test Abnormality | SARS-CoV-2 Group | 1.01 | 3.30 | |
| | 2020 Comparator Group | 0.63 | 1.36 | |
| | Risk Difference | 0.38 (0.19 to 0.52) | 1.95 (1.06 to 2.58) | |
| | p-value | <0.001 | <0.001 | <0.001 |
| **Nephrology** | | | | |
| Kidney Injury | SARS-CoV-2 Group | 0.32 | 3.02 | |
| | 2020 Comparator Group | 0.28 | 0.79 | |
| | Risk Difference | 0.04 (-0.03 to 0.16) | 2.22 (1.42 to 2.79) | |

| | | | | |
|---|---|---|---|---|
| | p-value | 1 | <0.001 | <0.001 |
| > Chronic Kidney Disease | SARS-CoV-2 Group | 0.21 | 2.06 | |
| | 2020 Comparator Group | 0.19 | 0.70 | |
| | Risk Difference | 0.02 (-0.06 to 0.09) | 1.36 (0.72 to 1.82) | |
| | p-value | 1 | <0.001 | <0.001 |
| > Acute Kidney Injury | SARS-CoV-2 Group | 0.18 | 2.85 | |
| | 2020 Comparator Group | 0.15 | 0.47 | |
| | Risk Difference | 0.03 (-0.04 to 0.09) | 2.38 (1.67 to 3.11) | |
| | p-value | 1 | <0.001 | <0.001 |
| **Respiratory** | | | | |
| Respiratory Failure | SARS-CoV-2 Group | 0.08 | 2.60 | |
| | 2020 Comparator Group | 0.06 | 0.19 | |
| | Risk Difference | 0.02 (-0.03 to 0.07) | 2.41 (1.35 to 3.2) | |
| | p-value | 1 | <0.001 | <0.001 |
| > Acute Respiratory Failure | SARS-CoV-2 Group | 0.07 | 2.58 | |
| | 2020 Comparator Group | 0.05 | 0.18 | |
| | Risk Difference | 0.01 (-0.02 to 0.07) | 2.4 (1.67 to 3.43) | |
| | p-value | 1 | <0.001 | <0.001 |
| > Chronic Respiratory Failure | SARS-CoV-2 Group | 0.04 | 1.53 | |
| | 2020 Comparator Group | 0.01 | 0.05 | |
| | Risk Difference | 0.04 (0.01 to 0.07) | 1.48 (0.97 to 1.75) | |
| | p-value | <0.001 | <0.001 | <0.001 |
| Interstitial Lung Disease | SARS-CoV-2 Group | 0.14 | 1.60 | |
| | 2020 Comparator Group | 0.05 | 0.13 | |
| | Risk Difference | 0.1 (0.02 to 0.14) | 1.47 (1.14 to 1.98) | |
| | p-value | <0.001 | <0.001 | <0.001 |
| **Dermatologic** | | | | |
| Atopic Dermatitis | SARS-CoV-2 Group | 0.34 | 0.27 | |
| | 2020 Comparator Group | 0.24 | 0.26 | |
| | Risk Difference | 0.1 (0.04 to 0.24) | 0.01 (-0.27 to 0.3) | |
| | p-value | <0.001 | 1 | 1 |
| Urticaria | SARS-CoV-2 Group | 0.28 | 0.33 | |
| | 2020 Comparator Group | 0.22 | 0.16 | |
| | Risk Difference | 0.07 (0.01 to 0.19) | 0.17 (-0.06 to 0.43) | |
| | p-value | <0.001 | 0.71 | 1 |
| Herpesviral Vesicular Dermatitis | SARS-CoV-2 Group | 0.11 | 0.05 | |
| | 2020 Comparator Group | 0.09 | 0.09 | |
| | Risk Difference | 0.02 (-0.02 to 0.1) | -0.04 (-0.2 to 0.08) | |

| | | | | |
|---|---|---|---|---|
| | p-value | 1 | 1 | 1 |
| **Others** | | | | |
| Sleep Apnea | SARS-CoV-2 Group | 0.90 | 4.30 | |
| | 2020 Comparator Group | 0.64 | 1.17 | |
| | Risk Difference | 0.26 (0.17 to 0.49) | 3.13 (2.32 to 4.11) | |
| | p-value | <0.001 | <0.001 | <0.001 |
| **Symptom** | | | | |
| Myalgia | SARS-CoV-2 Group | 4.14 | 7.87 | |
| | 2020 Comparator Group | 2.92 | 3.65 | |
| | Risk Difference | 1.22 (0.73 to 1.49) | 4.22 (3.36 to 6.51) | |
| | p-value | <0.001 | <0.001 | <0.001 |
| Fatigue Diagnosis | SARS-CoV-2 Group | 4.33 | 10.16 | |
| | 2020 Comparator Group | 2.22 | 2.78 | |
| | Risk Difference | 2.11 (1.73 to 2.41) | 7.38 (6.87 to 9.87) | |
| | p-value | <0.001 | <0.001 | <0.001 |
| > Fatigue | SARS-CoV-2 Group | 4.22 | 10.01 | |
| | 2020 Comparator Group | 2.22 | 2.76 | |
| | Risk Difference | 2.01 (1.72 to 2.39) | 7.25 (6.25 to 9.66) | |
| | p-value | <0.001 | <0.001 | <0.001 |
| > Chronic Fatigue | SARS-CoV-2 Group | 0.37 | 0.47 | |
| | 2020 Comparator Group | 0.21 | 0.25 | |
| | Risk Difference | 0.16 (0.09 to 0.31) | 0.23 (-0.1 to 0.49) | |
| | p-value | <0.001 | 0.31 | 1 |
| > Postviral Fatigue | SARS-CoV-2 Group | 0.22 | 0.38 | |
| | 2020 Comparator Group | 0.01 | 0.02 | |
| | Risk Difference | 0.21 (0.16 to 0.27) | 0.36 (0.14 to 0.6) | |
| | p-value | <0.001 | <0.001 | 0.002 |
| Anosmia | SARS-CoV-2 Group | 0.23 | 0.17 | |
| | 2020 Comparator Group | 0.04 | 0.07 | |
| | Risk Difference | 0.18 (0.12 to 0.26) | 0.1 (-0.08 to 0.3) | |
| | p-value | <0.001 | 1 | 0.69 |

† Cumulative incidence by Kaplan-Meier estimator 120 days after time origin (index date+21 days), which is 141 days after index date.

† Confidence intervals are calculated based on Bonferroni correction, i.e., (1 - 0.05/N) confidence interval.

†† Individuals were excluded if they had less than index date + 21 days of follow-up, had already been diagnosed with outcome of interest prior to index date + 21 days or their matched pair was excluded for the same reasons.

**Table 4e: Hazard Ratio* for Clinical Sequelae by Period (Jan-June and July-October), UnitedHealth Group Clinical Discovery Database through October 31, 2020.**

| Clinical Sequalae | Category | Index date < 07/01/2020 (N/person-year) † | Index date >= 07/01/2020 (N/person-year) † | p-interaction |
|---|---|---|---|---|
| **Total Matched Size** | | 90377 vs 90377 | 105793 vs 105793 | |
| **Follow-up Median (IQR)** | | 163 (134 – 194) | 79 (48 – 103) | |
| **Hypertension** | | | | |
| Hypertension | SARS-CoV-2 Group | 0.08 (1561 / 18454.11) | 0.08 (748 / 9202.06) | |
| | 2020 Comparator Group | 0.05 (1023 / 18966.80) | 0.05 (479 / 9325.47) | |
| | Hazard Ratio (CI) | 1.59 (1.42 - 1.78) *** | 1.56 (1.35 - 1.82) *** | |
| | p values | <0.001 | <0.001 | 1 |
| Pulmonary hypertension | SARS-CoV-2 Group | 0.004 (157 / 34733.09) | 0.004 (64 / 15735.33) | |
| | 2020 Comparator Group | 0.002 (56 / 34912.30) | 0.001 (20 / 15741.56) | |
| | Hazard Ratio (CI) | 2.73 (1.65 - 4.52) *** | 3.33 (1.55 - 7.18) *** | |
| | p values | <0.001 | <0.001 | 1 |
| **Cardiology** | | | | |
| Arrythmia | SARS-CoV-2 Group | 0.05 (1517 / 30819.90) | 0.05 (690 / 14504.07) | |
| | 2020 Comparator Group | 0.02 (671 / 31254.07) | 0.02 (298 / 14537.65) | |
| | Hazard Ratio (CI) | 2.26 (1.95 - 2.61) *** | 2.29 (1.89 - 2.78) *** | |
| | p values | <0.001 | <0.001 | 1 |
| > Cardiac arrhythmia | SARS-CoV-2 Group | 0.02 (745 / 33179.48) | 0.02 (339 / 15285.98) | |
| | 2020 Comparator Group | 0.01 (358 / 33468.72) | 0.009 (143 / 15289.32) | |
| | Hazard Ratio (CI) | 2.08 (1.69 - 2.55) *** | 2.31 (1.75 - 3.06) *** | |
| | p values | <0.001 | <0.001 | 1 |
| > Tachycardia | SARS-CoV-2 Group | 0.04 (1172 / 32047.61) | 0.03 (490 / 14888.29) | |
| | 2020 Comparator Group | 0.01 (443 / 32424.03) | 0.01 (219 / 14918.38) | |
| | Hazard Ratio (CI) | 2.60 (2.17 - 3.10) *** | 2.30 (1.83 - 2.90) *** | |
| | p values | <0.001 | <0.001 | 1 |
| > POTs | SARS-CoV-2 Group | 0.009 (324 / 34267.55) | 0.009 (141 / 15621.75) | |
| | 2020 Comparator Group | 0.004 (151 / 34483.46) | 0.003 (52 / 15629.95) | |
| | Hazard Ratio (CI) | 2.14 (1.55 - 2.94) *** | 2.63 (1.66 - 4.18) *** | |
| | p values | <0.001 | <0.001 | 1 |
| Congestive heart failure | SARS-CoV-2 Group | 0.007 (241 / 34276.66) | 0.007 (106 / 15593.90) | |
| | 2020 Comparator Group | 0.003 (116 / 34461.48) | 0.002 (29 / 15606.72) | |
| | Hazard Ratio (CI) | 2.07 (1.44 - 2.98) *** | 3.73 (2.06 - 6.76) *** | |
| | p values | <0.001 | <0.001 | 0.94 |

| | | | | |
|---|---|---|---|---|
| Cardiomyopathy | SARS-CoV-2 Group | 0.007 (236 / 34307.08) | 0.007 (105 / 15603.76) | |
| | 2020 Comparator Group | 0.003 (110 / 34500.92) | 0.003 (47 / 15607.65) | |
| | Hazard Ratio (CI) | 2.12 (1.46 - 3.09) *** | 2.71 (1.61 - 4.56) *** | |
| | p values | <0.001 | <0.001 | 1 |
| Myocarditis | SARS-CoV-2 Group | 0.001 (40 / 35026.30) | 0.002 (34 / 15818.51) | |
| | 2020 Comparator Group | 0.000 (2 / 35191.96) | 0.000 (1 / 15820.30) | |
| | Hazard Ratio (CI) | 20.60 (1.89 - 224.01) ** | 42.96 (1.53 - 1205.38) * | |
| | p values | 0.0015 | 0.01 | 1 |
| Coronary disease | SARS-CoV-2 Group | 0.007 (238 / 34326.83) | 0.007 (105 / 15591.58) | |
| | 2020 Comparator Group | 0.003 (105 / 34492.73) | 0.004 (55 / 15597.32) | |
| | Hazard Ratio (CI) | 2.28 (1.56 - 3.33) *** | 1.96 (1.19 - 3.24) *** | |
| | p values | <0.001 | <0.001 | 1 |
| > Myocardial infarction | SARS-CoV-2 Group | 0.004 (151 / 34570.93) | 0.004 (69 / 15665.05) | |
| | 2020 Comparator Group | 0.002 (61 / 34725.00) | 0.002 (38 / 15666.02) | |
| | Hazard Ratio (CI) | 2.53 (1.54 - 4.14) *** | 1.87 (1.01 - 3.46) * | |
| | p values | <0.001 | 0.042 | 1 |
| > Acute coronary syndrome | SARS-CoV-2 Group | 0.006 (222 / 34367.27) | 0.006 (99 / 15602.25) | |
| | 2020 Comparator Group | 0.003 (96 / 34531.77) | 0.004 (55 / 15607.53) | |
| | Hazard Ratio (CI) | 2.33 (1.57 - 3.47) *** | 1.88 (1.13 - 3.14) ** | |
| | p values | <0.001 | 0.0023 | 1 |
| > Cardiogenic shock | SARS-CoV-2 Group | 0.001 (30 / 34993.31) | 0.001 (11 / 15814.76) | |
| | 2020 Comparator Group | 0.000 (12 / 35152.64) | 0.000 (0 / 15817.40) | |
| | Hazard Ratio (CI) | 2.40 (0.81 - 7.12) | 13.99 (0.46 - 423.90) | |
| | p values | 0.42 | 0.55 | <0.001 |
| Hypercoagulability DVT PE | SARS-CoV-2 Group | 0.02 (704 / 33458.32) | 0.02 (260 / 15345.06) | |
| | 2020 Comparator Group | 0.006 (217 / 33753.73) | 0.007 (101 / 15369.33) | |
| | Hazard Ratio (CI) | 3.21 (2.50 - 4.11) *** | 2.50 (1.77 - 3.52) *** | |
| | p values | <0.001 | <0.001 | 1 |
| > Hypercoagulability | SARS-CoV-2 Group | 0.02 (602 / 33720.88) | 0.01 (203 / 15447.83) | |
| | 2020 Comparator Group | 0.006 (196 / 33989.63) | 0.006 (87 / 15471.47) | |
| | Hazard Ratio (CI) | 3.04 (2.33 - 3.95) *** | 2.28 (1.57 - 3.31) *** | |
| | p values | <0.001 | <0.001 | 1 |
| > Deep vein thrombosis | SARS-CoV-2 Group | 0.01 (397 / 34250.85) | 0.010 (151 / 15594.10) | |
| | 2020 Comparator Group | 0.004 (122 / 34487.19) | 0.003 (52 / 15609.98) | |
| | Hazard Ratio (CI) | 3.31 (2.36 - 4.64) *** | 2.70 (1.70 - 4.30) *** | |
| | p values | <0.001 | <0.001 | 1 |
| > Pulmonary embolism | SARS-CoV-2 Group | 0.007 (227 / 34544.31) | 0.007 (107 / 15658.58) | |
| | 2020 Comparator Group | 0.002 (61 / 34747.88) | 0.002 (27 / 15668.56) | |

| | | | | |
|---|---|---|---|---|
| | Hazard Ratio (CI) | 3.72 (2.33 - 5.94) *** | 3.86 (2.03 - 7.35) *** | |
| | p values | <0.001 | <0.001 | 1 |
| Stroke | SARS-CoV-2 Group | 0.008 (263 / 34369.94) | 0.006 (100 / 15619.68) | |
| | 2020 Comparator Group | 0.003 (109 / 34565.57) | 0.003 (43 / 15622.43) | |
| | Hazard Ratio (CI) | 2.40 (1.65 - 3.47) *** | 2.64 (1.52 - 4.59) *** | |
| | p values | <0.001 | <0.001 | 1 |
| > Ischemic stroke | SARS-CoV-2 Group | 0.007 (232 / 34477.01) | 0.006 (87 / 15651.94) | |
| | 2020 Comparator Group | 0.003 (93 / 34671.49) | 0.002 (37 / 15654.80) | |
| | Hazard Ratio (CI) | 2.49 (1.67 - 3.71) *** | 2.63 (1.44 - 4.77) *** | |
| | p values | <0.001 | <0.001 | 1 |
| > Hemorrhagic stroke | SARS-CoV-2 Group | 0.002 (71 / 34881.19) | 0.002 (26 / 15778.02) | |
| | 2020 Comparator Group | 0.001 (22 / 35046.24) | 0.001 (12 / 15778.42) | |
| | Hazard Ratio (CI) | 3.12 (1.42 - 6.86) *** | 2.69 (0.92 - 7.86) | |
| | p values | <0.001 | 0.12 | 1 |
| **Mental Health** | | | | |
| Mental Health Diagnosis | SARS-CoV-2 Group | 0.11 (2671 / 24348.67) | 0.12 (1278 / 10964.56) | |
| | 2020 Comparator Group | 0.07 (1829 / 24684.53) | 0.09 (963 / 11017.52) | |
| | Hazard Ratio (CI) | 1.47 (1.34 - 1.61) *** | 1.34 (1.20 - 1.50) *** | |
| | p values | <0.001 | <0.001 | 1 |
| > Anxiety | SARS-CoV-2 Group | 0.09 (2442 / 26519.19) | 0.10 (1169 / 11921.78) | |
| | 2020 Comparator Group | 0.06 (1691 / 26866.84) | 0.07 (874 / 11983.93) | |
| | Hazard Ratio (CI) | 1.46 (1.33 - 1.61) *** | 1.33 (1.18 - 1.50) *** | |
| | p values | <0.001 | <0.001 | 1 |
| > Depression | SARS-CoV-2 Group | 0.05 (1369 / 29899.01) | 0.05 (723 / 13426.74) | |
| | 2020 Comparator Group | 0.04 (1060 / 30078.17) | 0.04 (590 / 13443.59) | |
| | Hazard Ratio (CI) | 1.31 (1.15 - 1.49) *** | 1.25 (1.07 - 1.46) *** | |
| | p values | <0.001 | <0.001 | 1 |
| > PTSD | SARS-CoV-2 Group | 0.007 (249 / 34423.08) | 0.005 (75 / 15568.58) | |
| | 2020 Comparator Group | 0.005 (161 / 34598.93) | 0.005 (79 / 15565.79) | |
| | Hazard Ratio (CI) | 1.56 (1.12 - 2.17) *** | 1.03 (0.65 - 1.61) | |
| | p values | <0.001 | 1 | 0.5 |
| **Neurologic** | | | | |
| Amnesia/Memory Difficulty | SARS-CoV-2 Group | 0.02 (760 / 33470.58) | 0.02 (360 / 15322.17) | |
| | 2020 Comparator Group | 0.009 (304 / 33732.72) | 0.010 (151 / 15337.69) | |
| | Hazard Ratio (CI) | 2.49 (2.00 - 3.10) *** | 2.61 (1.95 - 3.49) *** | |
| | p values | <0.001 | <0.001 | 1 |
| Migraine | SARS-CoV-2 Group | 0.02 (769 / 32074.79) | 0.02 (325 / 14613.78) | |
| | 2020 Comparator Group | 0.02 (553 / 32312.96) | 0.02 (266 / 14647.27) | |

| | | | | |
|---|---|---|---|---|
| | Hazard Ratio (CI) | 1.37 (1.14 - 1.64) *** | 1.23 (0.97 - 1.55) | |
| | p values | <0.001 | 0.19 | 1 |
| Peripheral neuropathy | SARS-CoV-2 Group | 0.01 (356 / 34412.87) | 0.007 (105 / 15620.44) | |
| | 2020 Comparator Group | 0.005 (165 / 34628.22) | 0.005 (77 / 15631.03) | |
| | Hazard Ratio (CI) | 2.16 (1.59 - 2.93) *** | 1.35 (0.88 - 2.08) | |
| | p values | <0.001 | 1 | 0.45 |
| Encephalopathy | SARS-CoV-2 Group | 0.008 (264 / 34596.12) | 0.007 (107 / 15711.65) | |
| | 2020 Comparator Group | 0.002 (61 / 34792.28) | 0.001 (18 / 15723.76) | |
| | Hazard Ratio (CI) | 4.17 (2.64 - 6.58) *** | 6.25 (2.88 - 13.57) *** | |
| | p values | <0.001 | <0.001 | 1 |
| Seizure | SARS-CoV-2 Group | 0.005 (169 / 34475.54) | 0.004 (58 / 15620.31) | |
| | 2020 Comparator Group | 0.002 (77 / 34660.58) | 0.002 (36 / 15636.87) | |
| | Hazard Ratio (CI) | 2.13 (1.36 - 3.32) *** | 1.81 (0.97 - 3.39) . | |
| | p values | <0.001 | 0.088 | 1 |
| Dementia | SARS-CoV-2 Group | 0.002 (54 / 34965.11) | 0.001 (13 / 15813.05) | |
| | 2020 Comparator Group | 0.000 (12 / 35131.87) | 0.000 (6 / 15815.78) | |
| | Hazard Ratio (CI) | 4.59 (1.62 - 13.04) *** | 2.66 (0.55 - 12.90) | |
| | p values | <0.001 | 1 | 1 |
| GuillianBarre Syndrome | SARS-CoV-2 Group | 0.000 (9 / 35042.30) | 0.000 (6 / 15824.91) | |
| | 2020 Comparator Group | 0.000 (0 / 35202.09) | 0.000 (1 / 15826.43) | |
| | Hazard Ratio (CI) | 72344218.60 (25277507.37 - 207049132.15) *** | 5.00 (0.39 - 64.15) | |
| | p values | <0.001 | 1 | <0.001 |
| Alzheimer | SARS-CoV-2 Group | 0.000 (7 / 35032.21) | 0.000 (2 / 15828.16) | |
| | 2020 Comparator Group | 0.000 (5 / 35192.05) | 0.000 (4 / 15829.47) | |
| | Hazard Ratio (CI) | 1.41 (0.20 - 9.69) | 1.00 (0.10 - 10.28) | |
| | p values | 1 | 1 | 1 |
| **Diabetes** | | | | |
| Type 2 Diabetes | SARS-CoV-2 Group | 0.03 (905 / 26507.78) | 0.03 (356 / 12774.68) | |
| | 2020 Comparator Group | 0.02 (519 / 26933.85) | 0.02 (211 / 12873.56) | |
| | Hazard Ratio (CI) | 1.73 (1.47 - 2.04) *** | 1.90 (1.49 - 2.42) *** | |
| | p values | <0.001 | <0.001 | 1 |
| **Liver** | | | | |
| Liver Test Abnormality | SARS-CoV-2 Group | 0.04 (1197 / 31724.85) | 0.03 (493 / 14713.89) | |
| | 2020 Comparator Group | 0.02 (742 / 32030.02) | 0.02 (293 / 14763.49) | |
| | Hazard Ratio (CI) | 1.64 (1.42 - 1.91) *** | 1.72 (1.39 - 2.13) *** | |
| | p values | <0.001 | <0.001 | 1 |
| **Nephrology** | | | | |
| Kidney Injury | SARS-CoV-2 Group | 0.02 (530 / 32628.65) | 0.02 (246 / 15035.85) | |

| | | | | |
|---|---|---|---|---|
| | 2020 Comparator Group | 0.009 (300 / 32827.90) | 0.009 (128 / 15066.30) | |
| | Hazard Ratio (CI) | 1.73 (1.37 - 2.18) *** | 2.06 (1.49 - 2.83) *** | |
| | p values | <0.001 | <0.001 | 1 |
| > Chronic kidney disease | SARS-CoV-2 Group | 0.01 (395 / 33486.90) | 0.01 (167 / 15296.46) | |
| | 2020 Comparator Group | 0.007 (227 / 33699.84) | 0.006 (94 / 15318.45) | |
| | Hazard Ratio (CI) | 1.69 (1.29 - 2.21) *** | 1.96 (1.34 - 2.87) *** | |
| | p values | <0.001 | <0.001 | 1 |
| > Acute kidney injury | SARS-CoV-2 Group | 0.01 (402 / 33606.46) | 0.01 (200 / 15407.64) | |
| | 2020 Comparator Group | 0.005 (166 / 33792.91) | 0.004 (65 / 15430.79) | |
| | Hazard Ratio (CI) | 2.33 (1.74 - 3.12) *** | 3.19 (2.09 - 4.87) *** | |
| | p values | <0.001 | <0.001 | 1 |
| **Respiratory** | | | | |
| Respiratory Failure | SARS-CoV-2 Group | 0.008 (247 / 32712.14) | 0.007 (109 / 15223.76) | |
| | 2020 Comparator Group | 0.002 (80 / 32947.82) | 0.002 (31 / 15233.42) | |
| | Hazard Ratio (CI) | 3.00 (2.00 - 4.50) *** | 3.66 (2.03 - 6.60) *** | |
| | p values | <0.001 | <0.001 | 1 |
| > Acute respiratory failure | SARS-CoV-2 Group | 0.007 (234 / 32762.28) | 0.007 (108 / 15240.64) | |
| | 2020 Comparator Group | 0.002 (74 / 32992.44) | 0.002 (29 / 15248.57) | |
| | Hazard Ratio (CI) | 3.08 (2.02 - 4.70) *** | 3.98 (2.15 - 7.37) *** | |
| | p values | <0.001 | <0.001 | 1 |
| > Chronic respiratory failure | SARS-CoV-2 Group | 0.006 (194 / 34837.45) | 0.005 (87 / 15778.72) | |
| | 2020 Comparator Group | 0.001 (16 / 35050.65) | 0.000 (3 / 15787.40) | |
| | Hazard Ratio (CI) | 11.74 (5.10 - 27.00) *** | 18.33 (4.60 - 72.99) *** | |
| | p values | <0.001 | <0.001 | 1 |
| Interstitial lung disease | SARS-CoV-2 Group | 0.009 (307 / 34606.68) | 0.008 (121 / 15707.68) | |
| | 2020 Comparator Group | 0.002 (53 / 34843.54) | 0.002 (24 / 15718.75) | |
| | Hazard Ratio (CI) | 6.02 (3.69 - 9.83) *** | 4.79 (2.52 - 9.09) *** | |
| | p values | <0.001 | <0.001 | 1 |
| **Dermatologic** | | | | |
| Atopic dermatitis | SARS-CoV-2 Group | 0.01 (370 / 34110.32) | 0.009 (134 / 15469.60) | |
| | 2020 Comparator Group | 0.008 (282 / 34290.65) | 0.007 (109 / 15478.24) | |
| | Hazard Ratio (CI) | 1.31 (1.01 - 1.69) * | 1.22 (0.84 - 1.77) | |
| | p values | 0.034 | 1 | 1 |
| Urticaria | SARS-CoV-2 Group | 0.008 (281 / 34218.22) | 0.009 (137 / 15512.19) | |
| | 2020 Comparator Group | 0.007 (228 / 34406.74) | 0.005 (82 / 15518.79) | |
| | Hazard Ratio (CI) | 1.24 (0.93 - 1.66) | 1.73 (1.16 - 2.58) *** | |
| | p values | 0.73 | <0.001 | 1 |
| Herpesviral vesicular dermatitis | SARS-CoV-2 Group | 0.003 (101 / 34662.32) | 0.004 (56 / 15677.52) | |

| | | | | |
|---|---|---|---|---|
| | 2020 Comparator Group | 0.003 (109 / 34820.65) | 0.002 (37 / 15680.93) | |
| | Hazard Ratio (CI) | 0.92 (0.59 - 1.46) | 1.50 (0.81 - 2.77) | |
| | p values | 1 | 1 | 1 |
| **Other** | | | | |
| Sleep apnea | SARS-CoV-2 Group | 0.04 (1143 / 30512.97) | 0.04 (492 / 14070.44) | |
| | 2020 Comparator Group | 0.02 (595 / 30863.90) | 0.02 (279 / 14091.26) | |
| | Hazard Ratio (CI) | 1.93 (1.64 - 2.26) *** | 1.77 (1.44 - 2.17) *** | |
| | p values | <0.001 | <0.001 | 1 |
| **Symptoms** | | | | |
| Fatigue Diagnosis | SARS-CoV-2 Group | 0.16 (3867 / 24830.38) | 0.14 (1718 / 11903.12) | |
| | 2020 Comparator Group | 0.07 (1827 / 25627.58) | 0.07 (810 / 12064.19) | |
| | Hazard Ratio (CI) | 2.17 (1.99 - 2.36) *** | 2.05 (1.83 - 2.29) *** | |
| | p values | <0.001 | <0.001 | 1 |
| > Fatigue | SARS-CoV-2 Group | 0.15 (3776 / 24915.78) | 0.14 (1671 / 11927.04) | |
| | 2020 Comparator Group | 0.07 (1822 / 25682.81) | 0.07 (810 / 12083.76) | |
| | Hazard Ratio (CI) | 2.12 (1.95 - 2.31) *** | 2.00 (1.78 - 2.24) *** | |
| | p values | <0.001 | <0.001 | 1 |
| > Chronic Fatigue | SARS-CoV-2 Group | 0.01 (401 / 34163.32) | 0.01 (163 / 15504.16) | |
| | 2020 Comparator Group | 0.006 (192 / 34382.68) | 0.007 (107 / 15512.63) | |
| | Hazard Ratio (CI) | 2.10 (1.58 - 2.80) *** | 1.44 (1.01 - 2.06) * | |
| | p values | <0.001 | 0.031 | 1 |
| > Postviral fatigue | SARS-CoV-2 Group | 0.007 (261 / 34879.26) | 0.008 (120 / 15782.42) | |
| | 2020 Comparator Group | 0.001 (30 / 35107.92) | 0.000 (5 / 15795.46) | |
| | Hazard Ratio (CI) | 8.40 (4.54 - 15.57) *** | 28.61 (6.39 - 128.17) *** | |
| | p values | <0.001 | <0.001 | 1 |
| Myalgia | SARS-CoV-2 Group | 0.14 (3474 / 23987.28) | 0.12 (1376 / 11764.14) | |
| | 2020 Comparator Group | 0.09 (2231 / 24532.99) | 0.08 (1000 / 11848.03) | |
| | Hazard Ratio (CI) | 1.58 (1.46 - 1.72) *** | 1.40 (1.25 - 1.56) *** | |
| | p values | <0.001 | <0.001 | 0.25 |
| Anosmia | SARS-CoV-2 Group | 0.007 (254 / 34449.45) | 0.005 (75 / 15570.61) | |
| | 2020 Comparator Group | 0.001 (39 / 34673.56) | 0.002 (29 / 15580.87) | |
| | Hazard Ratio (CI) | 6.61 (3.77 - 11.57) *** | 2.77 (1.47 - 5.22) *** | |
| | p values | <0.001 | <0.001 | 0.043 |

* Hazard Ratio were utilized to evaluate period effect and account for follow-up periods having variable length.

† 2 decimals are kept for incidence rates >= 0.01 and 3 decimals are kept for incidence rates < 0.01.

**Table 5a: Incidence and Risk Difference (per 100 individuals) for Clinical Sequelae by SARS-CoV-2 Infection and Comparison Group (Index + 14 days), UnitedHealth Group Clinical Discovery Database through October 31, 2020.**

| Outcome | 2020 SARS-CoV-2 Infected † | 2020 Comparator group † | Risk Difference (CI ††) | 2019-matched 2020 SARS-CoV-2 Infected † | 2019 Comparator group † | Risk Difference (CI ††) | Viral LRTI-matched 2020 SARS-CoV-2 Infected † | Viral LRTI comparator group † | Risk Difference (CI ††) |
|---|---|---|---|---|---|---|---|---|---|
| **Matched Group Size** | **213357 †††** | **213357 †††** | | **213828 †††** | **213828 †††** | | **199683 †††** | **199683 †††** | |
| **Follow-up period (days): median (IQR) ††††** | **91 (45 – 127)** | **92 (46 – 128)** | | **91 (45 – 127)** | **91 (45 – 128)** | | **92 (43 – 135)** | **90 (42 – 134)** | |
| **Hypertension** | | | | | | | | | |
| Hypertension | 2.66 | 1.60 | 1.07 (0.81 to 1.30) *** | 2.69 | 1.70 | 0.98 (0.74 to 1.22) *** | 2.68 | 2.55 | 0.13 (-0.15 to 0.40) |
| Pulmonary hypertension | 0.15 | 0.05 | 0.10 (0.06 to 0.14) *** | 0.14 | 0.05 | 0.09 (0.05 to 0.13) *** | 0.15 | 0.17 | -0.02 (-0.08 to 0.02) |
| **Cardiology** | | | | | | | | | |
| Arrythmia | 1.60 | 0.64 | 0.96 (0.82 to 1.10) *** | 1.62 | 0.70 | 0.91 (0.80 to 1.05) *** | 1.66 | 1.15 | 0.50 (0.35 to 0.66) *** |
| > Cardiac arrhythmia | 0.74 | 0.31 | 0.43 (0.34 to 0.53) *** | 0.75 | 0.37 | 0.38 (0.28 to 0.47) *** | 0.76 | 0.53 | 0.23 (0.13 to 0.32) *** |
| > Tachycardia | 1.17 | 0.43 | 0.74 (0.64 to 0.84) *** | 1.17 | 0.44 | 0.74 (0.62 to 0.84) *** | 1.21 | 0.82 | 0.39 (0.26 to 0.50) *** |
| > POTs | 0.31 | 0.13 | 0.18 (0.13 to 0.24) *** | 0.31 | 0.16 | 0.15 (0.09 to 0.20) *** | 0.32 | 0.24 | 0.08 (0.01 to 0.15) *** |
| Congestive heart failure | 0.22 | 0.10 | 0.12 (0.08 to 0.16) *** | 0.22 | 0.10 | 0.12 (0.07 to 0.17) *** | 0.24 | 0.25 | -0.01 (-0.07 to 0.04) |
| Cardiomyopathy | 0.24 | 0.07 | 0.17 (0.12 to 0.21) *** | 0.24 | 0.10 | 0.15 (0.10 to 0.21) *** | 0.24 | 0.22 | 0.02 (-0.05 to 0.08) |
| Myocarditis | 0.05 | 0.00 | 0.05 (0.03 to 0.06) *** | 0.05 | 0.00 | 0.05 (0.03 to 0.06) *** | 0.05 | 0.01 | 0.04 (0.02 to 0.05) *** |

| | | | | | | | | | |
|---|---|---|---|---|---|---|---|---|---|
| Coronary disease | 0.22 | 0.10 | 0.12 (0.07 to 0.17) *** | 0.22 | 0.12 | 0.10 (0.05 to 0.15) *** | 0.23 | 0.23 | -0.00 (-0.05 to 0.05) |
| > Myocardial infarction | 0.14 | 0.06 | 0.08 (0.04 to 0.12) *** | 0.14 | 0.08 | 0.06 (0.03 to 0.10) *** | 0.14 | 0.13 | 0.01 (-0.04 to 0.06) |
| > Acute coronary syndrome | 0.21 | 0.10 | 0.11 (0.07 to 0.16) *** | 0.20 | 0.12 | 0.09 (0.04 to 0.13) *** | 0.21 | 0.21 | -0.00 (-0.06 to 0.05) |
| > Cardiogenic shock | 0.03 | 0.01 | 0.03 (0.01 to 0.04) *** | 0.03 | 0.01 | 0.02 (0.00 to 0.03) *** | 0.03 | 0.03 | 0.00 (-0.02 to 0.02) |
| Hypercoagulability DVT PE | 0.68 | 0.20 | 0.48 (0.40 to 0.56) *** | 0.70 | 0.21 | 0.48 (0.40 to 0.58) *** | 0.72 | 0.47 | 0.25 (0.16 to 0.35) *** |
| > Hypercoagulability | 0.58 | 0.18 | 0.40 (0.32 to 0.47) *** | 0.59 | 0.18 | 0.41 (0.34 to 0.48) *** | 0.61 | 0.43 | 0.18 (0.09 to 0.27) *** |
| > Deep vein thrombosis | 0.40 | 0.10 | 0.29 (0.22 to 0.36) *** | 0.40 | 0.10 | 0.30 (0.25 to 0.36) *** | 0.41 | 0.28 | 0.13 (0.06 to 0.21) *** |
| > Pulmonary embolism | 0.23 | 0.06 | 0.16 (0.13 to 0.20) *** | 0.23 | 0.06 | 0.17 (0.12 to 0.21) *** | 0.24 | 0.15 | 0.10 (0.05 to 0.16) *** |
| Stroke | 0.24 | 0.10 | 0.13 (0.08 to 0.18) *** | 0.24 | 0.14 | 0.10 (0.04 to 0.15) *** | 0.24 | 0.19 | 0.05 (-0.01 to 0.10) |
| > Ischemic stroke | 0.20 | 0.09 | 0.11 (0.06 to 0.17) *** | 0.20 | 0.11 | 0.09 (0.04 to 0.13) *** | 0.20 | 0.16 | 0.04 (-0.01 to 0.10) |
| > Hemorrhagic stroke | 0.07 | 0.03 | 0.04 (0.01 to 0.07) *** | 0.07 | 0.03 | 0.04 (0.01 to 0.07) *** | 0.07 | 0.05 | 0.01 (-0.02 to 0.04) |
| **Mental Health** | | | | | | | | | |
| Mental Health Diagnosis | 3.71 | 2.61 | 1.10 (0.84 to 1.38) *** | 3.72 | 2.46 | 1.25 (1.02 to 1.53) *** | 3.82 | 3.17 | 0.65 (0.34 to 0.96) *** |
| > Anxiety | 3.15 | 2.16 | 0.98 (0.76 to 1.19) *** | 3.16 | 1.98 | 1.18 (0.97 to 1.41) *** | 3.21 | 2.51 | 0.69 (0.47 to 0.92) *** |
| > Depression | 1.61 | 1.25 | 0.37 (0.23 to 0.53) *** | 1.63 | 1.31 | 0.32 (0.17 to 0.47) *** | 1.68 | 1.63 | 0.05 (-0.13 to 0.22) |
| > PTSD | 0.23 | 0.15 | 0.08 (0.02 to 0.13) *** | 0.21 | 0.16 | 0.05 (-0.00 to 0.10) . | 0.23 | 0.16 | 0.07 (0.01 to 0.13) *** |
| **Neurologic** | | | | | | | | | |
| Amnesia/Memory Difficulty | 0.72 | 0.31 | 0.41 (0.32 to 0.52) *** | 0.74 | 0.35 | 0.39 (0.30 to 0.46) *** | 0.76 | 0.49 | 0.27 (0.18 to 0.38) *** |
| Migraine | 0.77 | 0.58 | 0.18 (0.08 to 0.30) *** | 0.75 | 0.62 | 0.13 (0.04 to 0.24) *** | 0.78 | 0.79 | -0.01 (-0.12 to 0.11) |
| Peripheral neuropathy | 0.29 | 0.15 | 0.14 (0.08 to 0.21) *** | 0.29 | 0.16 | 0.13 (0.07 to 0.19) *** | 0.30 | 0.19 | 0.11 (0.05 to 0.17) *** |

| | | | | | | | | | |
|---|---|---|---|---|---|---|---|---|---|
| Encephalopathy | 0.26 | 0.04 | 0.22 (0.17 to 0.26) *** | 0.25 | 0.07 | 0.18 (0.14 to 0.23) *** | 0.27 | 0.16 | 0.10 (0.05 to 0.16) *** |
| Seizure | 0.14 | 0.08 | 0.07 (0.03 to 0.11) *** | 0.14 | 0.11 | 0.03 (-0.01 to 0.07) | 0.15 | 0.14 | 0.00 (-0.04 to 0.05) |
| Dementia | 0.03 | 0.01 | 0.02 (0.00 to 0.04) *** | 0.03 | 0.02 | 0.01 (-0.00 to 0.03) | 0.04 | 0.03 | 0.00 (-0.02 to 0.03) |
| GuillianBarre Syndrome | 0.02 | 0.01 | 0.01 (-0.00 to 0.02) | 0.01 | 0.00 | 0.01 (-0.00 to 0.02) | 0.02 | 0.01 | 0.01 (-0.00 to 0.03) |
| Alzheimer | 0.01 | 0.00 | 0.00 (-0.01 to 0.01) | 0.00 | 0.00 | 0.00 (-0.01 to 0.01) | 0.01 | 0.00 | 0.00 (-0.01 to 0.01) |
| **Diabetes** | | | | | | | | | |
| Type 2 Diabetes | 1.04 | 0.54 | 0.50 (0.39 to 0.62) *** | 1.02 | 0.61 | 0.41 (0.29 to 0.53) *** | 1.06 | 0.79 | 0.27 (0.13 to 0.38) *** |
| **Liver** | | | | | | | | | |
| Liver Test Abnormality | 1.18 | 0.66 | 0.51 (0.40 to 0.61) *** | 1.19 | 0.70 | 0.49 (0.38 to 0.61) *** | 1.22 | 1.07 | 0.15 (0.01 to 0.29) . |
| **Nephrology** | | | | | | | | | |
| Kidney Injury | 0.55 | 0.32 | 0.23 (0.15 to 0.32) *** | 0.56 | 0.32 | 0.23 (0.14 to 0.32) *** | 0.57 | 0.51 | 0.06 (-0.03 to 0.17) |
| > Chronic kidney disease | 0.38 | 0.24 | 0.14 (0.06 to 0.21) *** | 0.39 | 0.23 | 0.17 (0.10 to 0.23) *** | 0.39 | 0.33 | 0.06 (-0.00 to 0.14) |
| > Acute kidney injury | 0.42 | 0.15 | 0.27 (0.21 to 0.33) *** | 0.42 | 0.18 | 0.24 (0.18 to 0.31) *** | 0.43 | 0.35 | 0.08 (0.00 to 0.16) *** |
| **Respiratory** | | | | | | | | | |
| Respiratory Failure | 0.27 | 0.06 | 0.21 (0.16 to 0.26) *** | 0.28 | 0.10 | 0.17 (0.11 to 0.22) *** | 0.28 | 0.30 | -0.02 (-0.09 to 0.04) |
| > Acute respiratory failure | 0.26 | 0.06 | 0.21 (0.16 to 0.25) *** | 0.27 | 0.10 | 0.17 (0.13 to 0.23) *** | 0.27 | 0.28 | -0.02 (-0.08 to 0.03) |
| > Chronic respiratory failure | 0.19 | 0.01 | 0.18 (0.15 to 0.22) *** | 0.20 | 0.02 | 0.17 (0.14 to 0.21) *** | 0.20 | 0.12 | 0.09 (0.04 to 0.14) *** |
| Interstitial lung disease | 0.29 | 0.04 | 0.25 (0.20 to 0.31) *** | 0.29 | 0.05 | 0.23 (0.19 to 0.28) *** | 0.29 | 0.26 | 0.03 (-0.03 to 0.09) |
| **Dermatologic** | | | | | | | | | |
| Atopic dermatitis | 0.31 | 0.27 | 0.04 (-0.05 to 0.10) | 0.33 | 0.28 | 0.05 (-0.02 to 0.12) | 0.32 | 0.28 | 0.04 (-0.03 to 0.11) |
| Urticaria | 0.30 | 0.20 | 0.10 (0.05 to 0.16) *** | 0.29 | 0.25 | 0.04 (-0.02 to 0.12) | 0.29 | 0.37 | -0.07 (-0.16 to 0.00) |
| Herpesviral vesicular dermatitis | 0.10 | 0.07 | 0.03 (-0.01 to 0.07) | 0.11 | 0.10 | 0.01 (-0.04 to 0.04) | 0.10 | 0.12 | -0.02 (-0.05 to 0.03) |
| **Others** | | | | | | | | | |
| Sleep apnea | 1.17 | 0.67 | 0.50 (0.35 to 0.62) *** | 1.18 | 0.74 | 0.44 (0.31 to 0.56) *** | 1.20 | 1.39 | -0.19 (-0.34 to -0.04) |
| **Symptoms** | | | | | | | | | |

| | | | | | | | | | |
|---|---|---|---|---|---|---|---|---|---|
| Fatigue Diagnosis | 4.85 | 2.35 | 2.50 (2.23 to 2.78) *** | 4.93 | 2.38 | 2.55 (2.28 to 2.78) *** | 5.08 | 3.77 | 1.31 (1.02 to 1.59) *** |
| > Fatigue | 4.73 | 2.34 | 2.38 (2.13 to 2.61) *** | 4.79 | 2.36 | 2.43 (2.16 to 2.67) *** | 4.94 | 3.75 | 1.19 (0.88 to 1.53) *** |
| > Chronic Fatigue | 0.38 | 0.21 | 0.17 (0.11 to 0.25) *** | 0.39 | 0.23 | 0.16 (0.09 to 0.23) *** | 0.39 | 0.32 | 0.07 (-0.02 to 0.15) |
| > Postviral fatigue | 0.24 | 0.01 | 0.23 (0.19 to 0.27) *** | 0.26 | 0.01 | 0.24 (0.20 to 0.30) *** | 0.26 | 0.04 | 0.22 (0.18 to 0.27) *** |
| Myalgia | 4.34 | 2.82 | 1.52 (1.25 to 1.80) *** | 4.38 | 3.26 | 1.12 (0.84 to 1.41) *** | 4.51 | 4.29 | 0.23 (-0.06 to 0.51) |
| Anosmia | 0.25 | 0.04 | 0.21 (0.16 to 0.26) *** | 0.24 | 0.02 | 0.22 (0.17 to 0.26) *** | 0.25 | 0.04 | 0.21 (0.17 to 0.25) *** |

† Cumulative incidence by Kaplan-Meier estimator 120 days after time origin (index date+14 days), which is 134 days after index date.

†† Confidence intervals are calculated based on Bonferroni correction, i.e., (1 - 0.05/N) confidence interval.

††† Individuals were excluded if they had less than index date + 14 days of follow-up, had already been diagnosed with outcome of interest prior to index date + 14 days or their matched pair was excluded for the same reasons.

†††† The follow-up period (days) is measured from time origin (index date + 14 days) to end of observation of the member.

*** p value < 0.001
** p value < 0.01
* p value < 0.05

**Table 5b: Incidence and Risk Difference (per 100 individuals) for Clinical Sequelae by SARS-CoV-2 Infection and 2020 Comparison Group (Index+28 days), UnitedHealth Group Clinical Discovery Database through October 31, 2020.**

| Outcome | 2020 SARS-CoV-2 Infected † | 2020 Comparator group † | Risk Difference (CI ††) | 2019-matched 2020 SARS-CoV-2 Infected † | 2019 Comparator group † | Risk Difference (CI ††) | Viral LRTI-matched 2020 SARS-CoV-2 Infected † | Viral LRTI comparator group † | Risk Difference (CI ††) |
|---|---|---|---|---|---|---|---|---|---|
| **Matched Group Size** | **175435 †††** | **175435 †††** | | **175826 †††** | **175826 †††** | | **165673 †††** | **165673 †††** | |
| **Follow-up period (days): median (IQR) ††††** | **82 (45 – 122)** | **84 (46 – 121)** | | **82 (45 – 122)** | **82 (45 – 121)** | | **85 (44 – 129)** | **83 (41 – 124)** | |
| **Hypertension** | | | | | | | | | |
| Hypertension | 2.55 | 1.68 | 0.87 (0.60 to 1.13) *** | 2.60 | 1.69 | 0.91 (0.64 to 1.19) *** | 2.62 | 2.39 | 0.23 (-0.05 to 0.57) |
| Pulmonary hypertension | 0.14 | 0.04 | 0.10 (0.06 to 0.16) *** | 0.13 | 0.05 | 0.08 (0.03 to 0.12) *** | 0.14 | 0.16 | -0.01 (-0.07 to 0.04) |
| **Cardiology** | | | | | | | | | |
| Arrythmia | 1.41 | 0.64 | 0.76 (0.63 to 0.90) *** | 1.44 | 0.71 | 0.73 (0.56 to 0.86) *** | 1.45 | 1.07 | 0.38 (0.20 to 0.55) *** |
| > Cardiac arrhythmia | 0.70 | 0.32 | 0.38 (0.27 to 0.49) *** | 0.70 | 0.39 | 0.32 (0.21 to 0.43) *** | 0.70 | 0.50 | 0.19 (0.08 to 0.31) *** |
| > Tachycardia | 0.99 | 0.44 | 0.55 (0.44 to 0.67) *** | 1.00 | 0.42 | 0.58 (0.46 to 0.69) *** | 1.04 | 0.74 | 0.30 (0.14 to 0.42) *** |
| > POTs | 0.27 | 0.13 | 0.14 (0.08 to 0.20) *** | 0.27 | 0.15 | 0.11 (0.04 to 0.17) *** | 0.28 | 0.22 | 0.06 (-0.01 to 0.12) |
| Congestive heart failure | 0.21 | 0.09 | 0.12 (0.07 to 0.18) *** | 0.20 | 0.10 | 0.10 (0.05 to 0.16) *** | 0.22 | 0.21 | 0.01 (-0.05 to 0.07) |
| Cardiomyopathy | 0.22 | 0.07 | 0.14 (0.10 to 0.20) *** | 0.22 | 0.09 | 0.13 (0.07 to 0.18) *** | 0.22 | 0.20 | 0.02 (-0.03 to 0.09) |
| Myocarditis | 0.04 | 0.00 | 0.04 (0.02 to 0.06) *** | 0.04 | 0.00 | 0.04 (0.02 to 0.06) *** | 0.04 | 0.01 | 0.03 (0.01 to 0.05) *** |

| | | | | | | | | | |
|---|---|---|---|---|---|---|---|---|---|
| Coronary disease | 0.20 | 0.10 | 0.10 (0.04 to 0.16) *** | 0.20 | 0.13 | 0.08 (0.02 to 0.13) *** | 0.22 | 0.21 | 0.00 (-0.06 to 0.07) |
| > Myocardial infarction | 0.12 | 0.06 | 0.06 (0.02 to 0.11) *** | 0.13 | 0.08 | 0.05 (0.01 to 0.09) *** | 0.13 | 0.12 | 0.01 (-0.04 to 0.06) |
| > Acute coronary syndrome | 0.19 | 0.10 | 0.09 (0.04 to 0.16) *** | 0.19 | 0.12 | 0.07 (0.01 to 0.14) *** | 0.20 | 0.20 | -0.00 (-0.07 to 0.07) |
| > Cardiogenic shock | 0.02 | 0.00 | 0.02 (0.00 to 0.03) *** | 0.02 | 0.01 | 0.01 (-0.00 to 0.03) | 0.03 | 0.02 | 0.00 (-0.02 to 0.03) |
| Hypercoagulability DVT PE | 0.57 | 0.20 | 0.37 (0.28 to 0.46) *** | 0.58 | 0.20 | 0.38 (0.30 to 0.47) *** | 0.60 | 0.40 | 0.20 (0.11 to 0.29) *** |
| > Hypercoagulability | 0.48 | 0.17 | 0.31 (0.23 to 0.39) *** | 0.50 | 0.17 | 0.32 (0.24 to 0.40) *** | 0.52 | 0.37 | 0.15 (0.04 to 0.26) *** |
| > Deep vein thrombosis | 0.31 | 0.10 | 0.21 (0.14 to 0.28) *** | 0.31 | 0.09 | 0.22 (0.14 to 0.28) *** | 0.33 | 0.23 | 0.10 (0.03 to 0.18) *** |
| > Pulmonary embolism | 0.18 | 0.06 | 0.12 (0.08 to 0.17) *** | 0.17 | 0.06 | 0.11 (0.07 to 0.17) *** | 0.18 | 0.12 | 0.06 (0.01 to 0.12) *** |
| Stroke | 0.21 | 0.10 | 0.11 (0.06 to 0.16) *** | 0.20 | 0.14 | 0.06 (-0.00 to 0.13) | 0.21 | 0.17 | 0.04 (-0.02 to 0.11) |
| > Ischemic stroke | 0.18 | 0.08 | 0.10 (0.05 to 0.15) *** | 0.17 | 0.12 | 0.05 (0.00 to 0.10) . | 0.19 | 0.15 | 0.04 (-0.02 to 0.10) |
| > Hemorrhagic stroke | 0.05 | 0.03 | 0.02 (-0.01 to 0.05) | 0.05 | 0.04 | 0.01 (-0.01 to 0.04) | 0.05 | 0.04 | 0.01 (-0.02 to 0.04) |
| **Mental Health** | | | | | | | | | |
| Mental Health Diagnosis | 3.45 | 2.62 | 0.84 (0.58 to 1.11) *** | 3.50 | 2.45 | 1.06 (0.80 to 1.34) *** | 3.66 | 3.13 | 0.52 (0.23 to 0.83) *** |
| > Anxiety | 2.92 | 2.15 | 0.77 (0.49 to 1.03) *** | 2.99 | 1.94 | 1.05 (0.83 to 1.28) *** | 3.05 | 2.49 | 0.56 (0.31 to 0.80) *** |
| > Depression | 1.54 | 1.28 | 0.26 (0.08 to 0.44) *** | 1.55 | 1.32 | 0.23 (0.05 to 0.40) *** | 1.63 | 1.63 | 0.01 (-0.19 to 0.22) |
| > PTSD | 0.23 | 0.13 | 0.10 (0.04 to 0.17) *** | 0.22 | 0.16 | 0.05 (-0.01 to 0.12) | 0.22 | 0.16 | 0.07 (0.01 to 0.13) *** |
| **Neurologic** | | | | | | | | | |
| Amnesia/Memory Difficulty | 0.66 | 0.31 | 0.34 (0.24 to 0.43) *** | 0.68 | 0.35 | 0.33 (0.24 to 0.45) *** | 0.70 | 0.45 | 0.24 (0.14 to 0.35) *** |
| Migraine | 0.73 | 0.57 | 0.16 (0.03 to 0.28) *** | 0.71 | 0.61 | 0.10 (-0.04 to 0.20) | 0.75 | 0.78 | -0.03 (-0.16 to 0.09) |
| Peripheral neuropathy | 0.32 | 0.15 | 0.17 (0.09 to 0.24) *** | 0.31 | 0.16 | 0.16 (0.08 to 0.22) *** | 0.33 | 0.21 | 0.12 (0.03 to 0.20) *** |
| Encephalopathy | 0.20 | 0.03 | 0.17 (0.13 to 0.22) *** | 0.20 | 0.07 | 0.13 (0.09 to 0.18) *** | 0.21 | 0.13 | 0.08 (0.01 to 0.13) *** |

| | | | | | | | | | |
|---|---|---|---|---|---|---|---|---|---|
| Seizure | 0.14 | 0.07 | 0.07 (0.02 to 0.12) *** | 0.13 | 0.09 | 0.04 (-0.01 to 0.08) | 0.14 | 0.13 | 0.01 (-0.04 to 0.06) |
| Dementia | 0.03 | 0.01 | 0.02 (-0.00 to 0.04) . | 0.03 | 0.02 | 0.01 (-0.02 to 0.03) | 0.04 | 0.04 | 0.00 (-0.02 to 0.03) |
| GuillianBarre Syndrome | 0.01 | 0.00 | 0.01 (-0.01 to 0.02) | 0.01 | 0.00 | 0.01 (-0.00 to 0.02) | 0.01 | 0.00 | 0.01 (0.00 to 0.03) *** |
| Alzheimer | 0.01 | 0.00 | 0.00 (-0.00 to 0.02) | 0.01 | 0.00 | 0.00 (-0.01 to 0.01) | 0.00 | 0.00 | 0.00 (-0.01 to 0.01) |
| **Diabetes** | | | | | | | | | |
| Type 2 Diabetes | 1.04 | 0.57 | 0.47 (0.32 to 0.63) *** | 0.99 | 0.60 | 0.40 (0.25 to 0.56) *** | 1.06 | 0.73 | 0.33 (0.16 to 0.48) *** |
| **Liver** | | | | | | | | | |
| Liver Test Abnormality | 1.17 | 0.69 | 0.47 (0.34 to 0.61) *** | 1.18 | 0.70 | 0.47 (0.33 to 0.60) *** | 1.22 | 0.99 | 0.23 (0.07 to 0.37) *** |
| **Nephrology** | | | | | | | | | |
| Kidney Injury | 0.48 | 0.32 | 0.16 (0.05 to 0.25) *** | 0.50 | 0.34 | 0.17 (0.08 to 0.27) *** | 0.50 | 0.46 | 0.05 (-0.06 to 0.14) |
| > Chronic kidney disease | 0.34 | 0.25 | 0.09 (0.01 to 0.17) *** | 0.37 | 0.23 | 0.14 (0.05 to 0.21) *** | 0.36 | 0.30 | 0.06 (-0.03 to 0.15) |
| > Acute kidney injury | 0.34 | 0.14 | 0.20 (0.13 to 0.27) *** | 0.35 | 0.19 | 0.17 (0.09 to 0.25) *** | 0.36 | 0.32 | 0.04 (-0.04 to 0.14) |
| **Respiratory** | | | | | | | | | |
| Respiratory Failure | 0.21 | 0.06 | 0.14 (0.09 to 0.19) *** | 0.21 | 0.10 | 0.12 (0.07 to 0.17) *** | 0.21 | 0.25 | -0.04 (-0.10 to 0.04) |
| > Acute respiratory failure | 0.20 | 0.06 | 0.14 (0.09 to 0.19) *** | 0.20 | 0.09 | 0.11 (0.05 to 0.16) *** | 0.20 | 0.24 | -0.04 (-0.10 to 0.03) |
| > Chronic respiratory failure | 0.16 | 0.02 | 0.14 (0.11 to 0.17) *** | 0.16 | 0.02 | 0.14 (0.10 to 0.18) *** | 0.17 | 0.10 | 0.07 (0.02 to 0.12) *** |
| Interstitial lung disease | 0.27 | 0.04 | 0.22 (0.18 to 0.27) *** | 0.25 | 0.05 | 0.20 (0.14 to 0.25) *** | 0.26 | 0.22 | 0.04 (-0.03 to 0.12) |
| **Dermatologic** | | | | | | | | | |
| Atopic dermatitis | 0.31 | 0.28 | 0.03 (-0.05 to 0.10) | 0.32 | 0.27 | 0.05 (-0.02 to 0.12) | 0.33 | 0.28 | 0.05 (-0.02 to 0.13) |
| Urticaria | 0.27 | 0.19 | 0.08 (0.01 to 0.15) *** | 0.27 | 0.24 | 0.04 (-0.04 to 0.11) | 0.28 | 0.36 | -0.08 (-0.15 to -0.00) |
| Herpesviral vesicular dermatitis | 0.10 | 0.07 | 0.04 (-0.00 to 0.08) . | 0.10 | 0.10 | 0.00 (-0.05 to 0.05) | 0.10 | 0.12 | -0.02 (-0.06 to 0.02) |
| **Others** | | | | | | | | | |
| Sleep apnea | 1.19 | 0.65 | 0.54 (0.41 to 0.68) *** | 1.17 | 0.75 | 0.41 (0.27 to 0.56) *** | 1.24 | 1.30 | -0.06 (-0.23 to 0.12) |
| **Symptoms** | | | | | | | | | |

| | | | | | | | | | |
|---|---|---|---|---|---|---|---|---|---|
| Fatigue Diagnosis | 4.50 | 2.38 | 2.11 (1.85 to 2.39) *** | 4.48 | 2.29 | 2.20 (1.93 to 2.46) *** | 4.70 | 3.55 | 1.14 (0.80 to 1.49) *** |
| > Fatigue | 4.41 | 2.38 | 2.03 (1.75 to 2.32) *** | 4.39 | 2.28 | 2.11 (1.83 to 2.44) *** | 4.60 | 3.54 | 1.05 (0.70 to 1.36) *** |
| > Chronic Fatigue | 0.40 | 0.22 | 0.19 (0.11 to 0.27) *** | 0.41 | 0.23 | 0.18 (0.11 to 0.27) *** | 0.40 | 0.33 | 0.06 (-0.02 to 0.15) |
| > Postviral fatigue | 0.21 | 0.01 | 0.20 (0.16 to 0.25) *** | 0.21 | 0.01 | 0.20 (0.15 to 0.25) *** | 0.21 | 0.02 | 0.19 (0.15 to 0.24) *** |
| Myalgia | 4.39 | 2.89 | 1.50 (1.21 to 1.90) *** | 4.36 | 3.25 | 1.12 (0.79 to 1.47) *** | 4.58 | 4.21 | 0.36 (0.01 to 0.69) *** |
| Anosmia | 0.21 | 0.04 | 0.17 (0.11 to 0.22) *** | 0.21 | 0.02 | 0.19 (0.14 to 0.25) *** | 0.21 | 0.04 | 0.18 (0.12 to 0.23) *** |

† Cumulative incidence by Kaplan-Meier estimator 120 days after time origin (index date+28 days), which is 148 days after index date.

†† Confidence intervals are calculated based on Bonferroni correction, i.e., (1 - 0.05/N) confidence interval.

††† Individuals were excluded if they had less than index date + 28 days of follow-up, had already been diagnosed with outcome of interest prior to index date + 28 days or their matched pair was excluded for the same reasons.

†††† The follow-up period (days) is measured from time origin (index date + 28 days) to end of observation of the member.

*** p value < 0.001
** p value < 0.01
* p value < 0.05

**Figure 2: Cumulative Hazard Rate of PCP Visit by SARS-CoV-2 Diagnostic Method and Three Comparator Groups, UnitedHealth Group Clinical Research Discovery Database through October 31, 2020.**

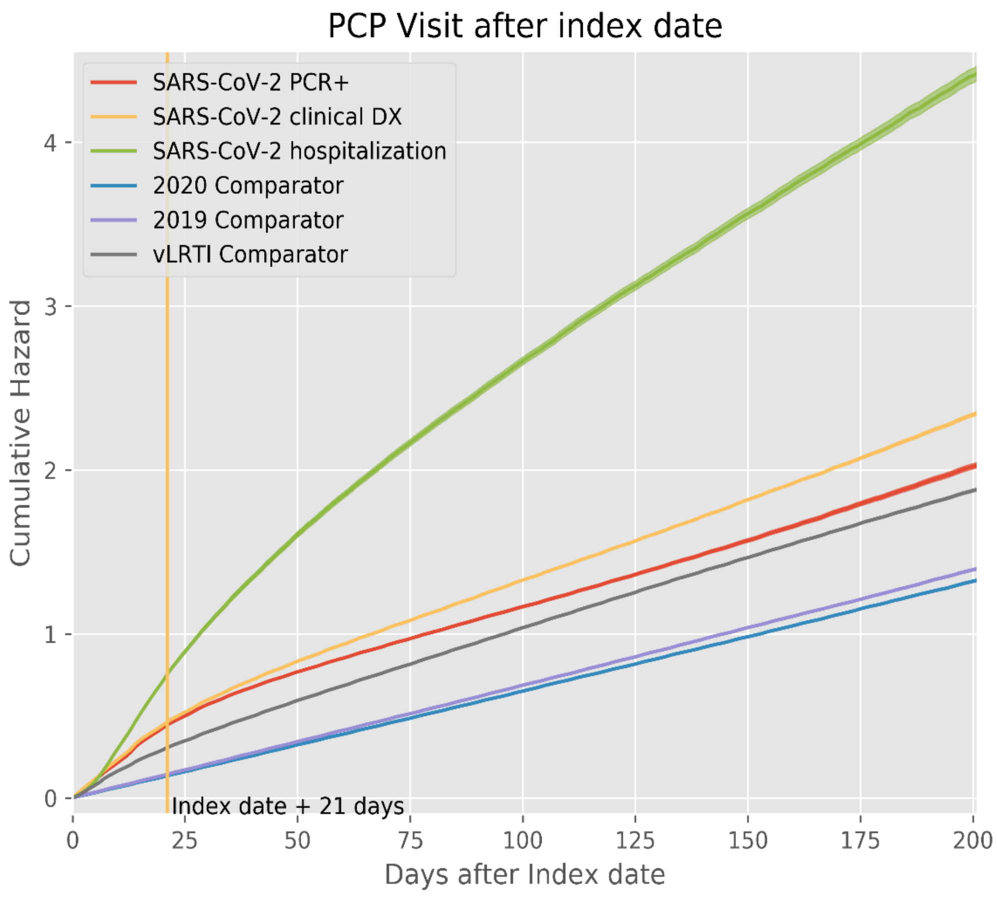

** PCP visits were considered as recurring events and Nelson-Aalen estimator for matched groups was applied.*
